# Integrative Electrophysiology and Neuroimaging Approach in Assessing Disorders of Consciousness: A Multimodal Multicentric Machine Learning Study

**DOI:** 10.1101/2024.11.22.24317805

**Authors:** Dragana Manasova, Laouen Mayal Louan Belloli, Martin Rosenfelder, Lina Willacker, Emilia Flo Rama, Chiara Valota, Bertrand Hermann, Brigitte Charlotte Kaufmann, Alice Pirastru, Chiara Camilla Derchi, Theresa Raiser, Melanie Valente, Aude Sangare, Başak Türker, Nadya Pyatigorskaya, Benoît Béranger, Michele Colombo, Esteban Munoz-Musat, Anira Escrichs, Tiziana Atzori, Francesca Baglio, Constantin Lapa, Ansgar Berlis, Kristina Krüger, Tina Luther, Vincent Perlbarg, Gustavo Deco, Yonathan Sanz-Perl, Enzo Tagliazucchi, Louis Puybasset, Benjamin Rohaut, Lionel Naccache, Angela Comanducci, Anat Arzi, Mario Rosanova, Andreas Bender, Jacobo Diego Sitt

**Author notes:** Correspondence to: Dragana Manasova and Jacobo Diego Sitt Paris Brain Institute - Institut du Cerveau, Hôpital Pitié, 47 Bd de l’Hôpital, 75013 Paris, France &.

## Abstract

Severely brain-injured patients may enter a spectrum of conditions collectively known as disorders of consciousness (DoC). This spectrum includes clinical categories such as unresponsive wakefulness syndrome or minimally conscious state, where the behavioral assessment of consciousness can often be deceptive.

To bridge this dissociation, neuroimaging techniques are employed to look for the residual brain functions. Each neuroimaging modality imperfectly captures distinct aspects of brain preservation - functional, anatomical, or both. In this study, we adopt a comprehensive approach by integrating the neurophysiology and neuroimaging modalities available from the standard and advanced clinical assessment through interpretable machine learning (ML). The electrophysiological modalities included high-density electroencephalography (EEG) (resting state and task), whereas neuroimaging modalities included anatomical and resting-state functional magnetic resonance imaging (MRI), diffusion MRI, and 18F-fluoro-deoxy-glucose positron emission tomography (FDG PET).

Our investigation reveals that specific modalities, such as functional assessments provide comprehensive insights into the currently evaluated state of consciousness - the diagnosis of the patients. Conversely, structural modalities offer valuable information about the patient’s evolution within the consciousness spectrum. We validate the proposed analysis with data coming from other centers with different acquisition parameters. Importantly, we show that there is an improved model performance with the increase in the number of modalities. We observe a higher inter-modality disagreement for MCS patients and those patients who improve. Lastly, we observe a difference in feature importances in diagnosis and prognosis.

This integrative multimodal and ML methodology presents a promising avenue for a more nuanced understanding of DoC, contributing to enhanced diagnostic precision and prognostic capabilities in clinical practice.

## Introduction

Disorders of consciousness (DoC) are a spectrum of conditions arising from numerous causes of brain injury. Patients with a DoC have a range of sensory-motor deficits that can differently impair their state of consciousness and behavioral capacity to various extents.^1^ Due to an incapacity to express consciousness through behavioral responses, there can be a dissociation between unresponsiveness (based on following commands with motor outputs) and unawareness.^2,3^ Furthermore, the assessment of patients with DoC can be limited due to the presence of medical devices (such as mechanical ventilation or tracheostomy tubes), acute pain, or medications that affect arousal.^4^ All of these aspects pose challenges to the correct assessment of the patient’s consciousness.

The main clinical categories on the DoC spectrum are the Unresponsive Wakefulness Syndrome (UWS) and the Minimally Conscious State (MCS). UWS patients are behaviorally diagnosed by eyes opening during arousal but with no signs of awareness^5^, whereas patients in an MCS show reproducible, though subtle, behavioral signs of consciousness (visual pursuit or a response to simple commands)^6^. Although there is no clear consensus where the spectrum of DoC ends and healthy consciousness begins^7^, generally patients are said to be emergent from MCS (EMCS) when they regain some basic communication capacity or when they are capable of functional object use^6^. This recovery of consciousness can occur at any point in the patient’s clinical evolution –from the acute to the chronic stage^7^.

The current diagnostic gold standard in the field is the Coma Recovery Scale-Revised (CRS-R)^8^. Although repeatedly using this scale decreases the rate of misdiagnosis^8^, the remaining uncertainty due to behavioral and neural disparities has yet to be systematically addressed. Thus, despite the extensive standardization of the administration of behavioral scales^9^, current guidelines recommend the use of neuronal or physiological signals across distinct modalities to increase the certainty of a correct assessment of a patient’s state^4,10–13^. In addition, individual electrophysiology or neuroimaging modalities are limited because they occupy a narrow space in the temporal-spatial resolution plane, and are designed to evaluate only part of the neural structures or activities. A natural question that arises is to what extent each modality is informative in terms of diagnosis and prognosis.

Throughout the years, various studies have focused on single modalities, and have shown the potential of each one in improving the patient’s diagnosis.^2,14–32^ However, clinical teams, especially those in hospitals with established expertise in treating DoC patients, have access to and can integrate the information of various multimodal tests.^11,33^ Recent evidence indicates that multimodal assessment enhances neuroprognostics in clinically unresponsive critical-care patients with brain injury.^33^ This suggests that latent integration of information by clinicians contributes to improved decision-making outcomes, thereby making a strong argument for the exploration of automatic fusion approaches using machine learning (ML). This argument is also reflected in the international guidelines for the treatment of DoC, calling for multimodal assessments especially due to the heterogeneous pathophysiology of DoC patients.^10,13,34^ On the question of how we can systematically make use of the complementary information contained in the different neurophysiological signals, numerous studies have assessed the possibilities of an integrative neuroimaging approach.^11,35–42^ These studies highlight the need to investigate various dimensions of brain preservation (for example anatomical and functional MRI, electrophysiology, or brain metabolism) in order to make a more accurate assessment of the patients’ current state and evolution. Yet, up to date there hasn’t been a large-scale multi-center study, involving commonly used neuroimaging modalities analyzed under the same methodological umbrella, to evaluate the differences and complementarity of the modalities assessing the patients’ current condition and evolution.

In our study, we take the multimodal integrative neuroimaging approach one step further and adopt a multicentric, comprehensive approach by separately analyzing and then integrating six neuroimaging modalities through interpretable ML, to investigate the patients’ presently evaluated consciousness (diagnosis) and future evolution (prognosis).

## Methods

This project is part of an EU-funded four-year consortium (PerBrain) involving several different institutions including the Pitié Salpetrière Hospital, the University Hospital of the Ludwig-Maximilians-University of Munich, the Therapiezentrum Burgau (hospital for neurological rehabilitation), the University of Milan, Fondazione Don Carlo Gnocchi and the Weizmann Institute of Science. The details of the consortium’s aims are explained in Willacker et al. (2022). The included patients come from three different centers split in Dataset 1 (France), Dataset 2 (Germany), and Dataset 3 (Italy).

### Ethics statements

This research was approved by the ethical committee of the Pitie-Salpetriere under the French label of ‘routine care research’ (Comité de Protection des Personnes n◦ 2013-A01385-40, Ile de France 1, Paris, France under the code ‘Recherche en soins courants’, protocol number M-Neuro-DOC, CE SRLF 20-2); the ethics committee of the medical faculty of the Ludwig-Maximilians-Universität München (protocol numbers 20-634 and 20-635); and the ethical committee section of the IRCCS Fondazione Don Carlo Gnocchi (ethics committee IRCCS Regione Lombardia, protocol number 32/2021/CE_FdG/FC/SA). Written informed consent from patients was obtained either through their legal guardian or, in the absence of one, from the closest relative.

### Patient inclusion and behavioral assessments

In the primary Dataset 1 (France), we included 326 (120 women, mean age 46.5 +/- 17.7 years) patients with a DoC who stayed at the Pitié Salpetrière Hospital for an expert assessment of their state of consciousness between 2008 and 2022. During this evaluation, the clinical team performed several behavioral or neuroimaging exams (clinical assessment, MRI, EEG, PET), as well as tracked the patients’ evolution. Each patient underwent neuroimaging acquisitions tailored to their specific clinical needs, whereas EEG RS and LG recordings were acquired as part of the standard clinical practice. The CRS-R^8^ was performed by expert clinicians during the patients’ stay at the hospital to assess their current state. Typically the multimodal assessments were done within a range of one week during the expert evaluation. We took the best CRS-R score of the patients during this clinical assessment and only kept the patients that were diagnosed as being in UWS and patients in an MCS. Throughout the text under ‘current’ or ‘present’ state, we refer to the evaluation of the patients using the CRS-R which is based on the best evaluation over an extended window of time. According to the CRS-R scores, 153 patients were diagnosed as UWS, and 173 as MCS. The behavioral scale used to infer the evolution of the patients after hospitalization is the Glasgow Outcome Scale-Extended (GOSE)^43^ together with a phone-guided CRS-R evaluation. The assessments were done through telephone interviews with the patient’s current physician (if they are still in a DoC) or family member (in case they have recovered consciousness). The best previously completed CRS-R (during the hospital stay) is taken as a starting point, and the clinician checks in each CRS-R category if the patient has improved, stagnated, or worsened. Using these assessments, the patients then have a GOSE score together with a CRS-R state of consciousness category at six months post-evaluation, one year, and two years. From the behavioral assessments during-hospitalization and post-hospitalization, we derive two prognostic categories of patients who have either not improved or worsened over two years, or patients who have improved (Figure 1 A). We exclude patients who have gone through a limitation of active therapy (life-sustaining care). The etiologies and demographics of the patients varied, and their summaries are given in Supplementary Tables 1 and 2.

**Figure 1.**
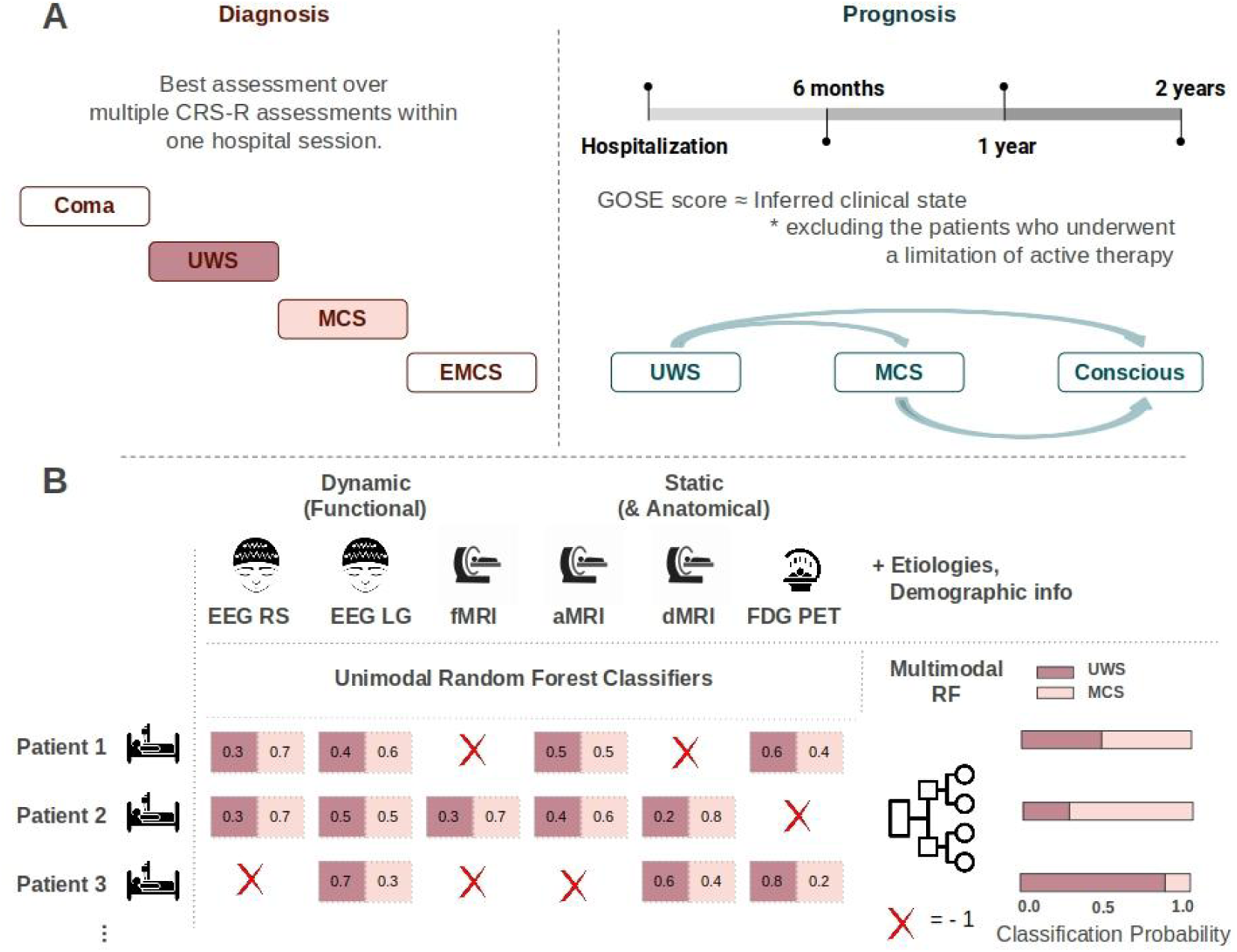
Multimodal assessment methods of diagnostic and prognostic categories of patients with DoC. (**A**) Patients underwent multiple CRS-R assessments during their hospital stay and the best assessment in that range of a week (typically) was taken as the gold-standard clinical diagnosis. The diagnostic clinical categories of patients included in the prediction are UWS and MCS. The prognostic categories are improved or not improved (explained in the Methods section). (**B**) For both the diagnostic and prognostic classification we ran unimodal RFC to obtain probabilistic estimates of each patient belonging to one or another category. The probabilistic outputs are then combined using a second-level RFC either alone or in combination with the etiologies and demographic information. Missing values are substituted with -1 (a data imputation approach). The final output is a probability of belonging to either a diagnostic or prognostic category. Abbreviations: Coma Recovery Scale-Revised (CRS-R), Unresponsive Wakefulness Syndrome (UWS), Minimally Conscious State (MCS), Emergent Minimally Conscious State (EMCS), Glasgow Outcome Scale-Extended (GOSE), Electroencephalography (EEG), Resting State (RS), Local Global (LG) paradigm, functional Magnetic Resonance Imaging (fMRI), anatomical Magnetic Resonance Imaging (aMRI), diffusion Magnetic Resonance Imaging (dMRI), 18F-fluoro-Deoxy-Glucose Positron Emission Tomography (FDG PET), Random Forest Classifiers (RFC).

**Table 1.** Overview of the number of patients from the Paris Dataset 1 having a particular neuroimaging modality per prediction category (diagnostic or prognostic).

| Modality | All patients | Diagnostic categories<br>(Current state of the patients) |  | Prognostic categories<br>(Evolution of the patients) |  |
| --- | --- | --- | --- | --- | --- |
|  |  | UWS | MCS | not-Improved | Improved |
| EEG RS | 120 | 63 | 57 | 50 | 39 |
| EEG LG | 290 | 138 | 152 | 117 | 90 |
| fMRI RS | 44 | 21 | 23 | 20 | 10 |
| FDG PET | 53 | 21 | 32 | 35 | 9 |
| dMRI | 151 | 79 | 72 | 49 | 46 |
| aMRI | 101 | 54 | 47 | 40 | 26 |

The table presents data obtained through various neuroimaging modalities for a total of 326 patients for diagnosis, and 232 patients with prognostic data. Modalities include electroencephalography resting state (EEG RS), EEG local-global paradigm (EEG LG), resting-state functional MRI (fMRI RS), anatomical MRI (aMRI), diffusion MRI (dMRI), and 18F-fluoro-Deoxy-Glucose Positron Emission Tomography (FDG PET). The patient population is further categorized based on diagnostic information, distinguishing between those in a vegetative state/unresponsive wakefulness state (UWS) and those in a minimally conscious state (MCS). Prognostic insights into patient evolution are also provided, indicating the number of patients showing improvement and those not showing improvement across different modalities. The counts in each cell represent the corresponding number of patients within the specified modality and diagnostic/prognostic category. Not all the patients have prognostic information, thus the sum of the prognostic categories is not equal to the counts in the All Patients column. Information on the patients' sex, etiology, age, and whether they were acute or chronic at the time of the tests is given in Supplementary Tables 1 and 2. Information on the Datasets 2 and 3 from Germany and Italy is presented in Supplementary tables 3 and 4.

**Table 2.** Overview of multimodal neuroimaging markers used as features.

| Modality type | Modality | Paradigm | Metrics |
| --- | --- | --- | --- |
| Dynamic, functional | EEG | Resting state | Spectral, information theory, connectivity markers |
| Dynamic, functional | EEG | Local-Global Auditory Task | Spectral, information theory, connectivity and evoked markers |
| Dynamic, functional | fMRI | Resting state | Cortical & subcortical functional connectivity |
| Static, functional | FDG PET | Resting state | Cortical & subcortical metabolic activity |
| Static, anatomical | dMRI | Resting state | White matter tract fractional anisotropy and mean diffusivity |
| Static, anatomical | aMRI | Resting state | Cortical thickness, subcortical volume |

This table provides a comprehensive overview of various multimodal neuroimaging modalities, encompassing both dynamic or static and anatomical or functional properties. Each modality is detailed with its associated paradigm and metric. Dynamic modalities, which have multiple time points, include EEG in the resting state and task local-global paradigms, as well as fMRI in the resting state. Static modalities include FDG PET, dMRI, and aMRI. The second type refers to whether the modality captures anatomical properties (aMRI and dMRI) or functional ones (EEG, fMRI, FDG PET). Each modality is associated with specific metrics such as cortical and subcortical functional connectivity, metabolic activity, white matter tract properties, and structural measures like cortical thickness and subcortical volume. Abbreviations: Electroencephalography (EEG), functional Magnetic Resonance Imaging (fMRI), anatomical Magnetic Resonance Imaging (aMRI), diffusion Magnetic Resonance Imaging (dMRI), 18F-fluoro-Deoxy-Glucose Positron Emission Tomography (FDG PET).

From Dataset 2 (Germany), we included 54 (16 women, age 56 +- 15.3) patients with DoC who stayed either at the University Hospital of the Ludwig-Maximilians-Universität München (Munich, Germany) or the Therapiezentrum Burgau (Hospital for Neurological Rehabilitation, Burgau, Germany) for an expert assessment of their state of consciousness and for acute neurological care or early neurological rehabilitation between 2020 and 2023. According to the CRS-R scores, 34 patients were diagnosed with UWS, and 20 were diagnosed with MCS. The etiologies and demographics of the patients varied, and their summaries are given in Supplementary Table 3.

From Dataset 3 (Italy), we included 30 (13 women, age 46.6 +- 16.9) patients with DoC who stayed at the IRCCS Fondazione Don Carlo Gnocchi for an expert assessment of their state of consciousness between 2020 and 2023. According to the CRS-R scores, 13 patients were diagnosed with UWS, and 17 were diagnosed with MCS. The etiologies and demographics of the patients varied, and their summaries are given in Supplementary Table 4.

### Modalities

In this study, the included modalities are high-density EEG resting state (RS) and a two levels Local-Global (LG) auditory regularity task, anatomical- and resting-state functional-magnetic resonance imaging (aMRI and RS-fMRI), diffusion MRI (dMRI), and 18F-fluoro-Deoxy-Glucose Positron Emission Tomography (FDG PET) (Table 2). The acquisition protocols and parameters, the preprocessing details, as well as the analyses of the markers are given in the Supplementary Materials. The markers we extracted from the modalities later used in the ML models are given in Table 2.

### Machine learning models

In the main analysis, we included 326 patients from Dataset 1 who had assessments from one to five different modalities (Table 1), while the unimodal generalization ability was tested on Dataset 2 and Dataset 3 (Supplementary Tables 3 and 4). We have a two by two focus of the analyses, looking into the unimodal and multimodal results both for the diagnostic and prognostic categories.

#### Unimodal models

In the first level analysis, we ran unimodal models either classifying the diagnostic (UWS or MCS) or prognostic (improved or not improved) categories. We used a Random Forest Classifier (RFC), which is an ensemble tree-based method, from the scikit-learn library (v1.2.0) with the default hyper-parameters (100 estimators, gini criterion, no max depth, 2 minimum samples split).^44^ For our goal, tree-based models seem to be the most adequate as they capture nonlinear relationships between features and labels. Tree-based models do not require feature scaling which is an important advantage when working with multimodal data. On Dataset 1, we ran 100 initializations of a five-fold stratified cross-validation, keeping the predictions from the fold that is used for testing. The resulting prediction and probability of a patient belonging to a given clinical category were collected over these 100 iterations. To understand whether the use of probability estimates was reasonable, we ran model calibration checks (Supplementary Figure 1). In both the diagnostic and prognostic classification, most models are sufficiently well calibrated except for aMRI for the prognosis. We used the scikit-learn impurity-based (Gini) feature importance (FI) algorithm. In each iteration of the models, we saved the FI and then averaged them across the 100 initializations to obtain a ranking of the features per modality. For the generalization, after z-scoring the features of both the training and testing datasets, we trained 100 new models per modality on all of the patients from Dataset 1 and then tested on the patients from Dataset 2 and 3. We also obtained a surrogate distribution per modality by shuffling the labels and running again the 100 iterations, 5-fold cross-validated (CV) training, and testing. We compared the two distributions of the model results and the surrogates using divergence metrics (Kullback-Leibler divergence, Jensen-Shannon divergence, Wasserstein distance) (Supplementary Figure 2). To quantify classification disparity between modalities within a patient, we created a metric called pairwise disagreement (PD), calculated as the difference in classification probabilities between two modalities for the same patient (Figure 3A).

**Figure 2.**
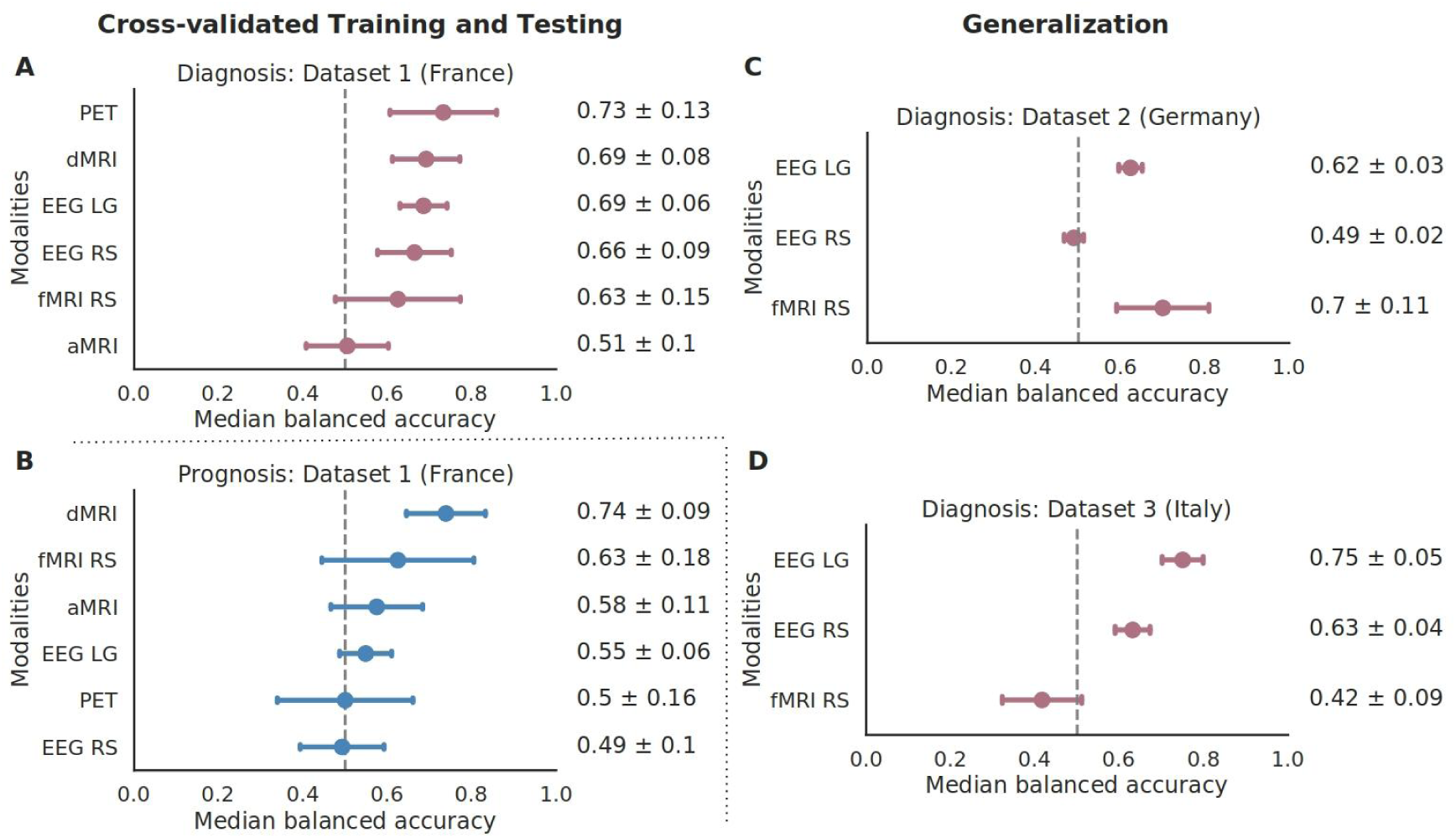
Multivariate unimodal models’ accuracy differs depending on the classification target that can be diagnostic or prognostic. (**A**) Unimodal Random Forest classifiers for the six modalities give a diagnostic classification accuracy which is given next to the distributions (median +/- standard deviation). The diagnostic classification patient categories are patients in UWS or MCS. (**B**) The same as in (**A**) but for the prognostic categories (improved and not improved). (**C**) and (**D**), same as (**A**) but for the diagnostic classification trained on Dataset 1: France and tested on the available modalities from Dataset 2: Germany and Dataset 3: Italy. Abbreviations: Electroencephalography (EEG), Resting State (RS), Local Global (LG) paradigm, functional Magnetic Resonance Imaging (fMRI), anatomical Magnetic Resonance Imaging (aMRI), diffusion Magnetic Resonance Imaging (dMRI), Positron Emission Tomography (PET).

**Figure 3.**
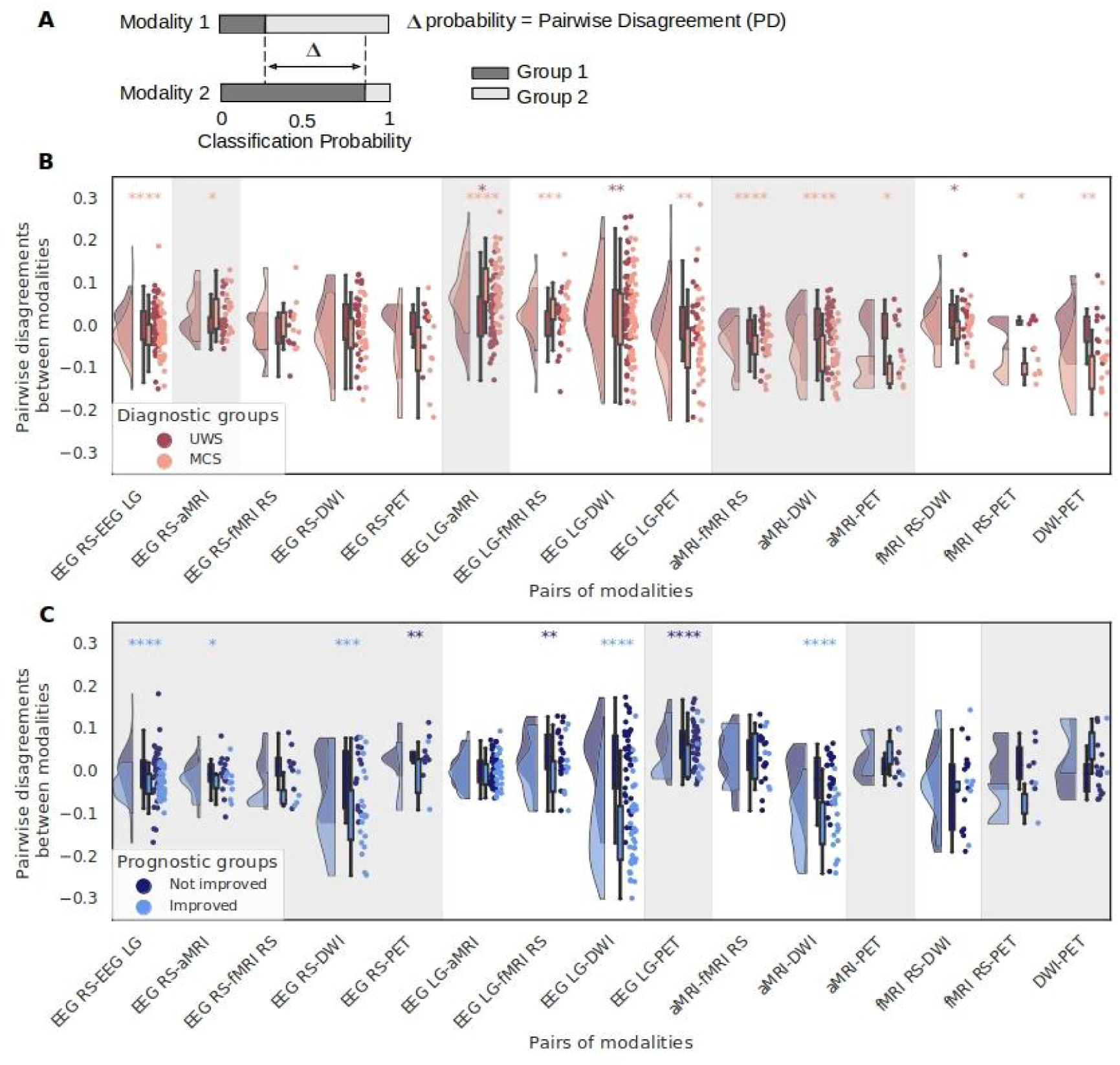
Pairwise disagreements (PD) of the classification probabilities are higher for patients in the Minimally Conscious State (MCS) and patients that show an improvement. (**A**) The pairwise disagreement is calculated per patient per pair of modalities as the absolute difference in the classification probabilities (probability of being in Group 1 vs Group 2, either for the diagnostic or the prognostic groups) between two modalities. (**B**) PD of the classification probabilities are more common in MCS patients. Results are displayed separately for the two diagnostic groups (UWS - dark red and MCS - light red). (**C**) PD are higher in improved patients than in not improved patients with a difference statistically less strong than the one of the diagnostic groups. PD of the classification probabilities are more common in improved patients. Results are displayed separately for the two diagnostic groups (Not improved - dark blue and Improved - light blue). The distributions of the PD are tested using a Wilcoxon signed-rank test to see if two paired samples are from the same distribution. The stars above the distributions denote the significance in the color related to the diagnostic group (* p<0.05; ** p<0.01; *** p<0.001; **** p<0.0001). The PD that have a gray background are those that include a model that was at chance level in Figure 2 A or B. Abbreviations: Unresponsive Wakefulness Syndrome (UWS), Minimally Conscious State (MCS), Electroencephalography (EEG), Resting State (RS), Local Global (LG) paradigm, functional Magnetic Resonance Imaging (fMRI), anatomical Magnetic Resonance Imaging (aMRI), diffusion Magnetic Resonance Imaging (dMRI), Positron Emission Tomography (PET).

#### Multimodal models

The multimodal model is a Late Fusion Multimodal (LFM) model that uses a stacking approach. In the first level, we have the unimodal RFC. The probabilistic classification outputs of these models (also called Basic models later on) are then given as features to the second layer where an ensemble RFC makes the final classification into the diagnostic or prognostic categories.

We also explored another version by adding demographic data and information on the patients’ etiologies using a one-hot-encoding approach alongside the results of the neural models, we call these Extended models. We add the value of -1 as a data imputation method when there is either a missing modality or a missing demographic or etiological information (the second was the case for only four patients who were admitted to the hospital for the evaluation in the period between 2010 and 2012, three were missing information on age and sex and the fourth one was missing information on the sex). The included demographic data is sex (female or male), and age at the time of the assessment. The additional etiological data we used is whether they are acute patients (within 90 days post-injury) or chronic, and four etiological categories: anoxic, traumatic brain injury (TBI), stroke, and other which also includes a few patients with mixed etiologies (N=14 for diagnosis from which 12 had TBI and anoxia, N=7 for prognosis out of which 6 had a TBI and anoxia).

Similarly to the unimodal models, we ran 100 initializations of five-fold stratified cross-validation, keeping the predictions from the fold that is used for testing. We then took either the average of the probability estimates per patient, or thresholded it to 1 when the value was above 0.5 and to 0 if the value was below 0.5. We then calculated summary metrics such as the balanced accuracy per number of modalities (Figure 5).

We analyzed the results based on the number of modalities each patient has, ranging from one to five. For each number of modalities (from one to five), and for each CV split (we have 500 splits because we ran 100 iterations each consisting of the 5-fold CV split), we calculate the balanced accuracies for the leave-out folds, and this gives us the distributions whose medians and interquartile ranges are given in Figure 5. Because the number of patients that have, for example, five modalities is smaller, in some of the leave-out CV splits there was not one of these patients present, thus the number of observations in this distribution is smaller (eg. 301 for diagnosis). Thus the number of observations reported in the Spearman correlation test results is the number of times a group of patients with a given number of modalities gets randomly selected in a leave-out fold.

### Statistical analysis

We used non-parametric statistical tests such as the Mann-Whitney U test, Bonferroni corrected with a significance level of 0.05 when comparing distributions of the basic and extended models per number of modalities. We used the Wilcoxon signed-rank test to test whether the median of PD distributions significantly differed from zero. We also performed Spearman correlation tests for the multimodal balanced accuracies plotted against the number of modalities, together with the 95% confidence intervals.

## Results

### Neuroimaging modalities carry independent diagnostic and prognostic information

In the diagnostic classification (Figure 2 A), the highest balanced accuracy was observed for PET (0.73 ± 0.13), followed by dMRI (0.69 ± 0.08), EEG LG (0.69 ± 0.06), EEG RS (0.66 ± 0.09), fMRI RS (0.63 ± 0.15), and the lowest for aMRI (0.51 ± 0.1). For the prognostic classification (Figure 2B), the highest balanced accuracy was achieved by dMRI (0.74 ± 0.09), followed by fMRI RS (0.63 ± 0.18), aMRI (0.58 ± 0.11), EEG LG (0.55 ± 0.06), PET (0.5 ± 0.16), and the lowest for EEG RS (0.49 ± 0.1). For both diagnostic and prognostic classification, we calculated the quality of the difference between the model distributions and the surrogates distributions (Supplementary Figure 2).

EEG recordings, whether RS or LG, exhibit a classification accuracy close to 0.7 for diagnosis but drop close to the chance level for prognosis. Conversely, aMRI shows an increase from chance level for diagnosis to 0.58 for prognosis, while PET exhibits the opposite trend, becoming non-informative in more of the CV splits for prognosis (in other words, one part of the distribution of the PET prognostic results is at chance level). dMRI and fMRI RS remain relevant for both diagnosis and prognosis, with dMRI gaining 0.05 points for prognosis.

Using two independent datasets (Dataset 2 from Germany and Dataset 3 from Italy), we examined the generalization of diagnostic prediction in four modalities. The balanced accuracy aligned with the training set for fMRI RS (0.7 +/- 0.12) and EEG LG (0.62 +/- 0.03) in Dataset 2 and for EEG LG (0.75 +/- 0.05), EEG RS (0.64 +/- 0.04), and aMRI (0.58 +/- 0.12) in Dataset 3 (Italy). The aMRI models for diagnosis for Dataset 1 are at chance level, which implies that we cannot test for their generalization. The models did not generalize above chance level only to fMRI RS (0.44 +/- 0.08 median balanced accuracy) in Dataset 3 (Italy), and to EEG RS (0.49 +/- 0.02 median balanced accuracy) in Dataset 2.

### Pairwise disagreements of unimodal models are higher in MCS and improved patients

When examining unimodal predictions per patient, we observed a discernible difference in the extent of disagreements across patient groups. Notably, PD are markedly higher in patients classified as being in an MCS compared to those in a UWS (Figure 3 B). The trend in the diagnosis is primarily driven by combinations involving aMRI, PET, fMRI, and EEG LG, leaving EEG RS to be more similar to the other modalities. A similar trend, though less pronounced, is evident when comparing patients who show improvement versus those who do not (Figure 3 C) where modalities contributing significantly to these disagreements are dMRI and the EEG paradigms.

### Feature importance differs in diagnostic and prognostic models

The fluctuations in diagnostic and prognostic accuracy across individual modalities require an exploration into the most influential features for prediction, examining whether these vary when assessing the patient’s current state or their outcome. To investigate this, we ranked the features per modality based on their average importance scores across all model iterations. Figure 4 A, D, and F depict the FI scores for fMRI, PET, and aMRI showcasing cortical and subcortical regions. Whereas Figure 4 B, C, and E show the mean FI combined in groups for the EEG RS and LG and the dMRI scans. The diagnostic aMRI models, together with the prognostic EEG RS and PET models, are all at chance level, thus we will not be analyzing their FI scores.

**Figure 4.**
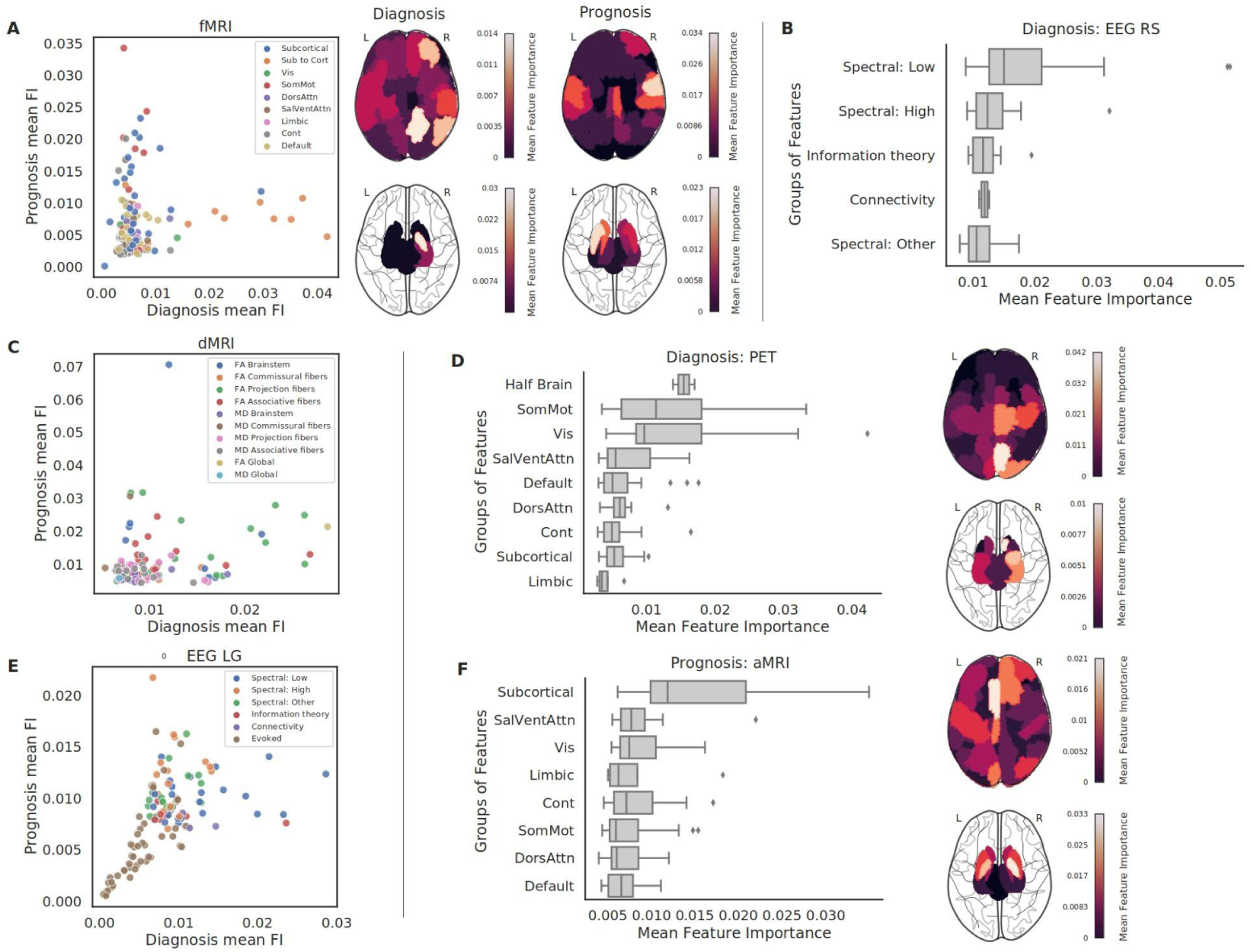
Reordering of the feature importance (FI) distributions per group of features for diagnosis and prognosis. (**A**) FI of the diagnostic and prognostic prediction using the fMRI RS scans. The swarm plot shows the FI split into within subcortical functional connectivity, within cortical connectivity subdivided into the 7 cortical networks, and subcortical to cortical functional connectivity. The brain plots show the FI of the within cortical and within subcortical functional connectivity per region of interest. (**B**) FI of the diagnostic prediction using the EEG RS recordings. The bar plot shows the FI split into conceptual families: connectivity (wSMI), information theory (Kolmogorov complexity and permutation entropy), low spectral (delta, theta, and alpha frequency bands), high spectral (beta and gamma bands), and other spectral marker summaries. (**C**) FI of the diagnostic and prognostic prediction using the dMRI scan. The swarm plot shows the FI split into two measures Fractional Anisotropy (FA) and Mean Diffusivity (MD) which are subdivided into global brain-wide measures and families of tracts: projection fibers, brainstem, commissural fibers, and associative fibers. (**D**) FI of the diagnostic prediction using the FDG PET scan. On the brain plots the FI of the metabolic activity per cortical or subcortical region of interest are shown. The bar plot shows the FI split into cortical networks and the subcortical regions, as well as the importance of the half-brain (left or right hemisphere) metabolic activity. (**E**) FI of the diagnostic and prognostic prediction using the LG task-based EEG recordings. The bar plot shows the FI split into the same conceptual families as the EEG RS with the addition of the evoked markers coming from the task-based paradigm. (**F**) FI of the prognostic prediction using the aMRI scan. On the brain plots the FI of the cortical thickness and the subcortical volume are shown. All of the bar plots, including the brain plots from four different views are given in Supplementary Figures 10, 11, 12. Abbreviations: Default Mode Network (Default), Dorsal Attention Network (DorsAttn), Salience/Ventral Attention Network (SalVentAttn), Somato-Motor Network (SomMot), Visual Network (Vis), Limbic Network (Limbic), Control or Frontoparietal Network (Cont), Electroencephalography (EEG), Resting State (RS), Local Global (LG) paradigm, functional Magnetic Resonance Imaging (fMRI), anatomical Magnetic Resonance Imaging (aMRI), diffusion Magnetic Resonance Imaging (dMRI), Positron Emission Tomography (PET).

Examining spectral subcategories of low-bands (delta, theta, alpha) and high-bands (beta and gamma), we found that, in both paradigms, for diagnosis, low-frequency-based features are most relevant (Figure 4 B, E and Supplementary Figure 10 G, I). In diagnostic EEG LG, connectivity-derived features are also highly relevant, while high-frequency-based features are less informative. Conversely, for EEG LG prognostic prediction (Figure 4 E and Supplementary FIgure 10 J), high-frequency features become the most informative. Whereas the evoked features remain the least important for both diagnosis and prognosis. In the aMRI prognostic models, the most relevant features are the subcortical volume features followed by the salience and visual network cortical thicknesses, with the features from the DMN being the lowest scoring. In fMRI RS functional connectivity analysis, subcortical regions to cortical networks connectivity features are most informative for diagnostic classification, followed by within-subcortical and cortical-to-cortical regions of interest connectivities (Figure 4 A). Conversely, for prognosis (Figure 4 A), the somatomotor cortical network gains importance, along with increased relevance of the limbic and the frontoparietal network. The visual and salience networks and the subcortical to cortical functional connectivity decrease in significance. In diagnostic FDG PET analysis (Figure 4 D), mean metabolic activity per left or right hemisphere emerges as the most informative feature, followed by cortical networks like somatomotor and visual, while subcortical areas are less informative. In dMRI, the most important feature distinguishing UWS and MCS patients is the combined fractional anisotropy (FA global) (Figure 4 C). This is followed by the right superior fronto-occipital fasciculus, right posterior limb of the internal capsule, and left and right corona radiata. When grouping tracts, projection fibers and brainstem fibers are most informative for diagnosis based on fractional anisotropy (Figure 4 C). For prognosis, the mean diffusivity of commissural fibers rises in importance (Figure 4 C), and the brainstem tracts measured by FA remain among the most informative.

In addition, we also calculated the univariate AUC values per feature (Supplementary Figures 3 to 8), both for diagnosis and prognosis. We also show a non-linear relationship between the mean feature importance scores and the feature AUC values (Supplementary Figure 13).

### Multimodal integration improves predictive accuracy

In this section we address two questions 1) whether the model accuracy improves with an increase in the number of modalities, and 2) whether extended models will perform better than basic ones.

In the case of diagnostic prediction (Figure 5 A), we observed an increase in balanced accuracy for both basic and extended models. The basic model started at a chance level and progressively improved to achieve an accuracy above 0.83. For prognosis (Figure 5 B), there was an upward trend in accuracy, with a notable deviation when patients had four modalities, leading to a drop in accuracy, particularly for the basic model. In most cases, the extended model demonstrated superior performance, indicating non-redundant information derived from demographic details and etiological divisions.

**Figure 5.**
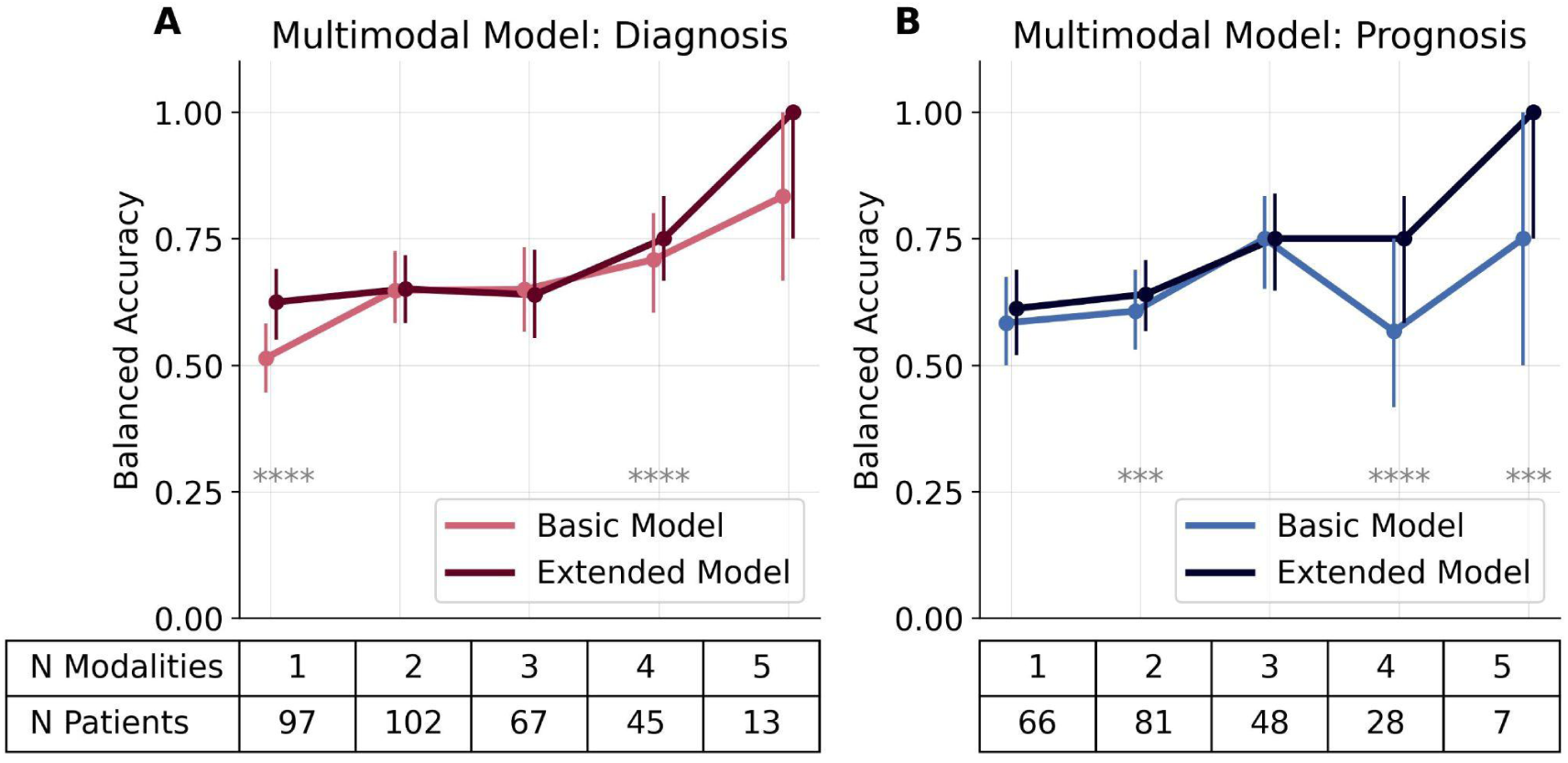
Increasing trends in the multimodal model balanced accuracy for the Basic models (only neural modalities) and the Extended models (neural modalities plus information on the patient etiologies and their demographics). (A) Balanced accuracy for the diagnostic models (basic and extended) for patients that have from 1 to 5 different neuroimaging modalities (x-axis). (B) Same as (A) but for the prognosis. The error bars represent the first (Q1) and third (Q3) quartile intervals of the distributions. On the x-axis, the first row represents the number of modalities across the function, and the second row is the number of patients that have the given number of modalities. The stars represent significance following Mann Whitney U tests (Bonferroni corrected) between the basic and extended models (* p<0.05; ** p<0.01; *** p<0.001; **** p<0.0001). Abbreviations: number of elements in the given distribution (N).

When looking into the trends for the models using a Spearman correlation test, for the diagnostic models we see an increase of balanced accuracy for the basic model r(2294)=0.49, p<0.0001, [95% CI: 0.46 - 0.52]; and for the extended model r(2294)=0.399, p<0.0001, [95% CI: 0.36 - 0.43]. For the prognosis, the positive correlation of the balanced accuracy with the number of modalities is less strong r(2037)=0.105, p<0.0001, [95% CI: 0.06 - 0.15], and increases for the extended model to r(2037)=0.335, p<0.0001, [95% CI: 0.3 - 0.37].

The statistical difference between the basic and extended model in diagnosis was for n=1 modalities (U(500,500)=55562, p<0.0001), and n=4 (U(495,495)=98743, p<0.0001), whereas in prognosis was for n=2 (U(500,500)=107557, p=0.0007), n=4 (U(465,465)=68766, p<0.0001), and n=5 (U(74,74)=1748.5, p=0.0002) modalities. For the diagnosis for n=2 (U(500,500)=124698, p=1), n=3 (U(500,500)=130672, p=1), and n=5 (U(301,301)=42645, p=0.92) the basic and extended model results are not statistically different, the same is true for the n=1 (U(500,500)=116245, p=0.28), and n=3 (U(500,500)=124105, p=1) for the prognosis.

## Discussion

### Interpretable modeling approach for sparse multimodal neuroimaging datasets

The assessment of DoC presents significant clinical challenges due to the dissociation between behavioral responsiveness and consciousness, as well as the limitations of behavioral scales in the presence of confounding factors. While individual neuroimaging and electrophysiological modalities have shown potential in improving diagnostic and prognostic accuracy, they are inherently limited in scope. Multimodal approaches, which integrate information across various modalities, hold promise for addressing these limitations, but there has been a lack of large-scale, multicentric studies systematically evaluating their complementarity and effectiveness. In this study, we adopt a comprehensive, multicentric approach to overcome these challenges. We integrate six neuroimaging modalities using interpretable ML methods. To handle the inherent challenges of sparse and heterogeneous multimodal datasets, we propose a two-model stacking approach. This methodology effectively addresses issues such as the low patient-to-feature ratio, modality-specific data sparsity, and the heterogeneity of modality combinations, enabling a robust analysis of both diagnostic and prognostic dimensions of DoC.

### Differentiating unimodal classification scores for the current state of the patients and their evolution

Our analysis of neuroimaging modalities for patients with DoC revealed intriguing modality differences in diagnostic and prognostic accuracy. Modalities that capture the structural preservation of the brain and its networks (aMRI and dMRI), become more relevant for the evolution of the patients. Whereas modalities that capture electrical activity (EEG) and metabolic activity (PET) are mostly relevant for diagnosis. This shift in accuracy rankings for modalities between diagnostic and prognostic classifications underscores their complementarity.

PET had the highest accuracy in discriminating between UWS and MCS patients but seems to carry little prognostic information in half of the CV splits (Figure 2 A and B). The diagnostic results are aligned with previous studies that show that metabolic data distinguishes MCS from UWS patients^35,41,45,46^, with one study showing that the gradient increase continues to EMCS patients and healthy controls^36^. Furthermore, FDG-PET has demonstrated better suitability in discriminating DoC diagnoses compared to MRI-derived measures, both active fMRI (where PET had higher sensitivity for identifying MCS patients)^35^, aMRI and RS-fMRI^46^. In our work, we observed a drop in the prognostic accuracy of PET compared to the diagnostic one, however, the distribution is large (Figure 2 A and B) which indicates that certain CV splits are better predicted whereas other splits show opposing trends to their training subset. This large variation in the accuracy could arise due to the big imbalance in patients that improve (n=9) versus those that do not (n=35) (Table 1). One study from the literature also shows a drop in the patient recovery value of PET compared to the diagnostic, albeit still as high as 74% ^35^, whereas another study, in a sample of 20 patients, could not show a prognostic value^47^. Comparing the PET with the EEG models, one study showed a higher sensitivity of EEG models compared to FDG-PET, although the AUC of the diagnostic prediction did not differ significantly.^41^

In our study, we used the same RS and LG paradigm EEG markers as those reported in two previous studies.^18,27^ There is a partial overlap in the data with the previous works and the results are in accordance with our study. Additionally, we summarize the markers into large groups and contrast their diagnostic and prognostic accuracy. We observe a drop in the accuracy of both EEG paradigms for the evolution of the patients (Figure 2 A and B) (in contrast to the higher accuracy in the diagnostic models), as previously shown in other studies^24,48^. On the contrary, other studies were able to show both the diagnostic and prognostic potential of EEG. ^23,39,40,49^

In our work, we observe a diagnostic accuracy of aMRI close to chance levels (Figure 2A). One study found similar diagnostic results (balanced accuracy ranging from 0.45 to 0.63)^31^. However, Annen et al.^32^ show a high diagnostic area under the receiver operating curve of 96% using gray matter and white matter volume, a prediction comparable to that of FDG-PET. Importantly, we used a different implementation of the cortical thickness and subcortical volume estimates that is created specifically for clinical data with various resolutions and originating from different neuroimaging centers^50–53^. The differences in the findings compared to Annen et al.^32^ could be arising due to the different methodology and diverse cohorts, leading to questions that should be answered in separate studies comparing both FreeSurfer implementations across different etiologies (for example traumatic versus anoxic). In comparison to the diagnostic models, we see an increase in accuracy when looking into the prognosis (Figure 2, A and B). When there is physical damage to the tissue that can be quantified with neuroimaging, the regeneration is slow and this could dictate the prognosis of the patients. In other words, patients who have more damage are less likely to improve, but the size of the damage is not directly related to their current state as there can be a substantial lesion in an area that does not directly affect the behaviors we test with the CRS-R.

For the fMRI RS modality, we see a similar median balanced accuracy for the diagnosis and prognosis but a larger distribution for the prognostic prediction, similar to the results of PET, suggesting the influence of the results depending on the random CV splits. However, the ordering of the modalities’ performance is different, which makes the fMRI RS one of the most informative modalities for the patients’ evolution. Previous work has shown fMRI RS potential of discriminating between patients and controls^17^, and between UWS and MCS patients^21,29,39^. The higher accuracy reported in the literature compared to our results could come from the methodological difference of seed-based versus atlas-based parcellation of RS networks or the fact that in one of the studies^21^ they used data only from patients for whom the clinical diagnosis based on CRS-R was congruent with PET scans. Two studies have tested the outcome prediction of DoC patients at three months with an accuracy range of 0.69-0.78^40^ and 0.81^22^, but no predictive value at 12 months was observed^40^. Given that we look at a different prognostic metric, the results are not directly comparable, but there is consistent evidence that fMRI activity does contain prognostically relevant information for DoC patients.

Previous diagnostic studies using dMRI have reported accuracies as high as 0.95^54^ and in a range of 0.81-0.84 using a multivariate searchlight analysis of whole brain thalamo-cortical tracts^55^. In prognostic studies of cardiac arrest patients, fractional anisotropy values have been shown to reach values of 0.95 sensitivity, 1 specificity^56^, 0.98 AUC in a larger follow-up study^28^, and 0.93 AUC 1-year prognostic value of global deep white matter metrics in TBI patients^57^. The consistency of our results with previously reported results emphasizes the importance and potential of using dMRI to aid in the diagnostic and prognostic assessment of patients in a DoC.

Generalization tests across independent datasets demonstrated varying performance (Figure 2 C and D). The modalities that could not be generalized include the fMRI RS from Dataset 3. This discrepancy may be attributed to the heterogeneity in acquisition parameters compared to the training set (see Methods). Task EEG (LG) outperforms resting state EEG across all centers, with a notably reduced effect in Dataset 2 from Germany. The higher performance of EEG LG-based models highlights the critical role of active paradigms^1^ in assessing patients with DoC, as these paradigms likely enhance and regulate patients’ attentional states. In contrast, resting-state paradigms may be less robust to cross-center variability due to their dependence on intrinsic brain activity, which is more susceptible to external and patient-specific factors. These findings together underscore the importance of accounting for modality-specific and center-related acquisition parameters to improve model generalizability across centers.

### Pairwise disagreements between modalities

Studying the PD across modalities is important as it can point toward cases of patients where a dissociation can elucidate the potential of recovery. We observed more PD in MCS patients and those that improve (Figure 3), implying that the more positive clinical picture can be captured by some signals and not others. The sources of these disagreements can be neural or non-neural. Neural examples include the case of a functional hemispherectomy when a patient showed almost no metabolic activity in the left hemisphere with preserved white matter tracts^58^ or islands of preserved cortical activity that are posited to exist in this group of patients^59^. Furthermore, UWS patients with unfavorable EEG features have shown an increase in fMRI between-network connectivity and DMN within-network connectivity decrease (but not significant).^39^ Although EEG and PET have shown to be highly correlated, EEG connectivity patterns differed in PET-negative and PET-positive patients.^24^ Another study found a diagnostic difference (between healthy controls and DoC patients) in metabolic activity and mixed results in positive and negative DMN connectivity, but no significant results in gray matter volume^36^. A disagreement between metabolic activity and gray matter has been found in the left-sided language network of patients in MCS- and MCS+.^37^ The first exhibited lower metabolic values in the left middle temporal cortex and a metabolic functional disconnection between the left angular gyrus and the left prefrontal cortex. The authors conclude that brain function and not gray matter structure supports the clinical signs of language processing.^37^ In some patients, only the dMRI images showed a consistent loss of white matter compared to the seemingly unchanged appearance of structural images.^60^ Furthermore, the reliability of discriminating between MCS and UWS patients is often compromised due to the limited sensitivity of scalp EEG, as demonstrated by instances where pathological brain activity can mask normal neuronal patterns in awake individuals, suggesting the potential for complex dissociations in severe brain injury cases^10^. The proposal to rename the MCS to a cortically mediated state^61^ underlines the fact that the MCS encompasses a broad and heterogeneous range of conditions. This spectrum includes both unconscious patients who exhibit residual cortical activity leading to observable behavior and conscious patients who, despite possibly being self-aware, are hindered by executive deficits, which prevent them from effectively using a communication code or responding functionally to commands.^61,62^ This may explain why there are more PD in MCS compared to UWS, and that some specific modalities might not capture the heterogeneity. A complementary perspective is that the differences in the prediction can also come from etiology-specificities such as EEG alpha power that has been shown to be suppressed in severe post-anoxic patients and does not differ in patient groups having other etiologies^30,63^. All these examples corroborate the fact that PD from neural origin are common, and their hierarchical importance in diagnosis and prognosis needs to be further studied.

One non-neural source of disagreement is the data quality which, even with stringent exclusion criteria, is lower in this patient group, and it can affect analyses down the line. Furthermore, the state of the patients fluctuates across various time scales, whereas in our case we work with one diagnosis per patient, which could be the source of disagreement in diagnostic models, which would not be the case for the prognostic prediction. A way to surpass this is to look at the variability of multiple CRS-R tests and check the disagreement in light of the patient’s clinical fluctuations.

### Differing importance of feature groups within modalities, for diagnostic and prognostic prediction EEG

Previous results showed that the most informative EEG features to differentiate between UWS and MCS patients are absolute alpha power, permutation entropy, Kolmogorov complexity, and a connectivity measure in the theta band (wSMI).^18,27^ In our work, we find that in EEG, the spectral feature groups are the most informative (Figure 4 B, E and Supplementary Figure 10 G, I, J), with the low frequencies being important for diagnosis and the high frequencies for prognosis. This finding can be related to the mesocircuit hypothesis.^7,64^ The importance of high frequencies for the prognosis in the DoC patients that we report could further help in distinguishing the MCS patients that belong in group C of the ‘ABCD’ model of corticothalamic dynamics as having the potential to behaviorally improve.

In prognostic EEG studies of prolonged DoC, the presence of dominant delta frequencies and reduced EEG amplitudes was related to worse outcomes, whereas the dominance of alpha frequencies, preserved EEG reactivity, and an increase of the dominant frequency were associated with improvement.^10,23^ On the contrary, in one study, higher delta power has been associated with improved outcomes for patients transitioning from UWS to MCS^47^ which we also find in EEG LG (Supplementary Figure 4). These discrepancies can come from etiology-dependent differences such as the slowing of EEG being relevant for prognosis in toxic encephalopathies, a transient increase in slow waves or suppression of sensory stimuli in patients with a traumatic brain injury, contrary to the increase in gamma and alpha frequencies in acute patients with a subarachnoid hemorrhage^10^.

In our work, we see that connectivity metrics have a stronger weight in the LG acquisition compared to RS (Figure 4 B, E and Supplementary Figures 3 and 4). Previous work has shown the importance of network metrics over frequency power^24^, the relevance of coherence across various regions and frequency bands for improvement of UWS patients^23^, and a stronger delta network connectivity in patients with negative outcomes^24^. On the contrary, a study found no network features related to outcome at three or six months post-injury^48^, with only relative alpha power improving prediction accuracy at three months in contrast to predicting using only clinical features. This is consistent with our findings, where univariate AUC values are at chance level for the prognosis, both in EEG RS and LG.

When using oddball auditory perception paradigms, the event-related potentials such as mismatch negativity (MMN) and the P300 have been reported to have a low sensitivity in MCS patients.^10,65^ Accordingly, in our work, most of the evoked features are the least informative ones (Figure 4 E).

#### Neuroimaging

Multiple studies using PET or fMRI RS have reported a brain-wide network difference between UWS and MCS patients^21,41,45^, with some reporting that the left hemisphere is more impaired in UWS^31,46^. In our work, in most neuroimaging modalities, we observe a distributed brain-wide FI (Figure 4, A, D, F, brain plots), with the exception of the fMRI RS prognostic FI.

However, the DMN has been mostly put into focus to differ in diagnostic groups, to have lower activity in UWS compared to MCS^15,17,22,46^, both at enrollment and at ICU discharge^39^ and to relate with recovery outcome^20,22,25,26,46^. Structural information such as gray matter volume FI^32^ and structural integrity^31^ have also been reported to be highest in DMN regions. In our prognostic results, the DMN features are moderately informative compared to the other networks in the aMRI and fMRI scans (Figure 4 A, F and Supplementary Figure 10 B, D). In the fMRI scans, where both diagnostic and prognostic models are comparable, they are more informative for the prognosis of the patients.

A few studies report the strongest metabolic activity reduction in frontoparietal areas^45,46,66^, or specifically in the medial prefrontal cortex (part of DMN) or lateral parietal cortex^22^. Specific networks that have been associated with diagnosis, are the primary and associative somatosensory areas^45^ with one FDG-PET study observing this in the best-preserved hemisphere^62^. A few studies report specific diagnostic differences only for the auditory network in fMRI RS^17,21^ and glucose metabolism to be higher in MCS than in UWS^47^. In PET and fMRI RS, we also observe the higher relevance of somatomotor and visual networks. In contrast, one study reports that higher-order networks (DMN, salience, dorsal attention network, left and right fronto-parietal network, and temporal network) had better diagnostic accuracy than low-order networks (sensorimotor, auditory, and visual networks) as derived by their structural integrity^31^, but we cannot compare this to our results because the aMRI model is at chance level.

In our work, we find that the subcortical areas become more important for prognosis (in contrast to diagnosis), however, this difference has not been the focus of neuroimaging investigations. Various subcortical areas have been reported to be different in diagnostic groups such as a lower metabolic activity in the brainstem^32,45^, thalamus^32,45^, the caudate and para-hippocampal areas^32^. Thalamic white matter integrity has also been affected in patients^54^, along with the pathway linking the posterior cingulate cortex/precuneus with the thalamus, as evidenced by their mean fractional anisotropy values^67^. Whereas brainstem white matter tract preservation was only observed in the ischaemic-hypoxic etiological group^60^, and no mean diffusivity brainstem differences between MCS and UWS^54^.

### Multimodal combinations

The increase of the balanced accuracy of patients that have multiple different modalities can be expected from literature either for the combination of EEG and FDG-PET^41^, EEG and fMRI RS^39,40^, EEG and dMRI^42^, aligning with our results. Similar studies in cardiac arrest patients show that a model utilizing only three FA features outperformed models incorporating either only the FA global scores, clinical data, or gray matter apparent diffusion coefficient^56^, or an enhanced AUC with the integration of scores and metrics from multiple modalities (EEG, aMRI, dMRI)^28^. A case study by Comanducci et al.^11^ illustrates how longitudinal multimodal analysis can reveal covert signs of consciousness in an unresponsive patient. Additionally, Rohaut et al.^33^ demonstrate that integrating multimodal observations enhances neuro-prognostication performance.

### Benefits, caveats, and the future of integrative multimodal neuroimaging for DoC

The current electrophysiology and neuroimaging modalities capture some aspect of brain preservation - either the underlying structure or dynamics, and they contain non-redundant information. The importance of having a multidimensional perspective on this clinical group has been increasingly emphasized.^4,13,33,39–41^ Overall, numerous reviews put focus on the advantage of having multimodal acquisitions^4,7,12,13,68^, however, the practical combination given the limitations already exposed in this paper makes the implementation a challenging one. Furthermore, ML-based approaches, trained on behavioral labels to differentiate between UWS and MCS patients, may overlook conscious but unresponsive individuals, posing a circularity problem; however, there is some robustness to mislabeling if classifiers are trained with a sufficient amount of data^27^. Importantly, RFC are non-linear models, thus the relationship between the FI and the models (measured here through the balanced accuracies) is non-linear. Although the FI can be very informative to understanding how decisions are made, they have important limitations, and further analysis of redundancy and synergy can paint a clearer image of their relationships. Furthermore, future investigations should focus on the distinction between different etiologies of patients with DoC using combined multimodal approaches (due to interactions of neural signals with etiology^30,63^.

## Conclusions

In this study, we developed an explainable ML approach for the classification of DoC patients from a large dataset of multimodal neuroimaging recordings. The observed distinctions in accuracy, feature importance, and pairwise disagreements, underscore the need for tailored strategies in leveraging neuroimaging modalities for enhanced clinical decision-making. The comparison of the current state of the patients and their evolution across modalities and features (regions or other signal summaries), may open the path to more careful investigations within etiologies or integrative neuroimaging studies that have more narrow hypotheses.

## Data availability

The data is not publicly available. Codes used in the analysis will be made publicly available following the publication.

## Data Availability

The data is not publicly available.

## Acknowledgements

We thank all the participants who took part in the studies. We would like to thank the work and support of the clinicians at the Neuro ICU, DMU Neurosciences, APHP- Sorbonne Université, Hôpital de la Pitié Salpêtrière, Paris, France; University Hospital of the Ludwig-Maximilians-University of Munich; Therapiezentrum Burgau; the University of Milan; Fondazione Don Carlo Gnocchi Santa Maria Nascente; the Weizmann Institute of Science, and the patient families whose consent and understanding are essential to the progress of the field.

## Supplementary material

The supplementary materials are available in a separate pdf file.

