## Supplementary Materials for "Integrative Electrophysiology and Neuroimaging Approach in Assessing Disorders of Consciousness: A Multimodal Multicentric Machine Learning Study"

Paris, France
[11] Medical intensive care unit, HEGP Hôpital, Assistance Publique - Hôpitaux de Paris-Centre (APHP-Centre), Paris, France
[12] Centre Mémoire de Ressources et de Recherche, Paris Nord/Université Paris-Cité
[13] Center for Brain and Cognition, Computational Neuroscience Group, Universitat Pompeu Fabra, Barcelona, Spain
[14] Nuclear Medicine, Faculty of Medicine, University of Augsburg, Stenglinstr. 2, 86156 Augsburg, Germany
[15] Diagnostic and interventional Neuroradiology, Faculty of Medicine, University of Augsburg, Stenglinstrasse 2, 86156 Augsburg, Germany
[16] BRAINTALE SAS, Paris, France
[17] Institució Catalana de la Recerca i Estudis Avançats (ICREA) Barcelona, Spain
[18] Latin American Brain Health Institute (BrainLat), Universidad Adolfo Ibáñez, Santiago, Chile
[19] Sorbonne University, GRC 29, AP-HP, DMU DREAM, Department of Anaesthesiology and Critical Care Medicine, AP–HP, Pitié-Salpêtrière Hospital, 47-83, boulevard de l'Hôpital, 75013, Paris, France
[20] AP-HP, Hôpital de la Pitié Salpêtrière, Neuro ICU, DMU Neurosciences, Paris, France
[21] AP-HP, Hôpital Pitié - Salpêtrière, Service de Neurophysiologie Clinique, Paris, France
[22] Department of Medical Neurobiology, Institute for Medical Research Israel Canada, Faculty of Medicine, The Hebrew University of Jerusalem, Jerusalem, Israel
[23] Department of Cognitive and Brain Sciences, The Hebrew University of Jerusalem, Jerusalem, Israel
[24] Department of Neurorehabilitation, Medical Faculty, University of Augsburg, Augsburg, Germany

**This PDF file includes:**

Supplementary Methods

Supplementary Results

Supplementary Figures 1 to 14

Supplementary Tables 1 to 7

Please note that some of the figures may appear small, but they are in vector format, so readers can zoom in without losing quality.

### Methods

#### Modalities

##### Electroencephalography (EEG)

Acquisitions

In Dataset 1, the EEG recordings were collected using a high density 256-electrode (EGI300) Geodesic Sensor Net (Electrical Geodesics, Inc. (EGI) system, Eugene, Oregon, USA) referenced to the vertex, at a sampling frequency of 250 Hz; electrode quality standards required impedance to be below 75 kΩ. In Dataset 2, the EEG recordings were collected using two systems: i) a 256-electrode (EGI400) Geodesic Sensor Net (Electrical Geodesics, Inc. (EGI) system, Eugene, Oregon, USA) referenced to the vertex, at a sampling frequency of 250 Hz in the center in Burgau (Germany) with electrode quality standards requiring impedance to be below 50 kΩ; ii) a 64-channel BrainVision EEG system (62 electrodes + 2 EOG channels) in the center in Munich (Germany); with electrode quality standards requiring impedance to be below 10 kΩ. In Dataset 3, the EEG recordings were collected using a 64-electrode (60 electrodes + 2 EOG channels + 2 EKG channels) BrainVision system (Brain Products GmbH, Gilching, Germany) referenced to the forehead, at a sampling frequency of 1000 Hz; electrode quality standards required impedance to be below 10 kΩ.

Before the start of the recordings, the CRS-R arousal facilitation protocol was administered. In Dataset 1, first the LG auditory paradigm was administered followed by a five-minute RS, whereas in Datasets 2 and 3, first there was a ten-minute RS followed by the LG recording. The LG paradigm developed by Bekinschtein et al. (2009), involves an oddball paradigm with sequences of tones (XXXXX or XXXXY). During the acquisitions, patients were not anesthetized and the data was acquired during a time window when they were more likely to be awake (either in the morning or early afternoon). In the Paris center (Dataset 1), the arousal protocol from the CRS-R was administered between each LG block. In the German centers (Dataset 2), in case the vigilance dropped and patients did not open the eyes again spontaneously, the recording was stopped after the present block (for motor imagery or local global) or stopped immediately (for resting-state). The arousal protocol from the CRS-R was administered, and the recording was continued.

#### Preprocessing

The methodological details of the EEG acquisitions, preprocessing, and markers analysis are the same as previously reported (Sitt et al., 2014; Engemann et al., 2018). EEG data was preprocessed and analyzed using the MNE-Python (Gramfort et al., 2013) and NICE (Engemann et al., 2018) packages (Supplementary Materials). All three datasets were preprocessed using the same algorithms which are EEG montage agnostic. The only difference is the cutting of the RS acquisitions length to five minutes to match the length in the Dataset 1.

Electrodes exhibiting voltages exceeding 100 µV in more than 50% of the epochs were excluded from the analysis. Additionally, voltage variance was calculated across all remaining electrodes, and those with a variance Z-score greater than four were also excluded. This procedure was iterated four times. Subsequently, bad electrodes were interpolated using a spline method (Perrin et al., 1989). Epochs were marked as bad and discarded if the voltage exceeded 100 µV in more than 10% of the electrodes. Voltage variance was again computed across all valid epochs, and those with a Z-score above four were removed, repeating this process four times. An average reference was then applied to the remaining epochs. Pre-processing was performed using the MNE-Python package.

In the Local-Global (LG) paradigm, trials were band-pass filtered between 0.5−45 Hz and segmented into epochs ranging from −200 ms to +1344 ms relative to the onset of the first sound. Non-scalp EEG electrodes were removed, resulting in 195 electrodes for subsequent preprocessing steps. The remaining stimulus-locked epochs were averaged and transformed to an average reference. A 200 ms baseline correction (before the onset of the fifth sound) was applied.

#### Markers

Normalized and raw power spectral analysis is applied to estimate power in five frequency bands. Markers of complexity include Permutation Entropy (PE) and Kolmogorov Chaitin complexity (KS), utilizing gzip compression for signal redundancy assessment. Markers of connectivity involve the weighted symbolic mutual information (wSMI), evaluating long-distance connectivity. Further details on the markers rationale and computation can be found in the supplementary materials of the original publications (King et al., 2013; Sitt et al., 2014). To summarize the large distributions of features, we created feature groups. For the EEG RS, the feature groups are spectral, information theory, connectivity, and other spectral. The spectral features are further split in low-bands for features involving the frequency bands delta (1-4 Hz), theta (4-8 Hz), and alpha (8-13 Hz), and in high-bands for the bands beta (13-30 Hz) and gamma (30-45 Hz). The EEG LG has an additional group of evoked features. The feature names and their associated groups are given in Supplementary Table 6.

#### Magnetic Resonance Imaging (MRI)

##### Acquisitions

In Dataset 1, three different MRI modalities were used: anatomical MRI (aMRI) T1 weighted acquisitions, resting-state functional MRI (RS-fMRI) scans of around five minutes, and diffusion MRI (dMRI). Anatomical and functional MRI images were acquired using two distinct acquisition protocols. In the first protocol, 3T General Electric Signa system (GE HealthCare Technologies Inc., Chicago, IL, USA) was utilized to acquire T2*-weighted whole-brain RS images with a gradient-echo EPI sequence in axial orientation (200 volumes, 48 slices, slice thickness: 3 mm, TR/TE: 2400 ms/30 ms, voxel size: 3.4375×3.4375×3.4375 mm, flip angle: 90°, FOV: 220 mm2). Additionally, an anatomical volume was obtained using a T1-weighted MPRAGE sequence in the same acquisition session (154 slices, slice thickness: 1.2 mm, TR/TE: 7.112 ms/3.084 ms, voxel size: 1×1×1 mm, flip angle: 15°). T2*-weighted whole-brain resting-state images were recorded using a 3T Siemens Skyra System (Siemens Healthineers, Erlangen, Germany) with a gradient-echo EPI sequence in axial orientation (180 volumes, 62 slices, slice thickness: 2.5 mm, TR/TE: 2000 ms/30 ms, voxel size: 2×2×2 mm, flip angle: 90°, FOV: 240 mm2, multiband factor: 2). In the same session, an anatomical volume was obtained using a T1-weighted MPRAGE sequence (208 slices, slice thickness: 1.2 mm, TR/TE: 1800 ms/2.35 ms, voxel size: 0.85×0.85×0.85 mm, flip angle: 8°). The patients who were administered medication that could greatly affect their vigilance state such as (propofol and benzodiazepine medication), before the fMRI scans, were excluded only from the fMRI analysis.

The dMRI acquisition details are reported in previous publications (Velly et al., 2018; Altmayer et al., 2024), further details can be found in the Supplementary Materials.

In Dataset 2 (Germany), 3T Philips Ingenia system (Philips Healthcare, Best, Netherlands) was used to acquire T2*-weighted whole-brain resting-state images (172 volumes, TR/TE: 2400 ms / 33 ms, flip angle 90°, voxel size: 1.4583x1.4583x3 mm, FOVx 210 mm, FOVy 210 mm, 45 slices, slice thickness 3 mm). Additionally, an anatomical volume was obtained using a T1-weighted sequence either in the same session or from a different center. The acquisition parameters are given in the Supplementary materials (Supplementary Table 7).

In Dataset 3 (Italy), 3T Siemens Prisma system (Siemens Healthineers, Erlangen, Germany) was used to acquire multi-echo T2*-weighted whole-brain resting-state images (400 volumes, TR 1000ms, TE: 15, 33.7, 52.4 ms, voxel size: 2.7x2.7x3 mm, flip angle 58°, FOVx 260 mm, FOVy 260 mm, 48 slices, slice thickness 2.5mm, gap 0.5mm). Additionally, an anatomical volume was obtained using a T1-weighted MPRAGE sequence in the same acquisition session (320 or 256 slices, slice thickness: 0.8 or 1 mm, TR/TE: 2300 ms / 3.1 ms, voxel size: 0.8×0.8×0.8 mm or 1x1x1 mm, flip angle: 8°).

#### Preprocessing

For the anatomical MRI scans, we used the FreeSurfer clinical software (v.7.4.1) (Iglesias et al., 2023) with the recon-all-clinical implementation (Gopinath et al., 2023 (preprint); Billot et al., 2023a; Billot et al., 2023b; Iglesias et al., 2023) to obtain the surface reconstruction and the extraction of whole brain and volumetric features per cortical and subcortical regions of interest (ROI). The clinical version was chosen due to the varying acquisition parameters within the long period of the study and the inclusion of the data from the two other centers in Germany and Italy. We excluded scans that did not pass the segmentation or normalization steps (either due to movement artifacts or lesions). The anatomical scans were also used to draw lesion maps where the deformation of the tissue was visible.

We preprocessed the functional MRI scans using fMRIPrep (version 22.0.2) (Esteban et al., 2018). On the anatomical T1w scans, where lesions were visible, lesion masks were created and used as input to the fmriprep processing. The full generated reports by fmriprep from the preprocessing of all three datasets (including specific differences in preprocessing the multiecho Dataset 3) are given in the Supplementary Materials. This was followed by a 24 head-motion parameter (HMP) confounds regression (Weiler et al. 2022), removal of the first four samples, as well as low pass and high pass filtering (0.1 - 0.01 Hz). From the fMRI dataset, we excluded 26 scans due to a failure in the preprocessing (failed alignment or segmentation due to the in-scanner movements or lesion size), or following a lenient exclusion criteria check (minimum 180 volumes, and a mean framewise displacement less than 0.55 mm).

The dMRI acquisitions were preprocessed as reported by (Velly et al., 2018; Altmayer et al., 2024), further details can be found in the Supplementary Materials. 37 scans were excluded either following a manual quality check of artifacts or because the same patient was scanned twice. All scans were normalized to the MNI template. We used the Harvard-Oxford subcortical atlas (Makris et al., 2006, Frazier et al., 2005, Desikan et al., 2005, Goldstein et al, 2007) and the Schaefer-Yeo (7 networks, 100 ROI) cortical atlas (Schaefer et al., 2018) for the aMRI, fMRI, and PET acquisitions, and the JHU-DTI-SS for the dMRI scans (Mori et al., 2008). The seven networks in the Schaefer-Yeo atlas are the Default Mode Network (Default), Dorsal Attention Network (DorsAttn), Salience/Ventral Attention Network (SalVentAttn), Somato-Motor Network (SomMot), Visual Network (Vis), Limbic Network (Limbic), Control or Frontoparietal Network (Cont). The subcortical regions included in the Harvard-Oxford atlas are the brain-stem, and the left and right sides of the: accumbens, amygdala, caudate, hippocampus, pallidum, putamen, and thalamus.

#### Markers

From the aMRI scans we obtained cortical thickness values and subcortical volume estimates per patient. From the fMRI RS scans, per ROI, we computed the functional connectivity on a whole brain level. We split the functional connectivity per cortical to cortical or subcortical to subcortical connectivity. In addition, we calculated the subcortical to cortical network connectivity (median and standard deviations), as well as the counts of the ROIs having a connectivity strength in a range of 10 bins from 0 to 1. Measures of Fractional Anisotropy (FA) and Mean Diffusivity (MD) were extracted per ROI from the dMRI scans per tracts within the atlas as well as the mean FA (FA global) or MD (MD global) per voxel within the atlas. For the aMRI, fMRI, and PET, the features are combined in feature groups comprising the seven RS networks and the subcortical regions.

For the dMRI, the feature groups are split into projection fibers, commissural fibers, associative fibers, brainstem fibers, and global values, separately for FA and MD. The feature names and their associated groups are given in Supplementary Table 5.

**Positron Emission Tomography (PET)**

FDG-PET scans were conducted on selected patients during their clinical admission. The acquisition parameters, inclusion criteria, and preprocessing steps are given in Hermann et al., (2021). We applied the Schaefer-Yeo cortical (7 networks, 100 ROIs) and Harvard-Oxford subcortical atlas parcellations on an MNI-transformed brain as for the aMRI and fMRI scans. The feature groups are the same as for the aMRI and fMRI, with the addition of the two half-brain metabolic activity features in the PET data.

**Supplementary Table 1**: **Overview of patient characteristics and modalities for the diagnostic models in the French dataset.** This table provides data on 326 patients, which includes diagnostic information, etiologies, acute/chronic status, sex, and age. Counts represent the number of patients within each specified category and modality. Abbreviations: Unresponsive Wakefulness Syndrome (UWS), Minimally Conscious State (MCS), Electroencephalography (EEG), Resting State (RS), Local Global (LG) paradigm, functional Magnetic Resonance Imaging (fMRI), anatomical Magnetic Resonance Imaging (aMRI), diffusion Magnetic Resonance Imaging (dMRI), 18F-fluoro-Deoxy-Glucose Positron Emission Tomography (FDG PET), Traumatic Brain Injury (TBI).

| Modality | All Patients | UWS, MCS | Anoxia | Other & Combined | Stroke | TBI | Acute, Chronic | Sex (F,M) | Age (mean, sd) |
|---|---|---|---|---|---|---|---|---|---|
| All patients | 326 | 153, 173 | 109 | 100 | 31 | 74 | 233, 90 | 127, 194 | 46.5, 17.7 |
| EEG RS | 120 | 63, 57 | 42 | 38 | 10 | 26 | 86, 34 | 47, 73 | 47.3, 17.2 |
| EEG LG | 290 | 138, 152 | 98 | 92 | 25 | 63 | 206, 84 | 115, 175 | 46.8, 17.1 |
| fMRI RS | 44 | 21, 23 | 20 | 11 | 3 | 9 | 30, 14 | 17, 27 | 43.9, 16.7 |
| FDG PET | 53 | 21, 32 | 14 | 14 | 6 | 16 | 12, 41 | 23, 30 | 43.2, 17.3 |
| dMRI | 151 | 79, 72 | 60 | 41 | 12 | 35 | 118, 30 | 56, 90 | 45.6, 19 |
| aMRI | 101 | 54, 47 | 46 | 27 | 4 | 21 | 78, 28 | 37, 64 | 46.4, 18.1 |

**Supplementary Table 2**: **Overview of patient characteristics and modalities for prognostic models in the French dataset.** This table provides data on 232 patients, which includes diagnostic information, etiologies, acute/chronic status, sex, and age. Counts represent the number of patients within each specified category and modality. Abbreviations: Electroencephalography (EEG), Resting State (RS), Local Global (LG) paradigm, functional Magnetic Resonance Imaging (fMRI), anatomical Magnetic Resonance Imaging (aMRI), diffusion Magnetic Resonance Imaging (dMRI), Positron Emission Tomography (PET), Traumatic Brain Injury (TBI).

| Modality | All Patients | Not Improved, Improved | Anoxia | Other & Combined | Stroke | TBI | Acute, Chronic | Sex (F, M) | Age (mean, sd) |
|---|---|---|---|---|---|---|---|---|---|
| All patients | 232 | 133, 99 | 72 | 66 | 24 | 60 | 154, 78 | 93, 138 | 45, 17.05 |
| EEG RS | 89 | 50, 39 | 26 | 26 | 10 | 23 | 63, 26 | 34, 55 | 46.7, 17.6 |
| EEG LG | 207 | 117, 90 | 63 | 59 | 21 | 54 | 134, 73 | 82, 125 | 45.4, 16.8 |
| fMRI RS | 30 | 20, 10 | 11 | 6 | 3 | 9 | 17, 13 | 11, 19 | 38.7, 15.4 |
| FDG PET | 44 | 35, 9 | 9 | 12 | 6 | 14 | 8, 36 | 18, 26 | 41.9, 16.9 |
| dMRI | 95 | 49, 46 | 33 | 24 | 8 | 39 | 71, 24 | 39, 55 | 43.4, 18.2 |
| aMRI | 66 | 40, 26 | 24 | 18 | 2 | 20 | 44, 22 | 23, 43 | 44, 17.9 |

**Supplementary Table 3**: **Overview of patient characteristics and modalities for the diagnostic models in the German dataset.** This table provides data on 30 patients, which includes diagnostic information, etiologies, acute/chronic status, sex, and age. Counts represent the number of patients within each specified category and modality. Abbreviations: Unresponsive Wakefulness Syndrome (UWS), Minimally Conscious State (MCS), Electroencephalography (EEG), Resting State (RS), Local Global (LG) paradigm, functional Magnetic Resonance Imaging (fMRI), anatomical Magnetic Resonance Imaging (aMRI), Traumatic Brain Injury (TBI), Intracerebral Hemorrhage (ICH), Subarachnoid Hemorrhage (SAH).

| Modality | All Patients | UWS, MCS | Etiologies | Acute, Chronic | Sex (F, M) | Age (mean, sd) |
|---|---|---|---|---|---|---|
| All patients | 54 | 34, 20 | 11 Anoxia, 11 TBI, 10 Cardiac arrest, 8 ICH, 5 SAH, 5 Stroke, 2 Other, 1 Shock, 1 Hypoglycemia | 47, 7 | 17, 37 | 56, 15.3 |
| EEG RS | 50 | 30, 20 | 11 Anoxia, 10 TBI, 9 Cardiac arrest, 8 ICH, 4 SAH, 4 Stroke, 2 Other, 1 Shock, 1 Hypoglycemia | 43, 7 | 16, 34 | 55.8, 15.7 |
| EEG LG | 42 | 24, 18 | 9 Cardiac arrest, 9 Anoxia, 9 TBI, 5 ICH, 4 Stroke, 3 SAH, 2 Other, 1 Hypoglycemia | 36, 6 | 15, 27 | 55.2, 16 |
| aMRI | 12 | 10, 2 | 3 ICH, 3 Anoxia, 2 Cardiac arrest, 2 SAH, 1 TBI, 1 Stroke | 10, 2 | 2, 10 | 52.4, 12.5 |
| fMRI RS | 7 | 5, 2 | 3 Anoxia, 2 Cardiac arrest, 1 ICH, 1 SAH | 6, 1 | 1, 6 | 49.9, 13.9 |

**Supplementary Table 4**: **Overview of patient characteristics and modalities for the diagnostic models in the Italian dataset.** This table provides data on 30 patients, which includes diagnostic information, etiologies, acute/chronic status, sex, and age. Counts represent the number of patients within each specified category and modality. Abbreviations: Unresponsive Wakefulness Syndrome (UWS), Minimally Conscious State (MCS), Emergent Minimally Conscious State (EMCS), Electroencephalography (EEG), Resting State (RS), Local Global (LG) paradigm, functional Magnetic Resonance Imaging (fMRI), anatomical Magnetic Resonance Imaging (aMRI), Traumatic Brain Injury (TBI), Intracerebral Hemorrhage (ICH), Subarachnoid Hemorrhage (SAH).

| Modality | All Patients | UWS, MCS | Etiologies | Acute, Chronic | Sex (F, M) | Age (mean, sd) |
|---|---|---|---|---|---|---|
| All patients | 30 | 13, 17 | 13 TBI, 5 ICH, 5 Cardiac arrest, 4 SAH, 2 Stroke, 1 Other | 21, 9 | 13, 17 | 46.6, 16.9 |
| EEG RS | 25 | 12, 13 | 10 TBI, 4 ICH, 4 Cardiac arrest, 4 SAH, 2 Stroke, 1 Other | 17, 8 | 11, 14 | 45.8, 18.2 |
| EEG LG | 17 | 6, 11 | 7 TBI, 4 ICH, 2 SAH, 2 Cardiac arrest, 1 Stroke, 1 Other | 11, 6 | 8, 9 | 47.7, 18.9 |
| aMRI | 12 | 3, 9 | 6 TBI, 3 ICH, 2 Cardiac arrest, 1 SAH | 10, 2 | 6, 6 | 51.3, 16 |
| fMRI RS | 12 | 3, 9 | 6 TBI, 3 ICH, 2 Cardiac arrest, 1 SAH | 10, 2 | 6, 6 | 51.3, 16 |

#### Diffusion MRI preprocessing and markers

FA values were extracted from DTI sequences acquired as part of routine clinical multimodal brain MRI protocols. The MRI scans were performed using 1.5 or 3 Tesla machines (General Electric Healthcare, Velizy, France). The DTI images underwent preprocessing with a validated method, using brainQuant Software (www.braintale.eu; research version 2.0). The preprocessing included corrections for eddy currents and motion, followed by the computation and normalization of FA maps into MNI-152 space. FA values were then extracted from 21 predefined regions of interest, selected based on the publicly available ICBM-DTI-81 atlas, which delineates the 56 major deep white matter bundles.

The MRI data were acquired with 1.5 or 3 Tesla machines equipped with either quadratic or multi-channel head coils. Specific parameters, including echo time (TE) and repetition time (TR), varied depending on the manufacturer and scanner model. The imaging sequences included sagittal localization T1, axial T2/FLAIR, axial T2, axial T2*, 3D T1, and DTI. The DTI acquisition was performed in an axial plane perpendicular to the main magnetic field B0. The gradient value (B) for diffusion was set to 1000 mT/m and applied in 11 to 50 directions:

- 1.5 T (12 directions): 3-mm thickness, no gap, matrix 96 × 96, FOV = 28 cm, TR = 10,000ms, TE = 87ms
- 1.5 T (23 directions): 5-mm thickness, no gap, matrix 96 × 96, FOV = 28 cm, TR = 8,000ms, TE = 80ms
- 3 T (12 directions): 3-mm thickness, no gap, matrix 96 × 96, FOV = 28 cm, TR = 12,000ms, TE = 81.9ms
- 3 T (50 directions): 2.5 mm thickness, no gap, matrix 128 × 128, FOV = 28 cm, TR = 14,000ms, TE = 84.5ms

Quality control was performed on all DTI images to exclude low-quality data, ensuring protocol compliance and absence of significant motion artifacts. The images were then processed using brainQuant Software (version 2.0), employing FMRIB Software Library (FSL) preprocessing steps. Corrections for head motion and eddy-current distortions were applied, followed by brain extraction to isolate brain tissue from non-brain volumes. A diffusion tensor model was applied to each voxel, generating FA and mean diffusivity maps.

FA maps were normalized to the Montreal Neurological Institute (MNI-152) standard space using both linear and nonlinear transformations. The quality of these processes was confirmed through several checks, including the entropy of the spatial distribution of the main diffusion direction and residuals of the diffusion tensor model. Visual inspection ensured the quality of coregistration between FA maps and the atlas. To account for intersequence variability, mean FA values were normalized by scaling each patient's FA against the mean FA from a normative control group that has acquired the same imaging sequence.

**Supplementary Table 5**: **Diffusion MRI tracts abbreviations, their full names and their attributed feature groups.** The feature groups are used in Figure 4 C and Supplementary Figure 10 K and L. The grouping is taken from Altmayer et al., 2024 and Mori et al., 2008.

| Abbreviation | Full name | Attributed feature group |
|---|---|---|
| MCP | Middle cerebellar peduncle | Brainstem |
| PCT | Pontine crossing tract (a part of MCP) | Brainstem |
| gCC | Genu of corpus callosum | Commissural fibers |
| bCC | Body of corpus callosum | Commissural fibers |
| sCC | Splenium of corpus callosum | Commissural fibers |
| CST_R | Corticospinal tract - Right | Brainstem |
| CST_L | Corticospinal tract - Left | Brainstem |
| mLEM_R | Medial lemniscus - Right | Brainstem |
| mLEM_L | Medial lemniscus - Left | Brainstem |
| ICP_R | Inferior cerebellar peduncle - Right | Brainstem |
| ICP_L | Inferior cerebellar peduncle - Left | Brainstem |
| SCP_R | Superior cerebellar peduncle - Right | Brainstem |
| SCP_L | Superior cerebellar peduncle - Left | Brainstem |
| CP_R | Cerebral peduncle - Right | Projection fibers |
| CP_L | Cerebral peduncle - Left | Projection fibers |
| ALIC_R | Anterior limb of internal capsule - Right | Projection fibers |
| ALIC_L | Anterior limb of internal capsule - Left | Projection fibers |
| PLIC_R | Posterior limb of internal capsule - Right | Projection fibers |
| PLIC_L | Posterior limb of internal capsule - Left | Projection fibers |
| RLIC_R | Retrolenticular part of internal capsule - Right | Projection fibers |
| RLIC_L | Retrolenticular part of internal capsule - Left | Projection fibers |
| ACR_R | Anterior corona radiata - Right | Projection fibers |
| ACR_L | Anterior corona radiata - Left | Projection fibers |
| SCR_R | Superior corona radiata - Right | Projection fibers |
| SCR_L | Superior corona radiata - Left | Projection fibers |
| PCR_R | Posterior corona radiata - Right | Projection fibers |
| PCR_L | Posterior corona radiata - Left | Projection fibers |

| Abbreviation | Full name | Attributed feature group |
|---|---|---|
| PTR_R | Posterolateral thalamic radiation - Right | Projection fibers |
| PTR_L | Posterolateral thalamic radiation - Left | Projection fibers |
| ILF_IFOF_R | Inferior longitudinal fasciculus-inferior fronto-occipital fasciculus - Right | Associative fibers |
| ILF_IFOF_L | Inferior longitudinal fasciculus-inferior fronto-occipital fasciculus - Left | Associative fibers |
| EC_R | External capsule - Right | Associative fibers |
| EC_L | External capsule - Left | Associative fibers |
| Cing_R | Cingulum - Right | Associative fibers |
| Cing_L | Cingulum - Left | Associative fibers |
| Cing_h_R | Cingulum (hippocampus) - Right | Associative fibers |
| Cing_h_L | Cingulum (hippocampus) - Left | Associative fibers |
| SLF_R | Superior longitudinal fasciculus - Right | Associative fibers |
| SLF_L | Superior longitudinal fasciculus - Left | Associative fibers |
| SFOF_R | Superior fronto-occipital fasciculus - Right | Associative fibers |
| SFOF_L | Superior fronto-occipital fasciculus - Left | Associative fibers |
| UNC_R | Uncinate fasciculus - Right | Associative fibers |
| UNC_L | Uncinate fasciculus - Left | Associative fibers |

**Supplementary Table 6**: **Overview of attributed feature groups and reductions for EEG-based feature extraction.** Each group contains specific features and their corresponding reduction methods applied to electrodes and epochs. The feature groups are used in Figure 4 and Supplementary Figures 3,4,9,10,13. The features are output from the EEG processing package NICE, reported in Engemann et al., (2018). The features derivations are explained in detail in the Supplementary Materials of Sitt et al., (2014) and Engemann et al., (2018). The table contains all the features of the EEG LG paradigm, but the EEG RS features are a subgroup which only differs for the Evoked category.

| **Attributed Feature Group** | **Feature** | **Reductions** |
|---|---|---|
| Spectral: Low | Delta, Delta Normalized, Theta, Theta Normalized, Alpha, Alpha Normalized | *Electrodes:* TrimMean or Standard Deviation. *Epochs:* TrimMean or Standard Deviation |
| Spectral: High | Beta, Beta Normalized, Gamma, Gamma Normalized | *Electrodes:* TrimMean or Standard Deviation. *Epochs:* TrimMean or Standard Deviation |
| Spectral: Other | Spectral Entropy (SE), Median Spectrum Frequency (MSF), Spectral Edge 90 (SEF90), Spectral Edge 95 (SEF95) | *Electrodes:* TrimMean or Standard Deviation. *Epochs:* TrimMean or Standard Deviation |
| Information Theory | Permutation Entropy (PE), Kolmogorov Complexity (K) | *Electrodes:* TrimMean or Standard Deviation. *Epochs:* TrimMean or Standard Deviation |
| Connectivity | Weighted Symbolic Mutual Information (wSMI) | *Electrodes:* TrimMean or Standard Deviation. *Epochs:* TrimMean or Standard Deviation |
| Evoked Potentials | P1, P3a, P3b, LSGS-LDGD, LSGD-LDGS, LD-LS, GD-GS, Decoding Local, Decoding Global | *Electrodes:* TrimMean or Standard Deviation. *Epochs:* TrimMean or Standard Deviation |

**Supplementary Table 7**: **Neuroimaging parameters of the T1w aMRI scans from Dataset 2 (Germany).**

| Magnetic Field Strength | Manufacturer & Model Name | Voxel Size | Echo Time | Repetition Time | Flip Angle |
|---|---|---|---|---|---|
| 3 | Philips, Ingenia | 0.75x0.75x0.75 | 0.005064 | 0.0109643 | 12 |
| 3 | Philips, Ingenia | 0.75x0.75x0.75 | 0.004746 | 0.0103699 | 12 |
| 3 | Philips, Ingenia | 0.75x0.75x0.75 | 0.004968 | 0.0108009 | 12 |
| 3 | Philips, Ingenia | 0.75x0.75x0.75 | 0.004839 | 0.0105603 | 12 |
| 3 | Philips, Ingenia | 0.75x0.75x0.75 | 0.00476 | 0.0103897 | 12 |
| 3 | Philips, Ingenia | 0.75x0.75x0.75 | 0.00481 | 0.0105034 | 12 |
| 3 | Philips, Ingenia | 0.75x0.75x0.75 | 0.0049 | 0.0106729 | 12 |
| 3 | GE, Signa HDxt | 0.70x0.45x0.45 | 0.00136 | 0.004528 | 15 |
| 3 | GE, Signa HDxt | 3x0.4x0.4 | 0.134884 | 6 | 90 |
| 3 | Siemens, MAGNETOM Vida | 0.98x0.98x0.98 | 0.00321 | 2.4 | 8 |
| 3 | Siemens, MAGNETOM Vida | 0.98x1x1 | 0.00321 | 2.4 | 8 |
| 1.5 | Siemens, Aera | 1x1x1 | 0.00313 | 1.98 | 15 |

#### fmriprep Generated Report for Dataset 1 (France) and Dataset 2 (Germany)

Results included in this manuscript come from preprocessing performed using fMRIPrep 22.0.2 (Esteban, Markiewicz, et al. (2018); Esteban, Blair, et al. (2018); RRID:SCR_016216), which is based on Nipype 1.8.5 (K. Gorgolewski et al. (2011); K. Gorgolewski et al. (2018); RRID: SCR_002502).

##### Anatomical data preprocessing

A total of 1 T1-weighted (T1w) images were found within the input BIDS dataset.The T1-weighted (T1w) image was corrected for intensity non-uniformity (INU) with N4BiasFieldCorrection (Tustison et al. 2010), distributed with ANTs 2.3.3 (Avants et al. 2008, RRID:SCR_004757), and used as T1w-reference throughout the workflow. The T1w-reference was then skull-stripped with a Nipype implementation of the antsBrainExtraction.sh workflow (from ANTs), using OASIS30ANTs as target template. Brain tissue segmentation of cerebrospinal fluid (CSF), white-matter (WM) and gray-matter (GM) was performed on the brain-extracted T1w using fast (FSL 6.0.5.1:57b01774, RRID:SCR_002823, Zhang, Brady, and Smith 2001). Volume-based spatial normalization to one standard space (MNI152NLin2009cAsym) was performed through nonlinear registration with antsRegistration (ANTs 2.3.3), using brain-extracted versions of both T1w reference and the T1w template. The following template was selected for spatial normalization: ICBM 152 Nonlinear Asymmetrical template version 2009c [Fonov et al. (2009), RRID:SCR_008796; TemplateFlow ID: MNI152NLin2009cAsym].

**Functional data preprocessing**

For each of the 1 BOLD runs found per subject (across all tasks and sessions), the following preprocessing was performed. First, a reference volume and its skull-stripped version were generated using a custom methodology of fMRIPrep. Head-motion parameters with respect to the BOLD reference (transformation matrices, and six corresponding rotation and translation parameters) are estimated before any spatiotemporal filtering using mcflirt (FSL 6.0.5.1:57b01774, Jenkinson et al. 2002). BOLD runs were slice-time corrected to 0.958s (0.5 of slice acquisition range 0s-1.92s) using 3dTshift from AFNI (Cox and Hyde 1997, RRID:SCR_005927). The BOLD time-series (including slice-timing correction when applied) were resampled onto their original, native space by applying the transforms to correct for head-motion. These resampled BOLD time-series will be referred to as preprocessed BOLD in original space, or just preprocessed BOLD. The BOLD reference was then co-registered to the T1w reference using mri_coreg (FreeSurfer) followed by flirt (FSL 6.0.5.1:57b01774, Jenkinson and Smith 2001) with the boundary-based registration (Greve and Fischl 2009) cost-function. Co-registration was configured with six degrees of freedom. Several confounding time-series were calculated based on the preprocessed BOLD: framewise displacement (FD), DVARS and three region-wise global signals. FD was computed using two formulations following Power (absolute sum of relative motions, Power et al. (2014)) and Jenkinson (relative root mean square displacement between affines, Jenkinson et al. (2002)). FD and DVARS are calculated for each functional run, both using their implementations in Nipype (following the definitions by Power et al. 2014). The three global signals are extracted within the CSF, the WM, and the whole-brain masks. Additionally, a set of physiological regressors were extracted to allow for component-based noise correction (CompCor, Behzadi et al. 2007). Principal components are estimated after high-pass filtering the preprocessed BOLD time-series (using a discrete cosine filter with 128s cut-off) for the two CompCor variants: temporal (tCompCor) and anatomical (aCompCor). tCompCor components are then calculated from the top 2% variable voxels within the brain mask. For aCompCor, three probabilistic masks (CSF, WM and combined CSF+WM) are generated in anatomical space. The implementation differs from that of Behzadi et al. in that instead of eroding the masks by 2 pixels on BOLD space, a mask of pixels that likely contain a volume fraction of GM is subtracted from the aCompCor masks. This mask is obtained by thresholding the corresponding partial volume map at 0.05, and it ensures components are not extracted from voxels containing a minimal fraction of GM. Finally, these masks are resampled into BOLD space and binarized by thresholding at 0.99 (as in the original implementation). Components are also calculated separately within the WM and CSF masks. For each CompCor decomposition, the k components with the largest singular values are retained, such that the retained components' time series are sufficient to explain 50 percent of variance across the nuisance mask (CSF, WM, combined, or temporal). The remaining components are dropped from consideration. The head-motion estimates calculated in the correction step were also placed within the corresponding confounds file. The confound time series derived from head motion estimates and global signals were expanded with the inclusion of temporal derivatives and quadratic terms for each (Satterthwaite et al. 2013). Frames that exceeded a threshold of 0.5 mm FD or 1.5 standardized DVARS were annotated as motion outliers. Additional nuisance timeseries are calculated by means of principal components analysis of the signal found within a thin band (crown) of voxels around the edge of the brain, as proposed by (Patriat, Reynolds, and Birn 2017). The BOLD time-series were resampled into standard space, generating a preprocessed BOLD run in MNI152NLin2009cAsym space. First, a reference volume and its skull-stripped version were generated using a custom methodology of fMRIPrep. All resamplings can be performed with a single interpolation step by composing all the pertinent transformations (i.e. head-motion transform matrices, susceptibility distortion correction when available, and co-registrations to anatomical and output spaces). Gridded (volumetric) resamplings were performed using antsApplyTransforms (ANTs), configured with Lanczos interpolation to minimize the smoothing effects of other kernels (Lanczos 1964). Non-gridded (surface) resamplings were performed using mri_vol2surf (FreeSurfer). Many internal operations of fMRIPrep use Nilearn 0.9.1 (Abraham et al. 2014, RRID:SCR_001362), mostly within the functional processing workflow. For more details of the pipeline, see the section corresponding to workflows in fMRIPrep's documentation.

#### fmriprep Generated Report for Dataset 3 (Italy)

Results included in this manuscript come from preprocessing performed using fMRIPrep 22.0.2 (Esteban, Markiewicz, et al. (2018); Esteban, Blair, et al. (2018); RRID:SCR_016216), which is based on Nipype 1.8.5 (K. Gorgolewski et al. (2011); K. Gorgolewski et al. (2018); RRID: SCR_002502).

**Preprocessing of B0 inhomogeneity mappings**

A total of 1 fieldmaps were found available within the input BIDS structure for this particular subject. A B0-nonuniformity map (or fieldmap) was estimated based on two (or more) echo-planar imaging (EPI) references with topup (Andersson, Skare, and Ashburner (2003); FSL 6.0.5.1:57b01774).

**Anatomical data preprocessing**

A total of 1 T1-weighted (T1w) images were found within the input BIDS dataset.The T1-weighted (T1w) image was corrected for intensity non-uniformity (INU) with N4BiasFieldCorrection (Tustison et al. 2010), distributed with ANTs 2.3.3 (Avants et al. 2008, RRID:SCR_004757), and used as T1w-reference throughout the workflow. The T1w-reference was then skull-stripped with a Nipype implementation of the antsBrainExtraction.sh workflow (from ANTs), using OASIS30ANTs as target template. Brain tissue segmentation of cerebrospinal fluid (CSF), white-matter (WM) and gray-matter (GM) was performed on the brain-extracted T1w using fast (FSL 6.0.5.1:57b01774, RRID:SCR_002823, Zhang, Brady, and Smith 2001). Volume-based spatial normalization to one standard space (MNI152NLin2009cAsym) was performed through nonlinear registration with antsRegistration (ANTs 2.3.3), using brain-extracted versions of both T1w reference and the T1w template. The following template was selected for spatial normalization: ICBM 152 Nonlinear Asymmetrical template version 2009c [Fonov et al. (2009), RRID:SCR_008796; TemplateFlow ID: MNI152NLin2009cAsym].

**Functional data preprocessing**

For each of the 1 BOLD runs found per subject (across all tasks and sessions), the following preprocessing was performed. First, a reference volume and its skull-stripped version were generated from the shortest echo of the BOLD run using a custom methodology of fMRIPrep. Head-motion parameters with respect to the BOLD reference (transformation matrices, and six corresponding rotation and translation parameters) are estimated before any spatiotemporal filtering using mcflirt (FSL 6.0.5.1:57b01774, Jenkinson et al. 2002). BOLD runs were slice-time corrected to 0.45s (0.5 of slice acquisition range 0s-0.9s) using 3dTshift from AFNI (Cox and Hyde 1997, RRID:SCR_005927). The BOLD time-series (including slice-timing correction when applied) were resampled onto their original, native space by applying the transforms to correct for head-motion. These resampled BOLD time-series will be referred to as preprocessed BOLD in original space, or just preprocessed BOLD. A T2★ map was estimated from the preprocessed EPI echoes, by voxel-wise fitting the maximal number of echoes with reliable signal in that voxel to a monoexponential signal decay model with nonlinear regression. The T2★/S0 estimates from a log-linear regression fit were used for initial values. The calculated T2★ map was then used to optimally combine preprocessed BOLD across echoes following the method described in (Posse et al. 1999). The optimally combined time series was carried forward as the preprocessed BOLD. The BOLD reference was then co-registered to the T1w reference using mri_coreg (FreeSurfer) followed by flirt (FSL 6.0.5.1:57b01774, Jenkinson and Smith 2001) with the boundary-based registration (Greve and Fischl 2009) cost-function. Co-registration was configured with six degrees of freedom. First, a reference volume and its skull-stripped version were generated using a custom methodology of fMRIPrep. Several confounding time-series were calculated based on the preprocessed BOLD: framewise displacement (FD), DVARS and three region-wise global signals. FD was computed using two formulations following Power (absolute sum of relative motions, Power et al. (2014)) and Jenkinson (relative root mean square displacement between affines, Jenkinson et al. (2002)). FD and DVARS are calculated for each functional run, both using their implementations in Nipype (following the definitions by Power et al. 2014). The three global signals are extracted within the CSF, the WM, and the whole-brain masks. Additionally, a set of physiological regressors were extracted to allow for component-based noise correction (CompCor, Behzadi et al. 2007). Principal components are estimated after high-pass filtering the preprocessed BOLD time-series (using a discrete cosine filter with 128s cut-off) for the two CompCor variants: temporal (tCompCor) and anatomical (aCompCor). tCompCor components are then calculated from the top 2% variable voxels within the brain mask. For aCompCor, three probabilistic masks (CSF, WM and combined CSF+WM) are generated in anatomical space. The implementation differs from that of Behzadi et al. in that instead of eroding the masks by 2 pixels on BOLD space, a mask of pixels that likely contain a volume fraction of GM is subtracted from the aCompCor masks. This mask is obtained by thresholding the corresponding partial volume map at 0.05, and it ensures components are not extracted from voxels containing a minimal fraction of GM. Finally, these masks are resampled into BOLD space and binarized by thresholding at 0.99 (as in the original implementation). Components are also calculated separately within the WM and CSF masks. For each CompCor decomposition, the k components with the largest singular values are retained, such that the retained components' time series are sufficient to explain 50 percent of variance across the nuisance mask (CSF, WM, combined, or temporal). The remaining components are dropped from consideration. The head-motion estimates calculated in the correction step were also placed within the corresponding confounds file. The confound time series derived from head motion estimates and global signals were expanded with the inclusion of temporal derivatives and quadratic terms for each (Satterthwaite et al. 2013). Frames that exceeded a threshold of 0.5 mm FD or 1.5 standardized DVARS were annotated as motion outliers. Additional nuisance timeseries are calculated by means of principal components analysis of the signal found within a thin band (crown) of voxels around the edge of the brain, as proposed by (Patriat, Reynolds, and Birn 2017). The BOLD time-series were resampled into standard space, generating a preprocessed BOLD run in MNI152NLin2009cAsym space. First, a reference volume and its skull-stripped version were generated using a custom methodology of fMRIPrep. All resamplings can be performed with a single interpolation step by composing all the pertinent transformations (i.e. head-motion transform matrices, susceptibility distortion correction when available, and co-registrations to anatomical and output spaces). Gridded (volumetric) resamplings were performed using antsApplyTransforms (ANTs), configured with Lanczos interpolation to minimize the smoothing effects of other kernels (Lanczos 1964). Non-gridded (surface) resamplings were performed using mri_vol2surf (FreeSurfer). Many internal operations of fMRIPrep use Nilearn 0.9.1 (Abraham et al. 2014, RRID:SCR_001362), mostly within the functional processing workflow. For more details of the pipeline, see the section corresponding to workflows in fMRIPrep's documentation.

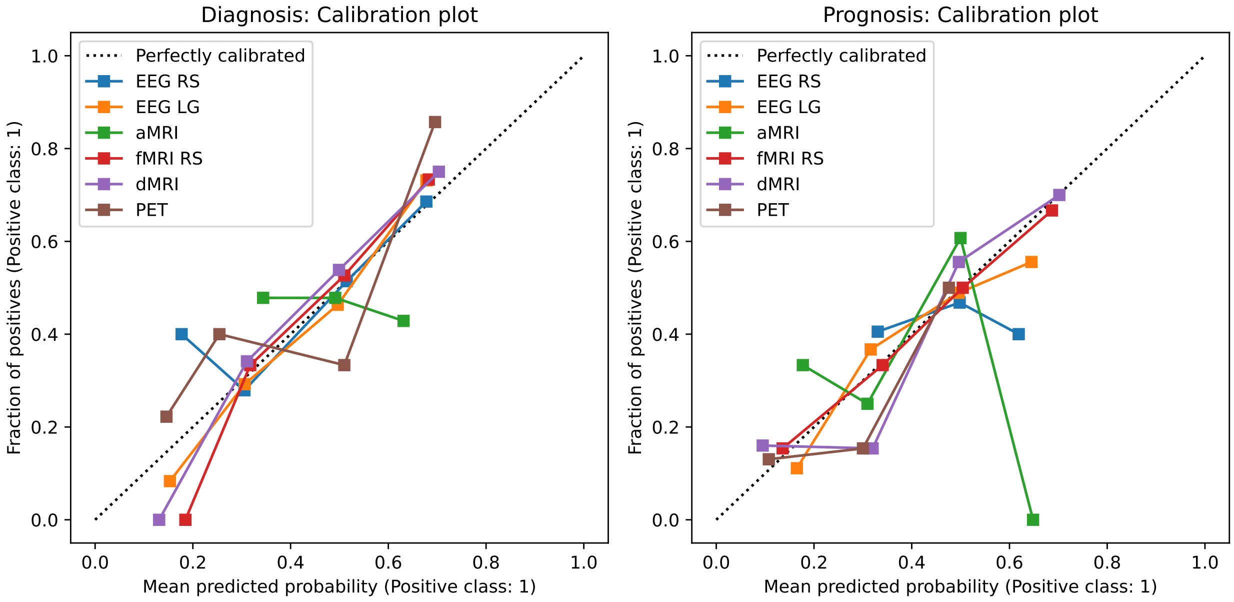


**Supplementary Figure 1**: **Calibration checks for the diagnostic and prognostic unimodal classification on Dataset 1 (France).** The diagonal denotes a perfectly calibrated model. Most models are fairly well calibrated except for the aMRI scans in the prognostic prediction. Abbreviations: Electroencephalography (EEG), Resting State (RS), Local Global (LG) paradigm, functional Magnetic Resonance Imaging (fMRI), anatomical Magnetic Resonance Imaging (aMRI), diffusion Magnetic Resonance Imaging (dMRI), Positron Emission Tomography (PET).

### Results

#### Unimodal models surrogates analysis

The diagnostic aMRI model distribution was the most similar to the surrogates distribution (Supplementary Figure 2 A). For the prognosis, EEG RS had the highest similarity to the surrogates, together with the aMRI because the surrogates median value was higher than 0.5 change level. The PET surrogates distribution more strongly differed because the model distribution has a high spread (Supplementary Figure 2 B).

**A**

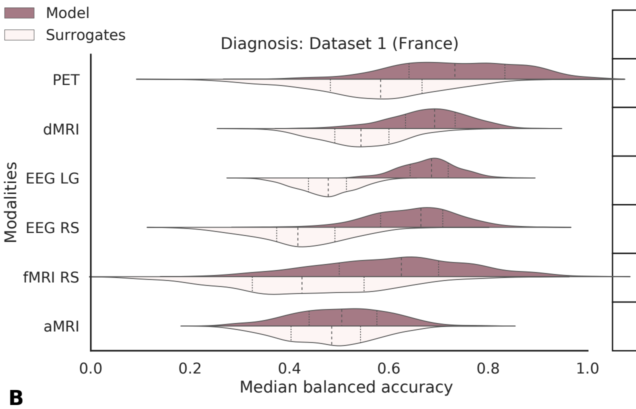


| Bal Acc | Errors | KLD | JSD | WD |
|---|---|---|---|---|
| 0.73 | 0.13 | 0.857 | 0.411 | 0.169 |
| 0.69 | 0.08 | 1.390 | 0.499 | 0.135 |
| 0.69 | 0.06 | 16.061 | 0.774 | 0.207 |
| 0.66 | 0.09 | 2.706 | 0.649 | 0.225 |
| 0.63 | 0.15 | 0.599 | 0.364 | 0.174 |
| 0.51 | 0.10 | 0.055 | 0.112 | 0.028 |

**B**

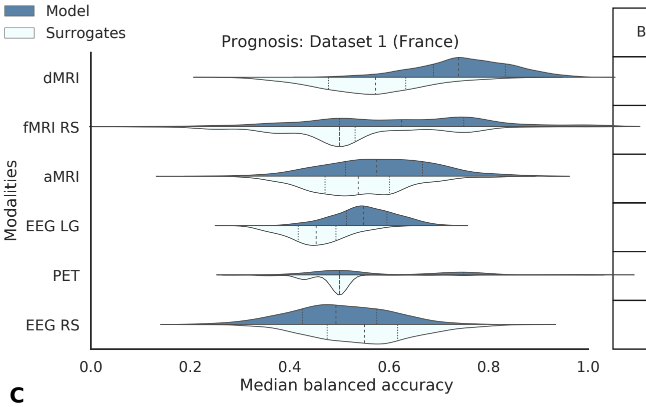


| Bal Acc | Errors | KLD | JSD | WD |
|---|---|---|---|---|
| 0.74 | 0.09 | 1.480 | 0.540 | 0.190 |
| 0.63 | 0.18 | 0.300 | 0.250 | 0.110 |
| 0.58 | 0.11 | 0.140 | 0.180 | 0.050 |
| 0.55 | 0.06 | 1.160 | 0.480 | 0.100 |
| 0.50 | 0.16 | 2.080 | 0.460 | 0.120 |
| 0.49 | 0.10 | 0.140 | 0.190 | 0.060 |

**C**

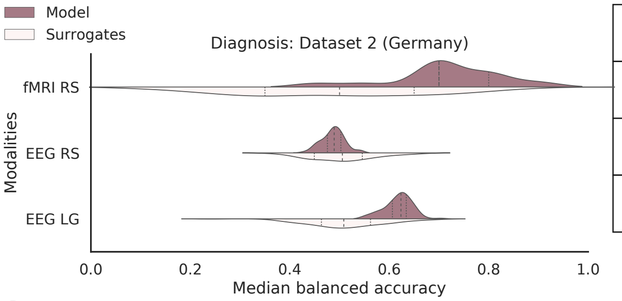


| Bal Acc | Errors | KLD | JSD | WD |
|---|---|---|---|---|
| 0.49 | 0.02 | 0.780 | 0.455 | 0.227 |
| 0.70 | 0.11 | 0.630 | 0.421 | 0.037 |
| 0.45 | 0.12 | 1.445 | 0.597 | 0.106 |

**D**

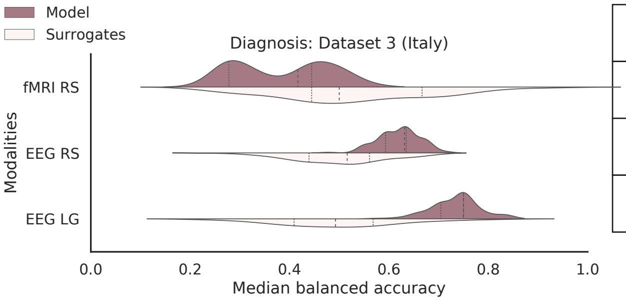


| Bal Acc | Errors | KLD | JSD | WD |
|---|---|---|---|---|
| 0.63 | 0.04 | 0.586 | 0.406 | 0.142 |
| 0.42 | 0.09 | 1.089 | 0.535 | 0.119 |
| 0.58 | 0.12 | 2.060 | 0.673 | 0.234 |

**Supplementary Figure 2**: (caption below)

**Supplementary Figure 2**: **Multivariate unimodal models' accuracy differs depending on the classification target that can be diagnostic or prognostic.** A) Unimodal Random Forest classifiers for the six modalities give a diagnostic classification accuracy which is given next to the distributions (median +/- standard deviation). The diagnostic classification patient categories are patients in UWS or MCS. The model distributions (in red or blue) are all different from the surrogates analysis (in light red or blue) performed by shuffling the labels based on Bonferroni corrected Mann-Whitney U tests. B) The same as in A) but for the prognostic categories (improved and not improved). All model distributions are significantly different from the surrogates distributions based on Bonferroni corrected Mann-Whitney U tests (given in Supplementary Materials). C) and D), same as A) but for the diagnostic classification trained on Dataset 1: France and tested on the available modalities from Dataset 2: Germany and Dataset 3: Italy. When there is n.s. it means that the model distribution is not significantly different from the surrogates one based on Bonferroni corrected Mann-Whitney U tests. Abbreviations: Electroencephalography (EEG), Resting State (RS), Local Global (LG) paradigm, functional Magnetic Resonance Imaging (fMRI), anatomical Magnetic Resonance Imaging (aMRI), diffusion Magnetic Resonance Imaging (dMRI), Positron Emission Tomography (PET), Balanced Accuracy (Bal Acc), Kullback-Leibler Divergence (KLD), Jensen-Shannon Divergence (JSD), Wasserstein Distance (WD).

#### Univariate analysis: area under the curve per feature

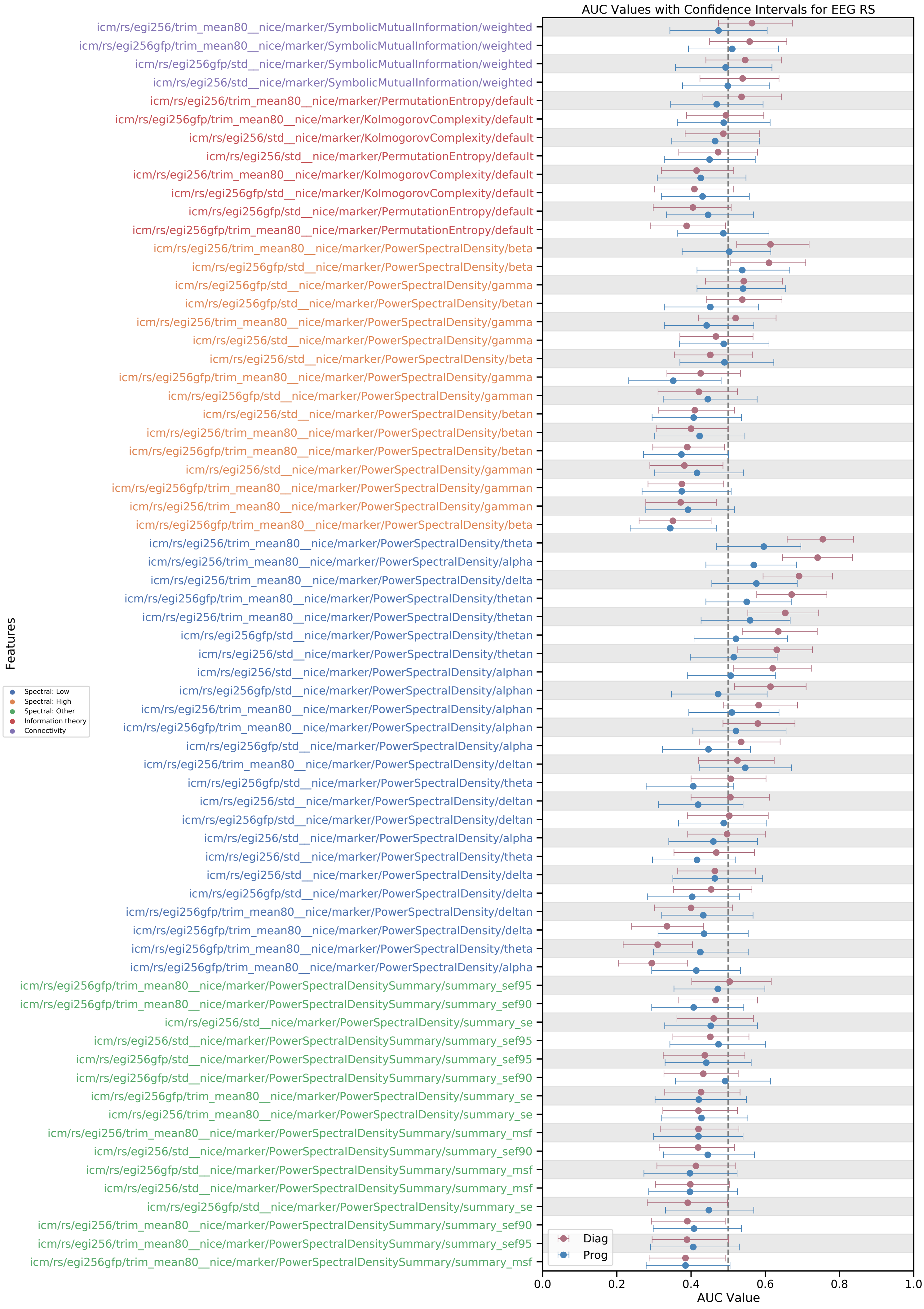
AUC Values with Confidence Intervals for EEG RS
icm/rs/egi256/trim_mean80__nice/marker/SymbolicMutualInformation/weighted
icm/rs/egi256gfp/trim_mean80__nice/marker/SymbolicMutualInformation/weighted
icm/rs/egi256gfp/std__nice/marker/SymbolicMutualInformation/weighted
icm/rs/egi256/std__nice/marker/SymbolicMutualInformation/weighted
icm/rs/egi256/trim_mean80__nice/marker/PermutationEntropy/default
icm/rs/egi256gfp/trim_mean80__nice/marker/KolmogorovComplexity/default
icm/rs/egi256/std__nice/marker/KolmogorovComplexity/default
icm/rs/egi256/std__nice/marker/PermutationEntropy/default
icm/rs/egi256/trim_mean80__nice/marker/KolmogorovComplexity/default
icm/rs/egi256gfp/std__nice/marker/KolmogorovComplexity/default
icm/rs/egi256gfp/std__nice/marker/PermutationEntropy/default
icm/rs/egi256gfp/trim_mean80__nice/marker/PermutationEntropy/default
icm/rs/egi256/trim_mean80__nice/marker/PowerSpectralDensity/beta
icm/rs/egi256gfp/std__nice/marker/PowerSpectralDensity/beta
icm/rs/egi256gfp/std__nice/marker/PowerSpectralDensity/gamma
icm/rs/egi256gfp/std__nice/marker/PowerSpectralDensity/betan
icm/rs/egi256/trim_mean80__nice/marker/PowerSpectralDensity/gamma
icm/rs/egi256/std__nice/marker/PowerSpectralDensity/gamma
icm/rs/egi256/std__nice/marker/PowerSpectralDensity/beta
icm/rs/egi256gfp/trim_mean80__nice/marker/PowerSpectralDensity/gamma
icm/rs/egi256gfp/std__nice/marker/PowerSpectralDensity/gamman
icm/rs/egi256/std__nice/marker/PowerSpectralDensity/betan
icm/rs/egi256/trim_mean80__nice/marker/PowerSpectralDensity/betan
icm/rs/egi256gfp/trim_mean80__nice/marker/PowerSpectralDensity/betan
icm/rs/egi256/std__nice/marker/PowerSpectralDensity/gamman
icm/rs/egi256gfp/trim_mean80__nice/marker/PowerSpectralDensity/gamman
icm/rs/egi256/trim_mean80__nice/marker/PowerSpectralDensity/gamman
icm/rs/egi256gfp/trim_mean80__nice/marker/PowerSpectralDensity/beta
icm/rs/egi256/trim_mean80__nice/marker/PowerSpectralDensity/theta
icm/rs/egi256/trim_mean80__nice/marker/PowerSpectralDensity/alpha
icm/rs/egi256/trim_mean80__nice/marker/PowerSpectralDensity/delta
icm/rs/egi256gfp/trim_mean80__nice/marker/PowerSpectralDensity/thetan
icm/rs/egi256/trim_mean80__nice/marker/PowerSpectralDensity/thetan
icm/rs/egi256gfp/std__nice/marker/PowerSpectralDensity/thetan
icm/rs/egi256/std__nice/marker/PowerSpectralDensity/thetan
icm/rs/egi256/std__nice/marker/PowerSpectralDensity/alphan
icm/rs/egi256gfp/std__nice/marker/PowerSpectralDensity/alphan
icm/rs/egi256/trim_mean80__nice/marker/PowerSpectralDensity/alphan
icm/rs/egi256gfp/trim_mean80__nice/marker/PowerSpectralDensity/alphan
icm/rs/egi256gfp/std__nice/marker/PowerSpectralDensity/alpha
icm/rs/egi256/trim_mean80__nice/marker/PowerSpectralDensity/deltan
icm/rs/egi256gfp/std__nice/marker/PowerSpectralDensity/theta
icm/rs/egi256/std__nice/marker/PowerSpectralDensity/deltan
icm/rs/egi256gfp/std__nice/marker/PowerSpectralDensity/deltan
icm/rs/egi256/std__nice/marker/PowerSpectralDensity/alpha
icm/rs/egi256/std__nice/marker/PowerSpectralDensity/theta
icm/rs/egi256/std__nice/marker/PowerSpectralDensity/delta
icm/rs/egi256gfp/std__nice/marker/PowerSpectralDensity/delta
icm/rs/egi256gfp/trim_mean80__nice/marker/PowerSpectralDensity/deltan
icm/rs/egi256gfp/trim_mean80__nice/marker/PowerSpectralDensity/delta
icm/rs/egi256gfp/trim_mean80__nice/marker/PowerSpectralDensity/theta
icm/rs/egi256gfp/trim_mean80__nice/marker/PowerSpectralDensity/alpha
icm/rs/egi256gfp/trim_mean80__nice/marker/PowerSpectralDensitySummary/summary_sef95
icm/rs/egi256gfp/trim_mean80__nice/marker/PowerSpectralDensitySummary/summary_sef90
icm/rs/egi256/std__nice/marker/PowerSpectralDensity/summary_se
icm/rs/egi256/std__nice/marker/PowerSpectralDensitySummary/summary_sef95
icm/rs/egi256gfp/std__nice/marker/PowerSpectralDensitySummary/summary_sef95
icm/rs/egi256gfp/std__nice/marker/PowerSpectralDensitySummary/summary_sef90
icm/rs/egi256gfp/trim_mean80__nice/marker/PowerSpectralDensity/summary_se
icm/rs/egi256/trim_mean80__nice/marker/PowerSpectralDensity/summary_se
icm/rs/egi256/trim_mean80__nice/marker/PowerSpectralDensitySummary/summary_msf
icm/rs/egi256/std__nice/marker/PowerSpectralDensitySummary/summary_sef90
icm/rs/egi256gfp/std__nice/marker/PowerSpectralDensitySummary/summary_msf
icm/rs/egi256/std__nice/marker/PowerSpectralDensitySummary/summary_msf
icm/rs/egi256gfp/std__nice/marker/PowerSpectralDensity/summary_se
icm/rs/egi256/trim_mean80__nice/marker/PowerSpectralDensitySummary/summary_sef90
icm/rs/egi256/trim_mean80__nice/marker/PowerSpectralDensitySummary/summary_sef95
icm/rs/egi256gfp/trim_mean80__nice/marker/PowerSpectralDensitySummary/summary_msf
Features
Spectral: Low
Spectral: High
Spectral: Other
Information theory
Connectivity
Diag
Prog
0.0
0.2
0.4
0.6
0.8
1.0
AUC Value

**Supplementary Figure 3**: **(Figure above) Univariate analysis for the EEG RS using the AUC values per feature for the diagnostic and prognostic targets.** The positive classes are MCS and Improved, this if the AUC value if from 0.5 to 1, then the given marker is higher in MCS or Improved patients. If the AUC value is from 0 to 0.5, then the given marker is higher in the UWS or not-Improved group of patients. Abbreviations: Institut du Cerveau (ICM), Local Global (LG), EEG cap with 256 channels (EGI256), trimmed mean taking out the 10% lowest and 10% highest end of the distribution (trim_mean80), Spectral Entropy (SE), Local Standard (LS), Local Deviant (LD), Global Standard (GS), Global Deviant (GD), Global Field Power (GFP), Permutation Entropy (PE), Median Power Frequency (MSF).

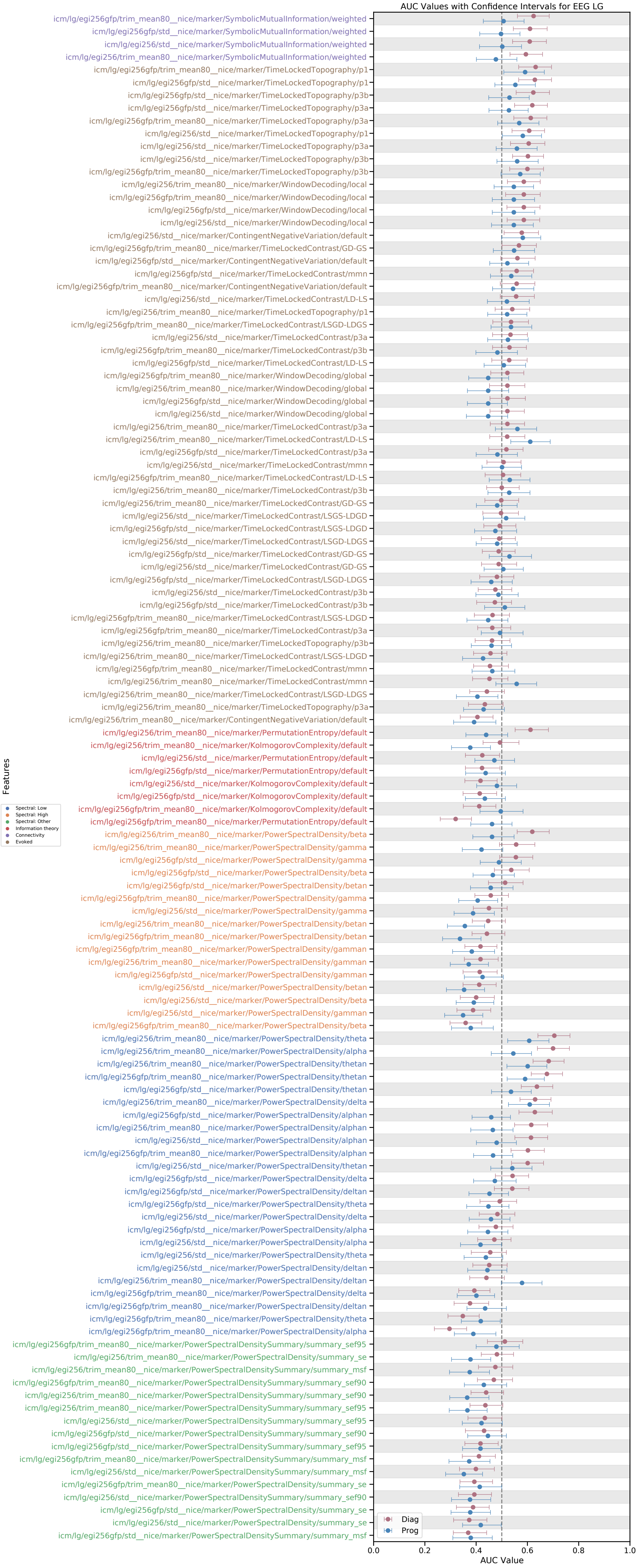
AUC Values with Confidence Intervals for EEG LG
icm/lg/egi256gfp/trim_mean80__nice/marker/SymbolicMutualInformation/weighted
icm/lg/egi256gfp/std__nice/marker/SymbolicMutualInformation/weighted
icm/lg/egi256/std__nice/marker/SymbolicMutualInformation/weighted
icm/lg/egi256/trim_mean80__nice/marker/SymbolicMutualInformation/weighted
icm/lg/egi256gfp/trim_mean80__nice/marker/TimeLockedTopography/p1
icm/lg/egi256gfp/std__nice/marker/TimeLockedTopography/p1
icm/lg/egi256gfp/std__nice/marker/TimeLockedTopography/p3b
icm/lg/egi256gfp/std__nice/marker/TimeLockedTopography/p3a
icm/lg/egi256gfp/trim_mean80__nice/marker/TimeLockedTopography/p3a
icm/lg/egi256/std__nice/marker/TimeLockedTopography/p1
icm/lg/egi256/std__nice/marker/TimeLockedTopography/p3a
icm/lg/egi256/std__nice/marker/TimeLockedTopography/p3b
icm/lg/egi256gfp/trim_mean80__nice/marker/TimeLockedTopography/p3b
icm/lg/egi256/trim_mean80__nice/marker/WindowDecoding/local
icm/lg/egi256gfp/trim_mean80__nice/marker/WindowDecoding/local
icm/lg/egi256gfp/std__nice/marker/WindowDecoding/local
icm/lg/egi256/std__nice/marker/WindowDecoding/local
icm/lg/egi256/std__nice/marker/ContingentNegativeVariation/default
icm/lg/egi256gfp/trim_mean80__nice/marker/TimeLockedContrast/GD-GS
icm/lg/egi256gfp/std__nice/marker/ContingentNegativeVariation/default
icm/lg/egi256gfp/std__nice/marker/TimeLockedContrast/mmn
icm/lg/egi256gfp/trim_mean80__nice/marker/ContingentNegativeVariation/default
icm/lg/egi256/std__nice/marker/TimeLockedContrast/LD-LS
icm/lg/egi256/trim_mean80__nice/marker/TimeLockedTopography/p1
icm/lg/egi256gfp/trim_mean80__nice/marker/TimeLockedContrast/LSGD-LDGS
icm/lg/egi256/std__nice/marker/TimeLockedContrast/p3a
icm/lg/egi256gfp/trim_mean80__nice/marker/TimeLockedContrast/p3b
icm/lg/egi256gfp/std__nice/marker/TimeLockedContrast/LD-LS
icm/lg/egi256gfp/trim_mean80__nice/marker/WindowDecoding/global
icm/lg/egi256/trim_mean80__nice/marker/WindowDecoding/global
icm/lg/egi256gfp/std__nice/marker/WindowDecoding/global
icm/lg/egi256/std__nice/marker/WindowDecoding/global
icm/lg/egi256/trim_mean80__nice/marker/TimeLockedContrast/p3a
icm/lg/egi256/trim_mean80__nice/marker/TimeLockedContrast/LD-LS
icm/lg/egi256gfp/std__nice/marker/TimeLockedContrast/p3a
icm/lg/egi256/std__nice/marker/TimeLockedContrast/mmn
icm/lg/egi256gfp/trim_mean80__nice/marker/TimeLockedContrast/LD-LS
icm/lg/egi256/trim_mean80__nice/marker/TimeLockedContrast/p3b
icm/lg/egi256/trim_mean80__nice/marker/TimeLockedContrast/GD-GS
icm/lg/egi256/std__nice/marker/TimeLockedContrast/LSGS-LDGD
icm/lg/egi256gfp/std__nice/marker/TimeLockedContrast/LSGS-LDGD
icm/lg/egi256/std__nice/marker/TimeLockedContrast/LSGD-LDGS
icm/lg/egi256gfp/std__nice/marker/TimeLockedContrast/GD-GS
icm/lg/egi256/std__nice/marker/TimeLockedContrast/GD-GS
icm/lg/egi256gfp/std__nice/marker/TimeLockedContrast/LSGD-LDGS
icm/lg/egi256/std__nice/marker/TimeLockedContrast/p3b
icm/lg/egi256gfp/std__nice/marker/TimeLockedContrast/p3b
icm/lg/egi256gfp/trim_mean80__nice/marker/TimeLockedContrast/LSGS-LDGD
icm/lg/egi256gfp/trim_mean80__nice/marker/TimeLockedContrast/p3a
icm/lg/egi256/trim_mean80__nice/marker/TimeLockedTopography/p3b
icm/lg/egi256/trim_mean80__nice/marker/TimeLockedContrast/LSGS-LDGD
icm/lg/egi256gfp/trim_mean80__nice/marker/TimeLockedContrast/mmn
icm/lg/egi256/trim_mean80__nice/marker/TimeLockedContrast/mmn
icm/lg/egi256/trim_mean80__nice/marker/TimeLockedContrast/LSGD-LDGS
icm/lg/egi256/trim_mean80__nice/marker/TimeLockedTopography/p3a
icm/lg/egi256/trim_mean80__nice/marker/ContingentNegativeVariation/default
icm/lg/egi256/trim_mean80__nice/marker/PermutationEntropy/default
icm/lg/egi256/trim_mean80__nice/marker/KolmogorovComplexity/default
icm/lg/egi256/std__nice/marker/PermutationEntropy/default
icm/lg/egi256gfp/std__nice/marker/PermutationEntropy/default
icm/lg/egi256/std__nice/marker/KolmogorovComplexity/default
icm/lg/egi256gfp/std__nice/marker/KolmogorovComplexity/default
icm/lg/egi256gfp/trim_mean80__nice/marker/KolmogorovComplexity/default
icm/lg/egi256gfp/trim_mean80__nice/marker/PermutationEntropy/default
icm/lg/egi256/trim_mean80__nice/marker/PowerSpectralDensity/beta
icm/lg/egi256/trim_mean80__nice/marker/PowerSpectralDensity/gamma
icm/lg/egi256gfp/std__nice/marker/PowerSpectralDensity/gamma
icm/lg/egi256gfp/std__nice/marker/PowerSpectralDensity/beta
icm/lg/egi256gfp/std__nice/marker/PowerSpectralDensity/betan
icm/lg/egi256gfp/trim_mean80__nice/marker/PowerSpectralDensity/gamma
icm/lg/egi256/std__nice/marker/PowerSpectralDensity/gamma
icm/lg/egi256/trim_mean80__nice/marker/PowerSpectralDensity/betan
icm/lg/egi256gfp/trim_mean80__nice/marker/PowerSpectralDensity/betan
icm/lg/egi256gfp/trim_mean80__nice/marker/PowerSpectralDensity/gamman
icm/lg/egi256/trim_mean80__nice/marker/PowerSpectralDensity/gamman
icm/lg/egi256gfp/std__nice/marker/PowerSpectralDensity/gamman
icm/lg/egi256/std__nice/marker/PowerSpectralDensity/betan
icm/lg/egi256/std__nice/marker/PowerSpectralDensity/beta
icm/lg/egi256/std__nice/marker/PowerSpectralDensity/gamman
icm/lg/egi256gfp/trim_mean80__nice/marker/PowerSpectralDensity/beta
icm/lg/egi256/trim_mean80__nice/marker/PowerSpectralDensity/theta
icm/lg/egi256/trim_mean80__nice/marker/PowerSpectralDensity/alpha
icm/lg/egi256/trim_mean80__nice/marker/PowerSpectralDensity/thetan
icm/lg/egi256gfp/trim_mean80__nice/marker/PowerSpectralDensity/thetan
icm/lg/egi256gfp/std__nice/marker/PowerSpectralDensity/thetan
icm/lg/egi256/trim_mean80__nice/marker/PowerSpectralDensity/delta
icm/lg/egi256gfp/std__nice/marker/PowerSpectralDensity/alphan
icm/lg/egi256/trim_mean80__nice/marker/PowerSpectralDensity/alphan
icm/lg/egi256/std__nice/marker/PowerSpectralDensity/alphan
icm/lg/egi256gfp/trim_mean80__nice/marker/PowerSpectralDensity/alphan
icm/lg/egi256/std__nice/marker/PowerSpectralDensity/thetan
icm/lg/egi256gfp/std__nice/marker/PowerSpectralDensity/delta
icm/lg/egi256gfp/std__nice/marker/PowerSpectralDensity/deltan
icm/lg/egi256gfp/std__nice/marker/PowerSpectralDensity/theta
icm/lg/egi256/std__nice/marker/PowerSpectralDensity/delta
icm/lg/egi256gfp/std__nice/marker/PowerSpectralDensity/alpha
icm/lg/egi256/std__nice/marker/PowerSpectralDensity/alpha
icm/lg/egi256/std__nice/marker/PowerSpectralDensity/theta
icm/lg/egi256/std__nice/marker/PowerSpectralDensity/deltan
icm/lg/egi256/trim_mean80__nice/marker/PowerSpectralDensity/deltan
icm/lg/egi256gfp/trim_mean80__nice/marker/PowerSpectralDensity/delta
icm/lg/egi256gfp/trim_mean80__nice/marker/PowerSpectralDensity/deltan
icm/lg/egi256gfp/trim_mean80__nice/marker/PowerSpectralDensity/theta
icm/lg/egi256gfp/trim_mean80__nice/marker/PowerSpectralDensity/alpha
icm/lg/egi256gfp/trim_mean80__nice/marker/PowerSpectralDensitySummary/summary_sef95
icm/lg/egi256/trim_mean80__nice/marker/PowerSpectralDensity/summary_se
icm/lg/egi256/trim_mean80__nice/marker/PowerSpectralDensitySummary/summary_msf
icm/lg/egi256gfp/trim_mean80__nice/marker/PowerSpectralDensitySummary/summary_sef90
icm/lg/egi256/trim_mean80__nice/marker/PowerSpectralDensitySummary/summary_sef90
icm/lg/egi256/trim_mean80__nice/marker/PowerSpectralDensitySummary/summary_sef95
icm/lg/egi256/std__nice/marker/PowerSpectralDensitySummary/summary_sef95
icm/lg/egi256gfp/std__nice/marker/PowerSpectralDensitySummary/summary_sef90
icm/lg/egi256gfp/std__nice/marker/PowerSpectralDensitySummary/summary_sef95
icm/lg/egi256gfp/trim_mean80__nice/marker/PowerSpectralDensitySummary/summary_msf
icm/lg/egi256/std__nice/marker/PowerSpectralDensitySummary/summary_msf
icm/lg/egi256gfp/trim_mean80__nice/marker/PowerSpectralDensity/summary_se
icm/lg/egi256/std__nice/marker/PowerSpectralDensitySummary/summary_sef90
icm/lg/egi256gfp/std__nice/marker/PowerSpectralDensity/summary_se
icm/lg/egi256/std__nice/marker/PowerSpectralDensity/summary_se
icm/lg/egi256gfp/std__nice/marker/PowerSpectralDensitySummary/summary_msf
Features
Spectral: Low
Spectral: High
Spectral: Other
Information theory
Connectivity
Evoked
Diag
Prog
0.0
0.2
0.4
0.6
0.8
1.0
AUC Value

**Supplementary Figure 4**: **(Figure above) Univariate analysis for the EEG LG using the AUC values per feature for the diagnostic and prognostic targets.** The positive classes are MCS and Improved, this if the AUC value if from 0.5 to 1, then the given marker is higher in MCS or Improved patients. If the AUC value is from 0 to 0.5, then the given marker is higher in the UWS or not-Improved group of patients. Abbreviations: Institut du Cerveau (ICM), Local Global (LG), EEG cap with 256 channels (EGI256), trimmed mean taking out the 10% lowest and 10% highest end of the distribution (trim_mean80), Spectral Entropy (SE), Local Standard (LS), Local Deviant (LD), Global Standard (GS), Global Deviant (GD), Global Field Power (GFP), Permutation Entropy (PE), Median Power Frequency (MSF).

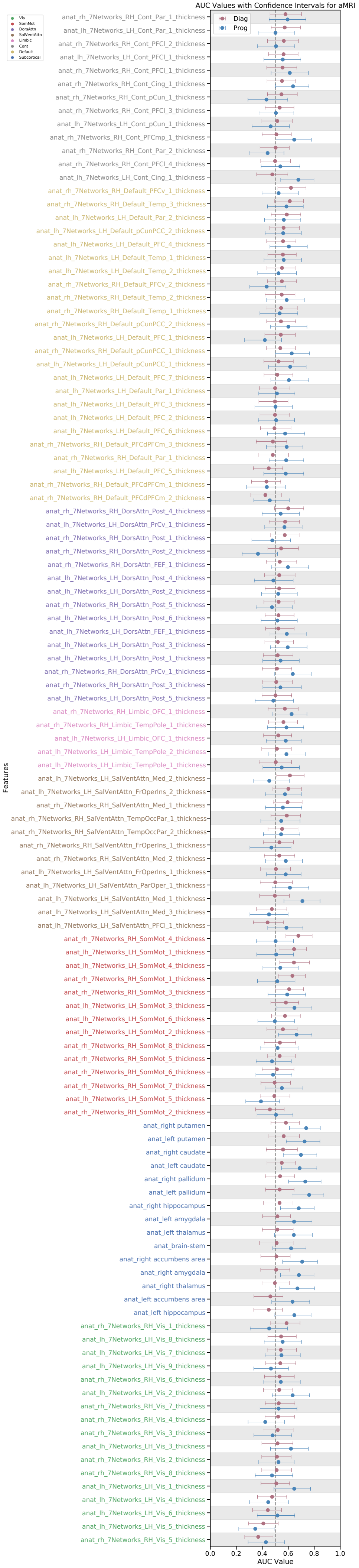
AUC Values with Confidence Intervals for aMRI
Vis
SomMot
DorsAttn
SalVentAttn
Limbic
Cont
Default
Subcortical
Diag
Prog
Features
anat_rh_7Networks_RH_Cont_Par_1_thickness
anat_lh_7Networks_LH_Cont_Par_1_thickness
anat_rh_7Networks_RH_Cont_PFCl_2_thickness
anat_lh_7Networks_LH_Cont_PFCl_1_thickness
anat_rh_7Networks_RH_Cont_PFCl_1_thickness
anat_rh_7Networks_RH_Cont_Cing_1_thickness
anat_rh_7Networks_RH_Cont_pCun_1_thickness
anat_rh_7Networks_RH_Cont_PFCl_3_thickness
anat_lh_7Networks_LH_Cont_pCun_1_thickness
anat_rh_7Networks_RH_Cont_PFCmp_1_thickness
anat_rh_7Networks_RH_Cont_Par_2_thickness
anat_rh_7Networks_RH_Cont_PFCl_4_thickness
anat_lh_7Networks_LH_Cont_Cing_1_thickness
anat_rh_7Networks_RH_Default_PFCv_1_thickness
anat_rh_7Networks_RH_Default_Temp_3_thickness
anat_lh_7Networks_LH_Default_Par_2_thickness
anat_lh_7Networks_LH_Default_pCunPCC_2_thickness
anat_lh_7Networks_LH_Default_PFC_4_thickness
anat_lh_7Networks_LH_Default_Temp_1_thickness
anat_lh_7Networks_LH_Default_Temp_2_thickness
anat_rh_7Networks_RH_Default_PFCv_2_thickness
anat_rh_7Networks_RH_Default_Temp_2_thickness
anat_rh_7Networks_RH_Default_Temp_1_thickness
anat_rh_7Networks_RH_Default_pCunPCC_2_thickness
anat_lh_7Networks_LH_Default_PFC_1_thickness
anat_rh_7Networks_RH_Default_pCunPCC_1_thickness
anat_lh_7Networks_LH_Default_pCunPCC_1_thickness
anat_lh_7Networks_LH_Default_PFC_7_thickness
anat_lh_7Networks_LH_Default_Par_1_thickness
anat_lh_7Networks_LH_Default_PFC_3_thickness
anat_lh_7Networks_LH_Default_PFC_2_thickness
anat_lh_7Networks_LH_Default_PFC_6_thickness
anat_rh_7Networks_RH_Default_PFCdPFCm_3_thickness
anat_rh_7Networks_RH_Default_Par_1_thickness
anat_lh_7Networks_LH_Default_PFC_5_thickness
anat_rh_7Networks_RH_Default_PFCdPFCm_1_thickness
anat_rh_7Networks_RH_Default_PFCdPFCm_2_thickness
anat_rh_7Networks_RH_DorsAttn_Post_4_thickness
anat_lh_7Networks_LH_DorsAttn_PrCv_1_thickness
anat_rh_7Networks_RH_DorsAttn_Post_1_thickness
anat_rh_7Networks_RH_DorsAttn_Post_2_thickness
anat_rh_7Networks_RH_DorsAttn_FEF_1_thickness
anat_lh_7Networks_LH_DorsAttn_Post_4_thickness
anat_lh_7Networks_LH_DorsAttn_Post_2_thickness
anat_rh_7Networks_RH_DorsAttn_Post_5_thickness
anat_lh_7Networks_LH_DorsAttn_Post_6_thickness
anat_lh_7Networks_LH_DorsAttn_FEF_1_thickness
anat_lh_7Networks_LH_DorsAttn_Post_3_thickness
anat_lh_7Networks_LH_DorsAttn_Post_1_thickness
anat_rh_7Networks_RH_DorsAttn_PrCv_1_thickness
anat_rh_7Networks_RH_DorsAttn_Post_3_thickness
anat_lh_7Networks_LH_DorsAttn_Post_5_thickness
anat_rh_7Networks_RH_Limbic_OFC_1_thickness
anat_rh_7Networks_RH_Limbic_TempPole_1_thickness
anat_lh_7Networks_LH_Limbic_OFC_1_thickness
anat_lh_7Networks_LH_Limbic_TempPole_2_thickness
anat_lh_7Networks_LH_Limbic_TempPole_1_thickness
anat_lh_7Networks_LH_SalVentAttn_Med_2_thickness
anat_lh_7Networks_LH_SalVentAttn_FrOperIns_2_thickness
anat_rh_7Networks_RH_SalVentAttn_Med_1_thickness
anat_rh_7Networks_RH_SalVentAttn_TempOccPar_1_thickness
anat_rh_7Networks_RH_SalVentAttn_TempOccPar_2_thickness
anat_rh_7Networks_RH_SalVentAttn_FrOperIns_1_thickness
anat_rh_7Networks_RH_SalVentAttn_Med_2_thickness
anat_lh_7Networks_LH_SalVentAttn_FrOperIns_1_thickness
anat_lh_7Networks_LH_SalVentAttn_ParOper_1_thickness
anat_lh_7Networks_LH_SalVentAttn_Med_1_thickness
anat_lh_7Networks_LH_SalVentAttn_Med_3_thickness
anat_lh_7Networks_LH_SalVentAttn_PFCl_1_thickness
anat_rh_7Networks_RH_SomMot_4_thickness
anat_lh_7Networks_LH_SomMot_1_thickness
anat_lh_7Networks_LH_SomMot_4_thickness
anat_rh_7Networks_RH_SomMot_1_thickness
anat_rh_7Networks_RH_SomMot_3_thickness
anat_lh_7Networks_LH_SomMot_3_thickness
anat_lh_7Networks_LH_SomMot_6_thickness
anat_lh_7Networks_LH_SomMot_2_thickness
anat_rh_7Networks_RH_SomMot_8_thickness
anat_rh_7Networks_RH_SomMot_5_thickness
anat_rh_7Networks_RH_SomMot_6_thickness
anat_rh_7Networks_RH_SomMot_7_thickness
anat_lh_7Networks_LH_SomMot_5_thickness
anat_rh_7Networks_RH_SomMot_2_thickness
anat_right putamen
anat_left putamen
anat_right caudate
anat_left caudate
anat_right pallidum
anat_left pallidum
anat_right hippocampus
anat_left amygdala
anat_left thalamus
anat_brain-stem
anat_right accumbens area
anat_right amygdala
anat_right thalamus
anat_left accumbens area
anat_left hippocampus
anat_rh_7Networks_RH_Vis_1_thickness
anat_lh_7Networks_LH_Vis_8_thickness
anat_lh_7Networks_LH_Vis_7_thickness
anat_lh_7Networks_LH_Vis_9_thickness
anat_rh_7Networks_RH_Vis_6_thickness
anat_lh_7Networks_LH_Vis_2_thickness
anat_rh_7Networks_RH_Vis_7_thickness
anat_rh_7Networks_RH_Vis_4_thickness
anat_rh_7Networks_RH_Vis_3_thickness
anat_lh_7Networks_LH_Vis_3_thickness
anat_rh_7Networks_RH_Vis_2_thickness
anat_rh_7Networks_RH_Vis_8_thickness
anat_lh_7Networks_LH_Vis_1_thickness
anat_lh_7Networks_LH_Vis_4_thickness
anat_lh_7Networks_LH_Vis_6_thickness
anat_lh_7Networks_LH_Vis_5_thickness
anat_rh_7Networks_RH_Vis_5_thickness
0.0
0.2
0.4
0.6
0.8
1.0
AUC Value

**Supplementary Figure 5**: **(Figure above) Univariate analysis for the aMRI using the AUC values per feature for the diagnostic and prognostic targets.** The positive classes are MCS and Improved, this if the AUC value if from 0.5 to 1, then the given marker is higher in MCS or Improved patients. If the AUC value is from 0 to 0.5, then the given marker is higher in the UWS or not-Improved group of patients.

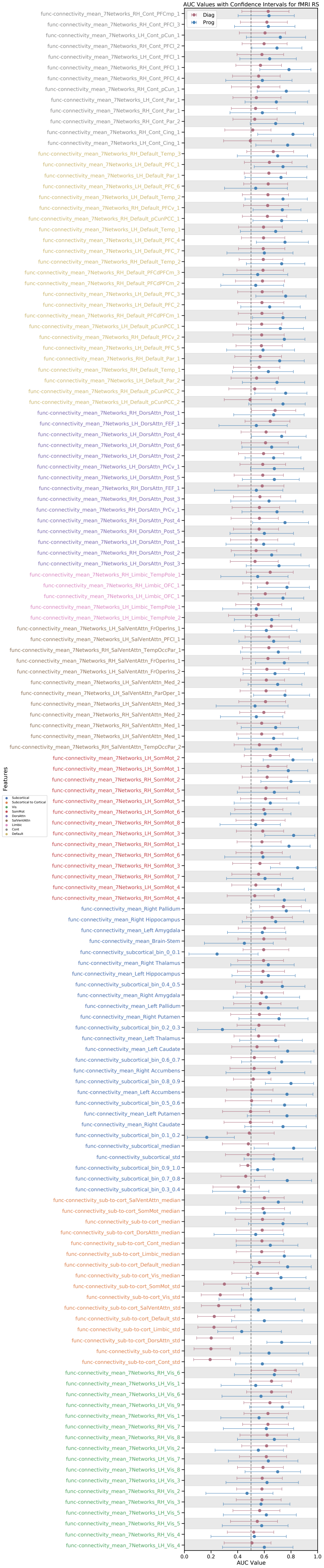
AUC Values with Confidence Intervals for fMRI RS
Diag
Prog
Features
Subcortical
Subcortical to Cortical
Vis
SomMot
DorsAttn
SalVentAttn
Limbic
Cont
Default
func-connectivity_mean_7Networks_RH_Cont_PFCmp_1
func-connectivity_mean_7Networks_RH_Cont_PFCl_3
func-connectivity_mean_7Networks_LH_Cont_pCun_1
func-connectivity_mean_7Networks_RH_Cont_PFCl_2
func-connectivity_mean_7Networks_LH_Cont_PFCl_1
func-connectivity_mean_7Networks_RH_Cont_PFCl_1
func-connectivity_mean_7Networks_RH_Cont_PFCl_4
func-connectivity_mean_7Networks_RH_Cont_pCun_1
func-connectivity_mean_7Networks_LH_Cont_Par_1
func-connectivity_mean_7Networks_RH_Cont_Par_1
func-connectivity_mean_7Networks_RH_Cont_Par_2
func-connectivity_mean_7Networks_RH_Cont_Cing_1
func-connectivity_mean_7Networks_LH_Cont_Cing_1
func-connectivity_mean_7Networks_RH_Default_Temp_3
func-connectivity_mean_7Networks_LH_Default_PFC_1
func-connectivity_mean_7Networks_LH_Default_Par_1
func-connectivity_mean_7Networks_LH_Default_PFC_6
func-connectivity_mean_7Networks_LH_Default_Temp_2
func-connectivity_mean_7Networks_RH_Default_PFCv_1
func-connectivity_mean_7Networks_RH_Default_pCunPCC_1
func-connectivity_mean_7Networks_LH_Default_Temp_1
func-connectivity_mean_7Networks_LH_Default_PFC_4
func-connectivity_mean_7Networks_LH_Default_PFC_7
func-connectivity_mean_7Networks_RH_Default_Temp_2
func-connectivity_mean_7Networks_RH_Default_PFCdPFCm_3
func-connectivity_mean_7Networks_RH_Default_PFCdPFCm_2
func-connectivity_mean_7Networks_LH_Default_PFC_3
func-connectivity_mean_7Networks_LH_Default_PFC_2
func-connectivity_mean_7Networks_RH_Default_PFCdPFCm_1
func-connectivity_mean_7Networks_LH_Default_pCunPCC_1
func-connectivity_mean_7Networks_RH_Default_PFCv_2
func-connectivity_mean_7Networks_LH_Default_PFC_5
func-connectivity_mean_7Networks_RH_Default_Par_1
func-connectivity_mean_7Networks_RH_Default_Temp_1
func-connectivity_mean_7Networks_LH_Default_Par_2
func-connectivity_mean_7Networks_RH_Default_pCunPCC_2
func-connectivity_mean_7Networks_LH_Default_pCunPCC_2
func-connectivity_mean_7Networks_RH_DorsAttn_Post_1
func-connectivity_mean_7Networks_LH_DorsAttn_FEF_1
func-connectivity_mean_7Networks_LH_DorsAttn_Post_4
func-connectivity_mean_7Networks_LH_DorsAttn_Post_6
func-connectivity_mean_7Networks_LH_DorsAttn_Post_2
func-connectivity_mean_7Networks_LH_DorsAttn_PrCv_1
func-connectivity_mean_7Networks_LH_DorsAttn_Post_5
func-connectivity_mean_7Networks_RH_DorsAttn_FEF_1
func-connectivity_mean_7Networks_RH_DorsAttn_Post_3
func-connectivity_mean_7Networks_RH_DorsAttn_PrCv_1
func-connectivity_mean_7Networks_RH_DorsAttn_Post_4
func-connectivity_mean_7Networks_RH_DorsAttn_Post_5
func-connectivity_mean_7Networks_LH_DorsAttn_Post_1
func-connectivity_mean_7Networks_RH_DorsAttn_Post_2
func-connectivity_mean_7Networks_LH_DorsAttn_Post_3
func-connectivity_mean_7Networks_RH_Limbic_TempPole_1
func-connectivity_mean_7Networks_RH_Limbic_OFC_1
func-connectivity_mean_7Networks_LH_Limbic_OFC_1
func-connectivity_mean_7Networks_LH_Limbic_TempPole_1
func-connectivity_mean_7Networks_LH_Limbic_TempPole_2
func-connectivity_mean_7Networks_LH_SalVentAttn_FrOperIns_1
func-connectivity_mean_7Networks_LH_SalVentAttn_PFCl_1
func-connectivity_mean_7Networks_RH_SalVentAttn_TempOccPar_1
func-connectivity_mean_7Networks_RH_SalVentAttn_FrOperIns_1
func-connectivity_mean_7Networks_LH_SalVentAttn_FrOperIns_2
func-connectivity_mean_7Networks_LH_SalVentAttn_Med_2
func-connectivity_mean_7Networks_LH_SalVentAttn_ParOper_1
func-connectivity_mean_7Networks_LH_SalVentAttn_Med_3
func-connectivity_mean_7Networks_RH_SalVentAttn_Med_2
func-connectivity_mean_7Networks_RH_SalVentAttn_Med_1
func-connectivity_mean_7Networks_LH_SalVentAttn_Med_1
func-connectivity_mean_7Networks_RH_SalVentAttn_TempOccPar_2
func-connectivity_mean_7Networks_LH_SomMot_2
func-connectivity_mean_7Networks_LH_SomMot_1
func-connectivity_mean_7Networks_RH_SomMot_2
func-connectivity_mean_7Networks_RH_SomMot_5
func-connectivity_mean_7Networks_LH_SomMot_5
func-connectivity_mean_7Networks_LH_SomMot_6
func-connectivity_mean_7Networks_RH_SomMot_8
func-connectivity_mean_7Networks_LH_SomMot_3
func-connectivity_mean_7Networks_RH_SomMot_1
func-connectivity_mean_7Networks_RH_SomMot_6
func-connectivity_mean_7Networks_RH_SomMot_3
func-connectivity_mean_7Networks_RH_SomMot_7
func-connectivity_mean_7Networks_LH_SomMot_4
func-connectivity_mean_7Networks_RH_SomMot_4
func-connectivity_mean_Right Pallidum
func-connectivity_mean_Right Hippocampus
func-connectivity_mean_Left Amygdala
func-connectivity_mean_Brain-Stem
func-connectivity_subcortical_bin_0_0.1
func-connectivity_mean_Right Thalamus
func-connectivity_mean_Left Hippocampus
func-connectivity_subcortical_bin_0.4_0.5
func-connectivity_mean_Right Amygdala
func-connectivity_mean_Left Pallidum
func-connectivity_mean_Right Putamen
func-connectivity_subcortical_bin_0.2_0.3
func-connectivity_mean_Left Thalamus
func-connectivity_mean_Left Caudate
func-connectivity_subcortical_bin_0.6_0.7
func-connectivity_mean_Right Accumbens
func-connectivity_subcortical_bin_0.8_0.9
func-connectivity_mean_Left Accumbens
func-connectivity_subcortical_bin_0.5_0.6
func-connectivity_mean_Left Putamen
func-connectivity_mean_Right Caudate
func-connectivity_subcortical_bin_0.1_0.2
func-connectivity_subcortical_median
func-connectivity_subcortical_std
func-connectivity_subcortical_bin_0.9_1.0
func-connectivity_subcortical_bin_0.7_0.8
func-connectivity_subcortical_bin_0.3_0.4
func-connectivity_sub-to-cort_SalVentAttn_median
func-connectivity_sub-to-cort_SomMot_median
func-connectivity_sub-to-cort_median
func-connectivity_sub-to-cort_DorsAttn_median
func-connectivity_sub-to-cort_Cont_median
func-connectivity_sub-to-cort_Limbic_median
func-connectivity_sub-to-cort_Default_median
func-connectivity_sub-to-cort_Vis_median
func-connectivity_sub-to-cort_SomMot_std
func-connectivity_sub-to-cort_Vis_std
func-connectivity_sub-to-cort_SalVentAttn_std
func-connectivity_sub-to-cort_Default_std
func-connectivity_sub-to-cort_Limbic_std
func-connectivity_sub-to-cort_DorsAttn_std
func-connectivity_sub-to-cort_std
func-connectivity_sub-to-cort_Cont_std
func-connectivity_mean_7Networks_RH_Vis_6
func-connectivity_mean_7Networks_LH_Vis_1
func-connectivity_mean_7Networks_LH_Vis_6
func-connectivity_mean_7Networks_LH_Vis_9
func-connectivity_mean_7Networks_RH_Vis_1
func-connectivity_mean_7Networks_RH_Vis_7
func-connectivity_mean_7Networks_RH_Vis_8
func-connectivity_mean_7Networks_LH_Vis_2
func-connectivity_mean_7Networks_LH_Vis_7
func-connectivity_mean_7Networks_LH_Vis_8
func-connectivity_mean_7Networks_LH_Vis_3
func-connectivity_mean_7Networks_RH_Vis_2
func-connectivity_mean_7Networks_RH_Vis_3
func-connectivity_mean_7Networks_LH_Vis_5
func-connectivity_mean_7Networks_RH_Vis_5
func-connectivity_mean_7Networks_RH_Vis_4
func-connectivity_mean_7Networks_LH_Vis_4
0.0
0.2
0.4
0.6
0.8
1.0
AUC Value

**Supplementary Figure 6**: **(Figure above) Univariate analysis for the fMRI RS using the AUC values per feature for the diagnostic and prognostic targets.** The positive classes are MCS and Improved, this if the AUC value if from 0.5 to 1, then the given marker is higher in MCS or Improved patients. If the AUC value is from 0 to 0.5, then the given marker is higher in the UWS or not-Improved group of patients.

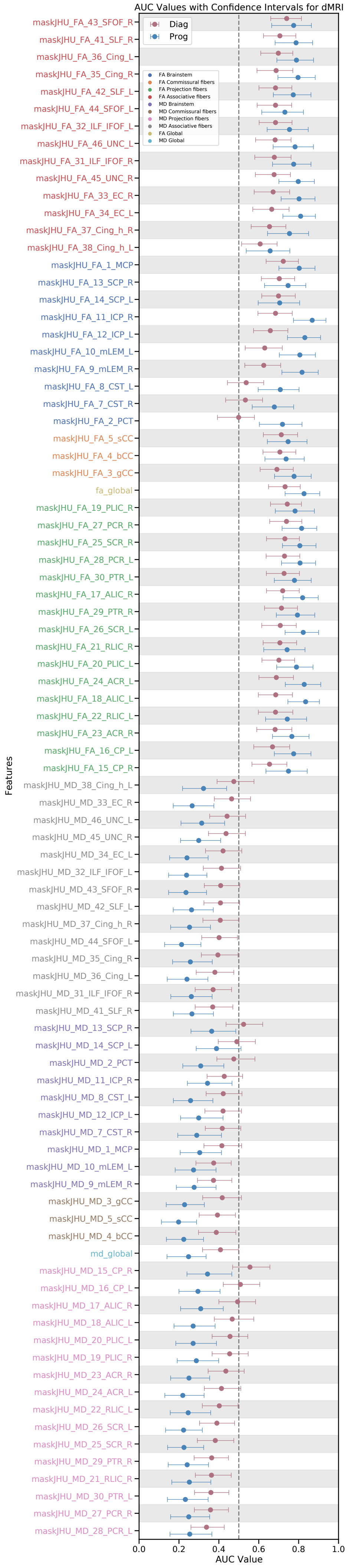

AUC Values with Confidence Intervals for dMRI
Diag
Prog
FA Brainstem
FA Commissural fibers
FA Projection fibers
FA Associative fibers
MD Brainstem
MD Commissural fibers
MD Projection fibers
MD Associative fibers
FA Global
MD Global
maskJHU_FA_43_SFOF_R
maskJHU_FA_41_SLF_R
maskJHU_FA_36_Cing_L
maskJHU_FA_35_Cing_R
maskJHU_FA_42_SLF_L
maskJHU_FA_44_SFOF_L
maskJHU_FA_32_ILF_IFOF_L
maskJHU_FA_46_UNC_L
maskJHU_FA_31_ILF_IFOF_R
maskJHU_FA_45_UNC_R
maskJHU_FA_33_EC_R
maskJHU_FA_34_EC_L
maskJHU_FA_37_Cing_h_R
maskJHU_FA_38_Cing_h_L
maskJHU_FA_1_MCP
maskJHU_FA_13_SCP_R
maskJHU_FA_14_SCP_L
maskJHU_FA_11_ICP_R
maskJHU_FA_12_ICP_L
maskJHU_FA_10_mLEM_L
maskJHU_FA_9_mLEM_R
maskJHU_FA_8_CST_L
maskJHU_FA_7_CST_R
maskJHU_FA_2_PCT
maskJHU_FA_5_sCC
maskJHU_FA_4_bCC
maskJHU_FA_3_gCC
fa_global
maskJHU_FA_19_PLIC_R
maskJHU_FA_27_PCR_R
maskJHU_FA_25_SCR_R
maskJHU_FA_28_PCR_L
maskJHU_FA_30_PTR_L
maskJHU_FA_17_ALIC_R
maskJHU_FA_29_PTR_R
maskJHU_FA_26_SCR_L
maskJHU_FA_21_RLIC_R
maskJHU_FA_20_PLIC_L
maskJHU_FA_24_ACR_L
maskJHU_FA_18_ALIC_L
maskJHU_FA_22_RLIC_L
maskJHU_FA_23_ACR_R
maskJHU_FA_16_CP_L
maskJHU_FA_15_CP_R
maskJHU_MD_38_Cing_h_L
maskJHU_MD_33_EC_R
maskJHU_MD_46_UNC_L
maskJHU_MD_45_UNC_R
maskJHU_MD_34_EC_L
maskJHU_MD_32_ILF_IFOF_L
maskJHU_MD_43_SFOF_R
maskJHU_MD_42_SLF_L
maskJHU_MD_37_Cing_h_R
maskJHU_MD_44_SFOF_L
maskJHU_MD_35_Cing_R
maskJHU_MD_36_Cing_L
maskJHU_MD_31_ILF_IFOF_R
maskJHU_MD_41_SLF_R
maskJHU_MD_13_SCP_R
maskJHU_MD_14_SCP_L
maskJHU_MD_2_PCT
maskJHU_MD_11_ICP_R
maskJHU_MD_8_CST_L
maskJHU_MD_12_ICP_L
maskJHU_MD_7_CST_R
maskJHU_MD_1_MCP
maskJHU_MD_10_mLEM_L
maskJHU_MD_9_mLEM_R
maskJHU_MD_3_gCC
maskJHU_MD_5_sCC
maskJHU_MD_4_bCC
md_global
maskJHU_MD_15_CP_R
maskJHU_MD_16_CP_L
maskJHU_MD_17_ALIC_R
maskJHU_MD_18_ALIC_L
maskJHU_MD_20_PLIC_L
maskJHU_MD_19_PLIC_R
maskJHU_MD_23_ACR_R
maskJHU_MD_24_ACR_L
maskJHU_MD_22_RLIC_L
maskJHU_MD_26_SCR_L
maskJHU_MD_25_SCR_R
maskJHU_MD_29_PTR_R
maskJHU_MD_21_RLIC_R
maskJHU_MD_30_PTR_L
maskJHU_MD_27_PCR_R
maskJHU_MD_28_PCR_L
Features
0.0
0.2
0.4
0.6
0.8
1.0
AUC Value

**Supplementary Figure 7**: **(Figure above) Univariate analysis for the dMRI using the AUC values per feature for the diagnostic and prognostic targets.** The positive classes are MCS and Improved, this if the AUC value if from 0.5 to 1, then the given marker is higher in MCS or Improved patients. If the AUC value is from 0 to 0.5, then the given marker is higher in the UWS or not-Improved group of patients.

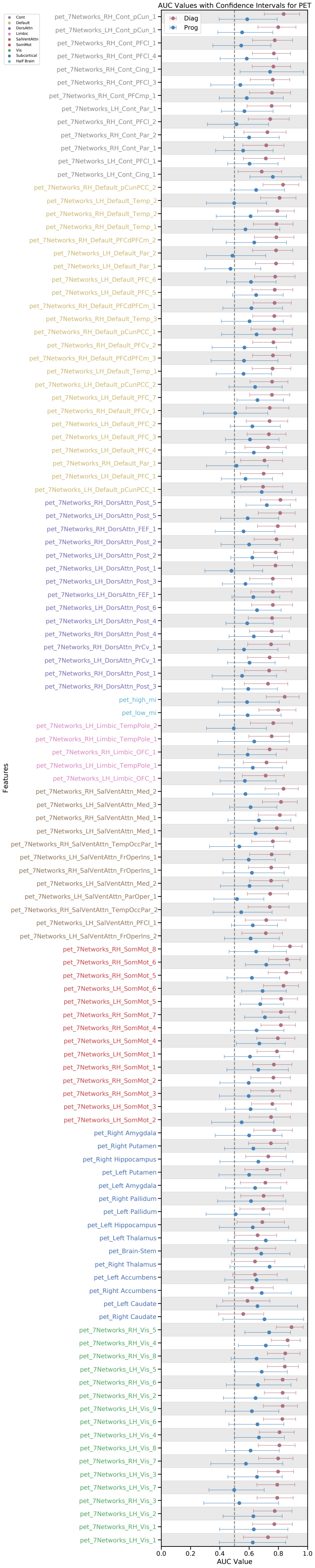
AUC Values with Confidence Intervals for PET
Cont
Default
DorsAttn
Limbic
SalVentAttn
SomMot
Vis
Subcortical
Half Brain
Diag
Prog
Features
pet_7Networks_RH_Cont_pCun_1
pet_7Networks_LH_Cont_pCun_1
pet_7Networks_RH_Cont_PFCl_1
pet_7Networks_RH_Cont_PFCl_4
pet_7Networks_RH_Cont_Cing_1
pet_7Networks_RH_Cont_PFCl_3
pet_7Networks_RH_Cont_PFCmp_1
pet_7Networks_LH_Cont_Par_1
pet_7Networks_RH_Cont_PFCl_2
pet_7Networks_RH_Cont_Par_2
pet_7Networks_RH_Cont_Par_1
pet_7Networks_LH_Cont_PFCl_1
pet_7Networks_LH_Cont_Cing_1
pet_7Networks_RH_Default_pCunPCC_2
pet_7Networks_LH_Default_Temp_2
pet_7Networks_RH_Default_Temp_2
pet_7Networks_RH_Default_Temp_1
pet_7Networks_RH_Default_PFCdPFCm_2
pet_7Networks_LH_Default_Par_2
pet_7Networks_LH_Default_Par_1
pet_7Networks_LH_Default_PFC_6
pet_7Networks_LH_Default_PFC_5
pet_7Networks_RH_Default_PFCdPFCm_1
pet_7Networks_RH_Default_Temp_3
pet_7Networks_RH_Default_pCunPCC_1
pet_7Networks_RH_Default_PFCv_2
pet_7Networks_RH_Default_PFCdPFCm_3
pet_7Networks_LH_Default_Temp_1
pet_7Networks_LH_Default_pCunPCC_2
pet_7Networks_LH_Default_PFC_7
pet_7Networks_RH_Default_PFCv_1
pet_7Networks_LH_Default_PFC_2
pet_7Networks_LH_Default_PFC_3
pet_7Networks_LH_Default_PFC_4
pet_7Networks_RH_Default_Par_1
pet_7Networks_LH_Default_PFC_1
pet_7Networks_LH_Default_pCunPCC_1
pet_7Networks_RH_DorsAttn_Post_5
pet_7Networks_LH_DorsAttn_Post_5
pet_7Networks_RH_DorsAttn_FEF_1
pet_7Networks_RH_DorsAttn_Post_2
pet_7Networks_LH_DorsAttn_Post_2
pet_7Networks_LH_DorsAttn_Post_1
pet_7Networks_LH_DorsAttn_Post_3
pet_7Networks_LH_DorsAttn_FEF_1
pet_7Networks_LH_DorsAttn_Post_6
pet_7Networks_LH_DorsAttn_Post_4
pet_7Networks_RH_DorsAttn_Post_4
pet_7Networks_RH_DorsAttn_PrCv_1
pet_7Networks_LH_DorsAttn_PrCv_1
pet_7Networks_RH_DorsAttn_Post_1
pet_7Networks_RH_DorsAttn_Post_3
pet_high_mi
pet_low_mi
pet_7Networks_LH_Limbic_TempPole_2
pet_7Networks_RH_Limbic_TempPole_1
pet_7Networks_RH_Limbic_OFC_1
pet_7Networks_LH_Limbic_TempPole_1
pet_7Networks_LH_Limbic_OFC_1
pet_7Networks_RH_SalVentAttn_Med_2
pet_7Networks_LH_SalVentAttn_Med_3
pet_7Networks_RH_SalVentAttn_Med_1
pet_7Networks_LH_SalVentAttn_Med_1
pet_7Networks_RH_SalVentAttn_TempOccPar_1
pet_7Networks_LH_SalVentAttn_FrOperIns_1
pet_7Networks_RH_SalVentAttn_FrOperIns_1
pet_7Networks_LH_SalVentAttn_Med_2
pet_7Networks_LH_SalVentAttn_ParOper_1
pet_7Networks_RH_SalVentAttn_TempOccPar_2
pet_7Networks_LH_SalVentAttn_PFCl_1
pet_7Networks_LH_SalVentAttn_FrOperIns_2
pet_7Networks_RH_SomMot_8
pet_7Networks_RH_SomMot_6
pet_7Networks_RH_SomMot_5
pet_7Networks_LH_SomMot_6
pet_7Networks_LH_SomMot_5
pet_7Networks_RH_SomMot_7
pet_7Networks_RH_SomMot_4
pet_7Networks_LH_SomMot_4
pet_7Networks_LH_SomMot_1
pet_7Networks_RH_SomMot_1
pet_7Networks_RH_SomMot_2
pet_7Networks_RH_SomMot_3
pet_7Networks_LH_SomMot_3
pet_7Networks_LH_SomMot_2
pet_Right Amygdala
pet_Right Putamen
pet_Right Hippocampus
pet_Left Putamen
pet_Left Amygdala
pet_Right Pallidum
pet_Left Pallidum
pet_Left Hippocampus
pet_Left Thalamus
pet_Brain-Stem
pet_Right Thalamus
pet_Left Accumbens
pet_Right Accumbens
pet_Left Caudate
pet_Right Caudate
pet_7Networks_RH_Vis_5
pet_7Networks_RH_Vis_4
pet_7Networks_RH_Vis_8
pet_7Networks_LH_Vis_5
pet_7Networks_RH_Vis_6
pet_7Networks_RH_Vis_2
pet_7Networks_LH_Vis_9
pet_7Networks_LH_Vis_6
pet_7Networks_LH_Vis_4
pet_7Networks_LH_Vis_8
pet_7Networks_RH_Vis_7
pet_7Networks_LH_Vis_3
pet_7Networks_LH_Vis_7
pet_7Networks_RH_Vis_3
pet_7Networks_LH_Vis_2
pet_7Networks_RH_Vis_1
pet_7Networks_LH_Vis_1
0.0
0.2
0.4
0.6
0.8
1.0
AUC Value

**Supplementary Figure 8**: **(Figure above) Univariate analysis for the PET using the AUC values per feature for the diagnostic and prognostic targets.** The positive classes are MCS and Improved, this if the AUC value if from 0.5 to 1, then the given marker is higher in MCS or Improved patients. If the AUC value is from 0 to 0.5, then the given marker is higher in the UWS or not-Improved group of patients.

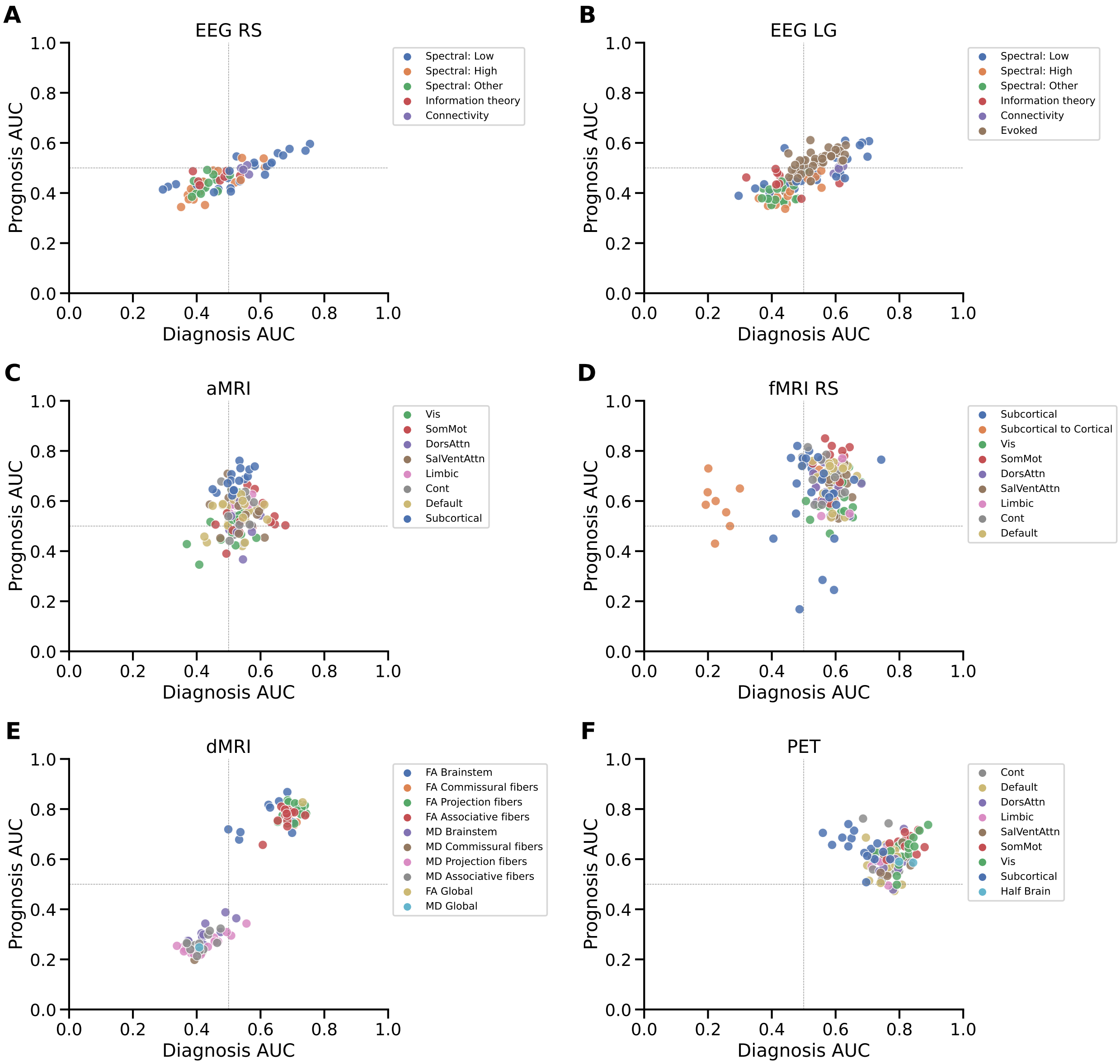


**Supplementary Figure 9**: **Relationship between the AUC values per feature for the diagnostic and prognostic target.** The positive classes are MCS and Improved, this if the AUC value if from 0.5 to 1, then the given marker is higher in MCS or Improved patients. If the AUC value is from 0 to 0.5, then the given marker is higher in the UWS or not-Improved group of patients.

#### Feature importances

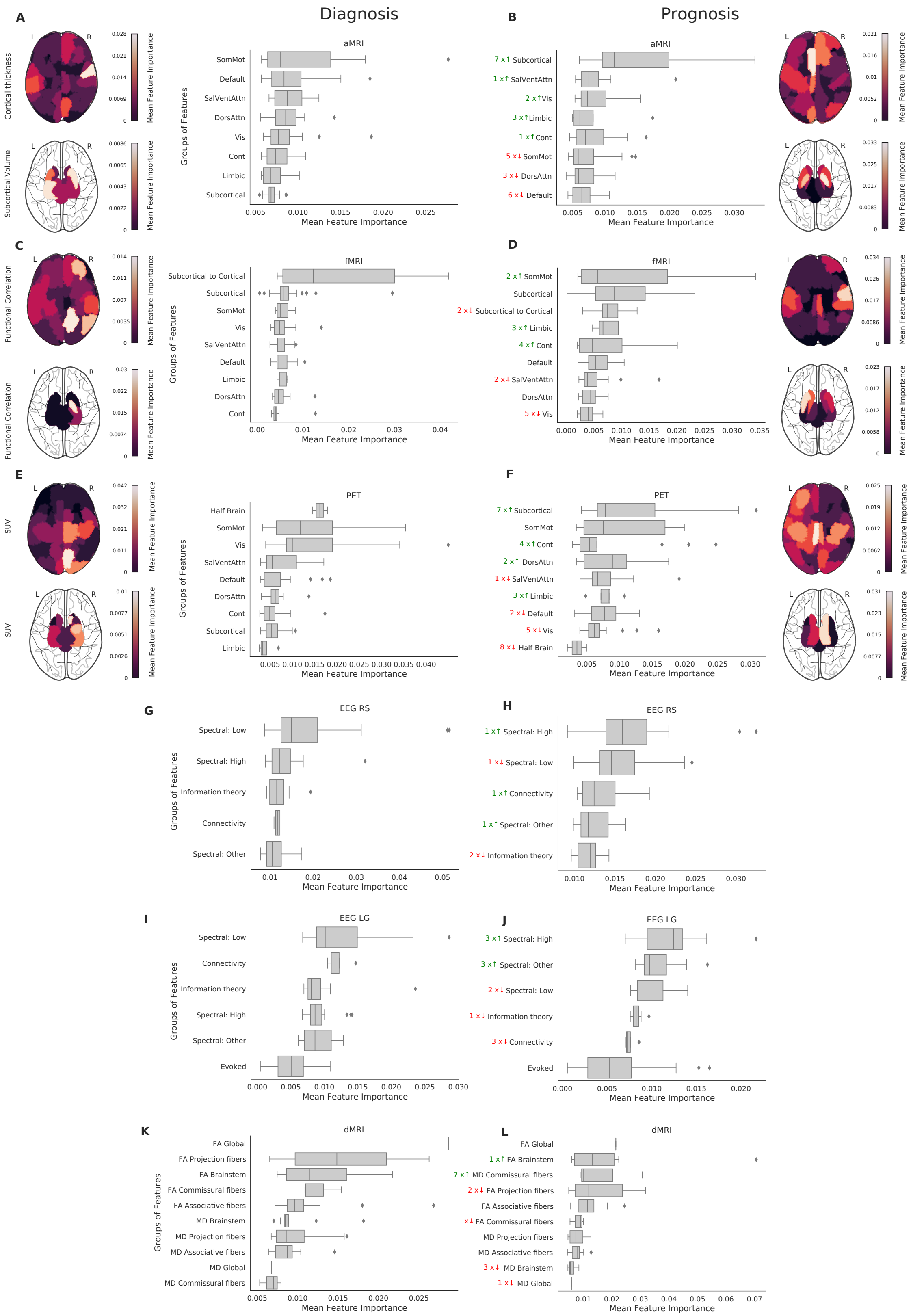


**Supplementary Figure 10**: (caption below)

**Supplementary Figure 10**: **(Figure above) Reordering of the feature importance distributions per group of features for diagnosis and prognosis.** The ranking of the groups of features is informative and not the values of the importance which differs from one model to another. A) Feature importances of the diagnostic prediction using the anatomical MRI scan. On the brain plots the feature importances of the cortical thickness and the subcortical volume are shown. The brain plots from four different views are given in Supplementary Figures 4 and 5. The bar plot shows the feature importances split into cortical networks and subcortical regions. B) Same as A) but for the prognostic prediction. C) Feature importances of the diagnostic prediction using the resting state functional MRI scan. On the brain plots the feature importances of the within cortical and within subcortical functional connectivity per region of interest. The bar plot shows the feature importances split into within subcortical functional connectivity, within cortical connectivity subdivided into the 7 cortical networks, and subcortical to cortical functional connectivity. D) Same as C) but for the prognostic prediction. E) Feature importances of the diagnostic prediction using the FDG PET scan. On the brain plots the feature importances of the metabolic activity per cortical or subcortical region of interest are shown. The bar plot shows the feature importances split into cortical networks and the subcortical regions, as well as the importance of the half-brain (left or right hemisphere) metabolic activity. F) Same as E) but for the prognostic prediction. G) Feature importances of the diagnostic prediction using the resting state EEG recordings. The bar plot shows the feature importances split into conceptual families: connectivity (wSMI), information theory (Kolmogorov complexity and permutation entropy), low spectral (delta, theta, and alpha frequency bands), high spectral (beta and gamma bands), and other spectral marker summaries. H) Same as G) but for the prognostic prediction. I) Feature importances of the diagnostic prediction using the Local-Global task-based EEG recordings. The bar plot shows the feature importances split into the same conceptual families as the resting state EEG with the addition of the evoked markers coming from the task-based paradigm. J) Same as I) but for the prognostic prediction. (continues below)

**Supplementary Figure 10**: (continuation) K) Feature importances of the diagnostic prediction using the dMRI scan. The bar plot shows the feature importances split into two measures Fractional Anisotropy (FA) and Mean Diffusivity (MD) which are subdivided into global brain-wide measures and families of tracts: projection fibers, brainstem, commissural fibers, and associative fibers. L) Same as K) but for the prognostic prediction. The arrows next to the feature groups in the bar plots on the right indicate the change in position of the feature groups in the prognosis compared to diagnosis (eg. 3-arrow-up means that the given feature group is 3 positions higher in prognosis compared to the ranking of the feature groups in the diagnosis). Abbreviations: Default Mode Network (Default), Dorsal Attention Network (DorsAttn), Salience/Ventral Attention Network (SalVentAttn), Somato-Motor Network (SomMot), Visual Network (Vis), Limbic Network (Limbic), Control or Frontoparietal Network (Cont), Electroencephalography (EEG), Resting State (RS), Local Global (LG) paradigm, functional Magnetic Resonance Imaging (fMRI), anatomical Magnetic Resonance Imaging (aMRI), diffusion Magnetic Resonance Imaging (dMRI), Positron Emission Tomography (PET).

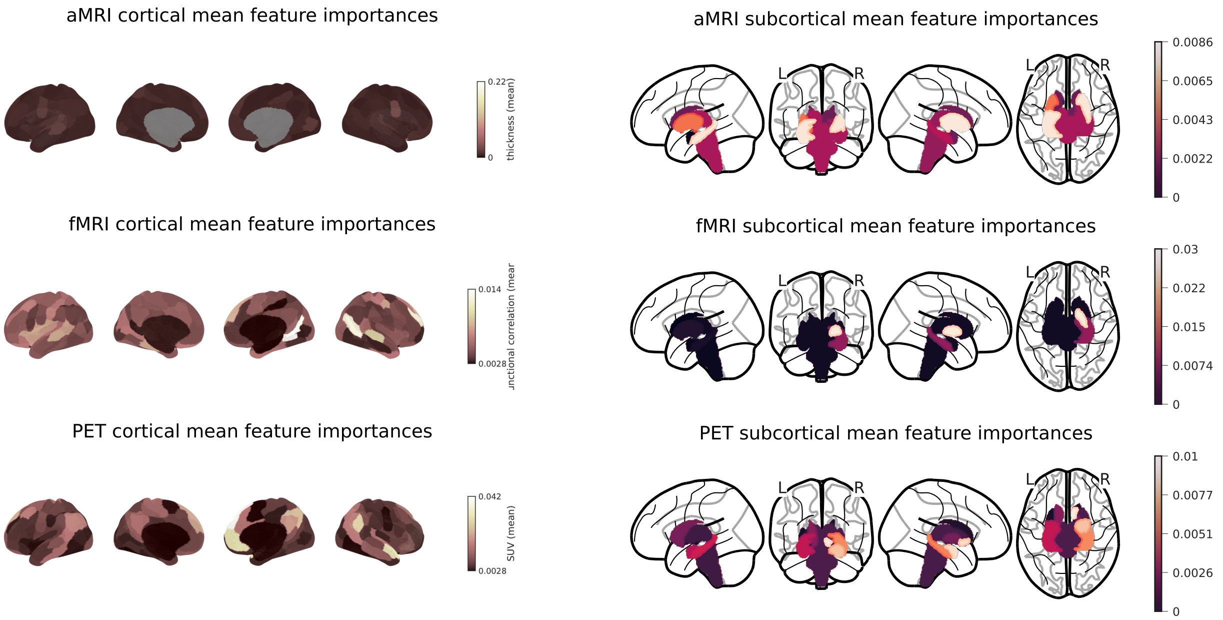


**Supplementary Figure 11**: **Brain plots of the diagnostic cortical (left) and subcortical (right) mean feature importances per region of interest.** In the first row are the feature importances of the aMRI cortical thickness and the subcortical volume, in the second row the fMRI RS feature importance of the within cortical and within subcortical functional connectivity per region of interest, and on the third row the feature importance of the PET metabolic activity. Abbreviations: Resting State (RS), functional Magnetic Resonance Imaging (fMRI), anatomical Magnetic Resonance Imaging (aMRI), Positron Emission Tomography (PET).

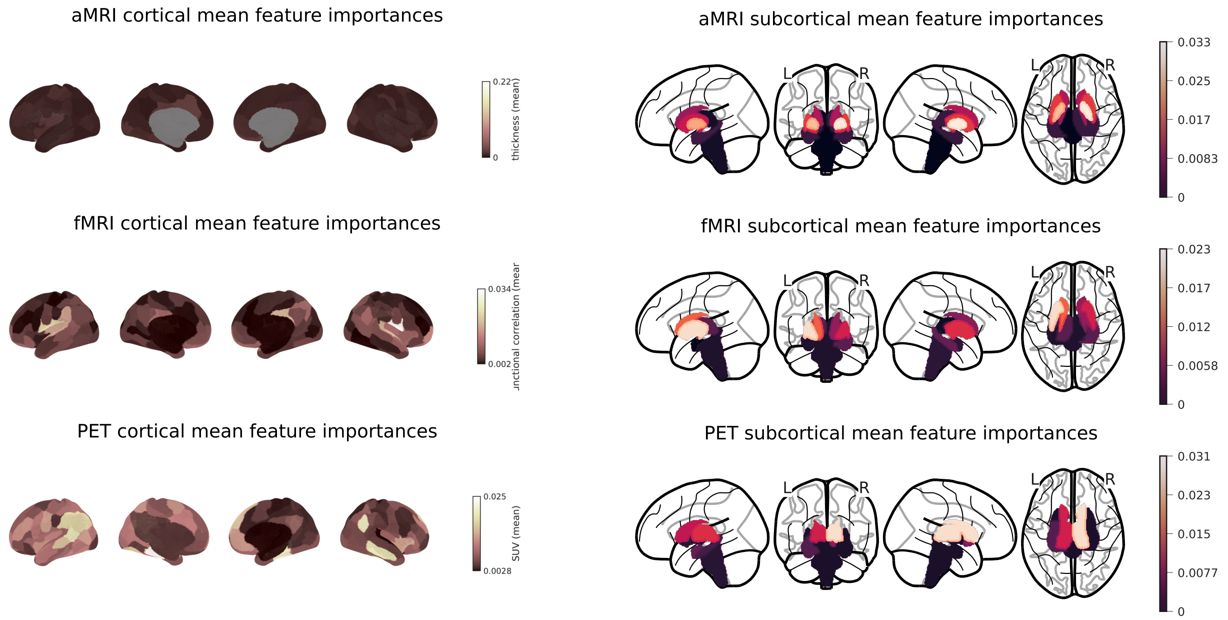


**Supplementary Figure 12**: **Brain plots of the prognostic cortical (left) and subcortical (right) mean feature importances per region of interest.** In the first row are the feature importances of the aMRI cortical thickness and the subcortical volume, in the second row the fMRI RS feature importance of the within cortical and within subcortical functional connectivity per region of interest, and on the third row the feature importance of the PET metabolic activity. Abbreviations: Resting State (RS), functional Magnetic Resonance Imaging (fMRI), anatomical Magnetic Resonance Imaging (aMRI), Positron Emission Tomography (PET).

#### AUC per feature and feature importance relationships

For the AUC analysis, the positive classes are MCS and Improved, this if the AUC value if from 0.5 to 1, then the given marker is higher in MCS or Improved patients. If the AUC value is from 0 to 0.5, then the given marker is higher in the UWS or not-Improved group of patients.

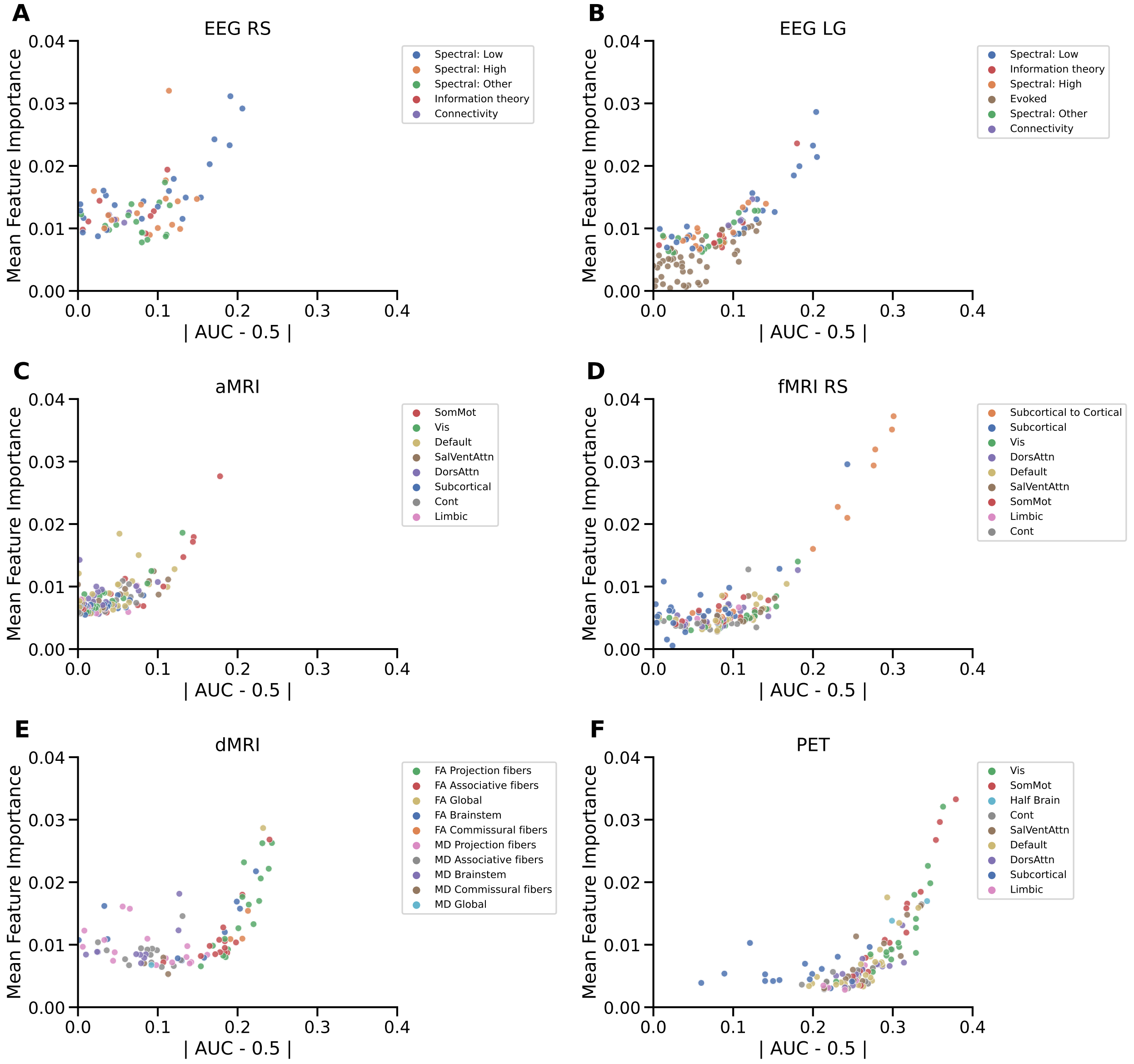


**Supplementary Figure 13**: **Relationship between the AUC values feature and the feature importance values for the diagnostic target.** For the AUC analysis, the positive classes are MCS and Improved, this if the AUC value if from 0.5 to 1, then the given marker is higher in MCS or Improved patients. If the AUC value is from 0 to 0.5, then the given marker is higher in the UWS or not-Improved group of patients.

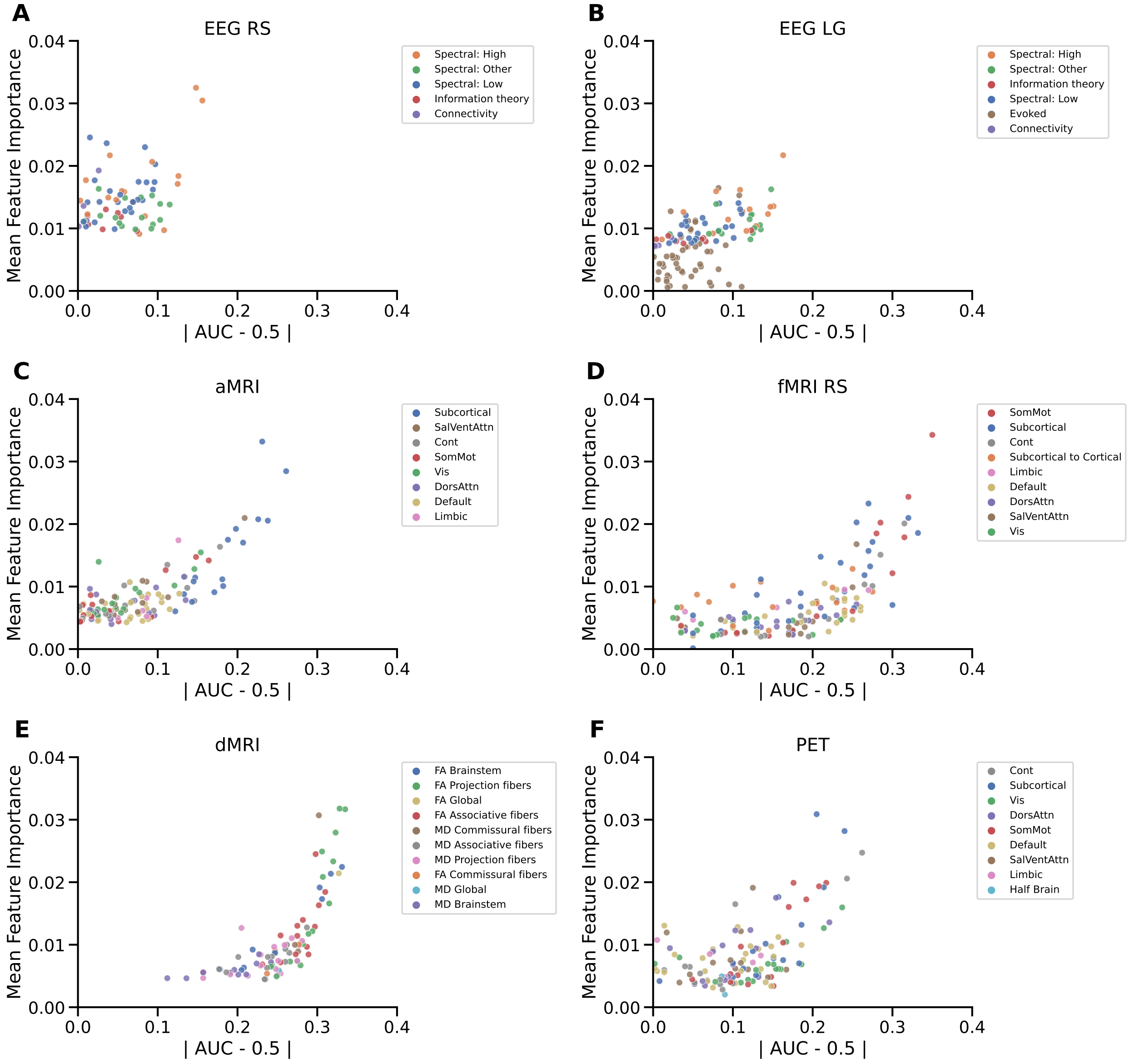


**Supplementary Figure 14**: **Relationship between the AUC values feature and the feature importance values for the prognostic target.** For the AUC analysis, the positive classes are MCS and Improved, this if the AUC value if from 0.5 to 1, then the given marker is higher in MCS or Improved patients. If the AUC value is from 0 to 0.5, then the given marker is higher in the UWS or not-Improved group of patients.

### Suppementary References


Abraham, Alexandre, Fabian Pedregosa, Michael Eickenberg, Philippe Gervais, Andreas Mueller, Jean Kossaifi, Alexandre Gramfort, Bertrand Thirion, and Gael Varoquaux. 2014. "Machine Learning for Neuroimaging with Scikit-Learn." Frontiers in Neuroinformatics 8. https://doi.org/10.3389/fninf.2014.00014.

Avants, B. B., C. L. Epstein, M. Grossman, and J. C. Gee. 2008. "Symmetric Diffeomorphic Image Registration with Cross-Correlation: Evaluating Automated Labeling of Elderly and Neurodegenerative Brain." Medical Image Analysis 12 (1): 26–41. https://doi.org/10.1016/j.media.2007.06.004.

Behzadi, Yashar, Khaled Restom, Joy Liau, and Thomas T. Liu. 2007. "A Component Based Noise Correction Method (CompCor) for BOLD and Perfusion Based fMRI." NeuroImage 37 (1): 90–101. https://doi.org/10.1016/j.neuroimage.2007.04.042.

Cox, Robert W., and James S. Hyde. 1997. "Software Tools for Analysis and Visualization of fMRI Data." NMR in Biomedicine 10 (4-5): 171–78. https://doi.org/10.1002/(SICI)1099-1492(199706/08)10:4/5¡171::AID-NBM453¿3.0.CO;2-L.

Engemann, D.A., Raimondo, F. et al. (2018). Robust EEG-based cross-site and cross-protocol classification of states of consciousness. Brain 141, 3179–3192. https://doi.org/10.1093/brain/awy251

Esteban, Oscar, Ross Blair, Christopher J. Markiewicz, Shoshana L. Berleant, Craig Moodie, Feilong Ma, Ayse Ilkay Isik, et al. 2018. "fMRIPrep 22.0.2." Software. https://doi.org/10.5281/zenodo.852659.

Esteban, Oscar, Christopher Markiewicz, Ross W Blair, Craig Moodie, Ayse Ilkay Isik, Asier Erramuzpe Aliaga, James Kent, et al. 2018. "fMRIPrep: A Robust Preprocessing Pipeline for Functional MRI." Nature Methods. https://doi.org/10.1038/s41592-018-0235-4.

Fonov, VS, AC Evans, RC McKinstry, CR Almli, and DL Collins. 2009. "Unbiased Nonlinear Average Age-Appropriate Brain Templates from Birth to Adulthood." NeuroImage 47, Supplement 1: S102.
https://doi.org/10.1016/S1053-8119(09)70884-5.

Gorgolewski, K., C. D. Burns, C. Madison, D. Clark, Y. O. Halchenko, M. L. Waskom, and S. Ghosh. 2011. "Nipype: A Flexible, Lightweight and Extensible Neuroimaging Data Processing Framework in Python." Frontiers in Neuroinformatics 5: 13.
https://doi.org/10.3389/fninf.2011.00013.

Gorgolewski, Krzysztof J., Oscar Esteban, Christopher J. Markiewicz, Erik Ziegler, David Gage Ellis, Michael Philipp Notter, Dorota Jarecka, et al. 2018. "Nipype." Software.
https://doi.org/10.5281/zenodo.596855.

Greve, Douglas N, and Bruce Fischl. 2009. "Accurate and Robust Brain Image Alignment Using Boundary-Based Registration." NeuroImage 48 (1): 63–72.
https://doi.org/10.1016/j.neuroimage.2009.06.060.

Jenkinson, Mark, Peter Bannister, Michael Brady, and Stephen Smith. 2002. "Improved Optimization for the Robust and Accurate Linear Registration and Motion Correction of Brain Images." NeuroImage 17 (2): 825–41.
https://doi.org/10.1006/nimg.2002.1132.

Jenkinson, Mark, and Stephen Smith. 2001. "A Global Optimisation Method for Robust Affine Registration of Brain Images." Medical Image Analysis 5 (2): 143–56.
https://doi.org/10.1016/S1361-8415(01)00036-6.

Lanczos, C. 1964. "Evaluation of Noisy Data." Journal of the Society for Industrial and Applied Mathematics Series B Numerical Analysis 1 (1): 76–85.
https://doi.org/10.1137/0701007.

Mori, S., Oishi, K., Jiang, H., Jiang, L., Li, X., Akhter, K., Hua, K., Faria, A. v., Mahmood, A., Woods, R., Toga, A. W., Pike, G. B., Neto, P. R., Evans, A., Zhang, J., Huang, H., Miller, M. I., van

Zijl, P., & Mazziotta, J. (2008). Stereotaxic white matter atlas based on diffusion tensor imaging in an ICBM template. NeuroImage, 40(2), 570–582.
https://doi.org/10.1016/J.NEUROIMAGE.2007.12.035

Patriat, Rémi, Richard C. Reynolds, and Rasmus M. Birn. 2017. "An Improved Model of Motion-Related Signal Changes in fMRI." NeuroImage 144, Part A (January): 74–82.
https://doi.org/10.1016/j.neuroimage.2016.08.051.

Power, Jonathan D., Anish Mitra, Timothy O. Laumann, Abraham Z. Snyder, Bradley L. Schlaggar, and Steven E. Petersen. 2014. "Methods to Detect, Characterize, and Remove Motion Artifact in Resting State fMRI." NeuroImage 84 (Supplement C): 320–41.
https://doi.org/10.1016/j.neuroimage.2013.08.048.

Satterthwaite, Theodore D., Mark A. Elliott, Raphael T. Gerraty, Kosha Ruparel, James Loughead, Monica E. Calkins, Simon B. Eickhoff, et al. 2013. "An improved framework for confound regression and filtering for control of motion artifact in the preprocessing of resting-state functional connectivity data." NeuroImage 64 (1): 240–56.
https://doi.org/10.1016/j.neuroimage.2012.08.052.

Sitt, J.D., King, J.-R. et al. (2014). Large scale screening of neural signatures of consciousness in patients in a vegetative or minimally conscious state. Brain 137, 2258–2270.
https://doi.org/10.1093/brain/awu141

Tustison, N. J., B. B. Avants, P. A. Cook, Y. Zheng, A. Egan, P. A. Yushkevich, and J. C. Gee. 2010. "N4itk: Improved N3 Bias Correction." IEEE Transactions on Medical Imaging 29 (6): 1310–20.
https://doi.org/10.1109/TMI.2010.2046908.

Zhang, Y., M. Brady, and S. Smith. 2001. "Segmentation of Brain MR Images Through a Hidden Markov Random Field Model and the Expectation-Maximization Algorithm." IEEE Transactions on Medical Imaging 20 (1): 45–57.
https://doi.org/10.1109/42.906424.
